# Not Such a Long Way Off? Contemporary Artificial intelligence Performance Evaluation on Adult Medicine “Long Cases”

**DOI:** 10.1101/2025.01.25.25321118

**Authors:** Christina Gao, Jamie Bellinge, Shaddy El-Masri, Ivana Chim, Michael Sorich, Ishish Seth, James Gorcilov, Matthew Lim, Liam McCoy, Andrew Vanlint, Lauren Lim, Jessica Stranks, Andrew Zannettino, John Maddison, Stephen Bacchi

**Author notes:** **Corresponding author** Lyell McEwin Hospital, Neurology Department, Haydown Road, Elizabeth Vale SA 5112. **Email:**.

## Abstract

The Royal Australasian College of Physicians (RACP) “Long Cases” assess a trainee’s clinical reasoning beyond what is tested in multiple-choice questions, which large language models (LLMs) have already demonstrated proficiency in. This study evaluated a LLM’s ability to perform a “Long Case” assessment, including history-taking, case presentation, and answering examiner questions. The LLM achieved passing consensus scores of 4-5 out of 6 on five cases, suggesting potential for LLMs in complex clinical evaluations.

## Introduction

Contemporary artificial intelligence (AI), in particular large language models (LLM), are becoming increasingly performant on medical tasks.^1-4^ However, these evaluations often pertain to multiple choice examination questions or curated vignettes.^5-7^ The Royal Australasian College of Physicians (RACP) Divisional Clinical Examinations (DCE) must be passed prior to advanced specialty training. One component of the RACP DCE are “Long Cases”, in which a trainee has one hour with a complex patient to complete a comprehensive evaluation before presenting the patient and their pertinent issues to a set of examiners and answering questions regarding the patient. These “Long Cases” require trainees to demonstrate higher order thought processes, including prioritisation of competing issues and a wholistic approach. Accordingly, evaluation of AI on such an assessment may provide insights into their current capabilities.

The aim of this study was to evaluate the performance of AI, namely an LLM, undertaking DCE “Long Cases”.

## Methods

In this cross-sectional study, an LLM was tested on five hypothetical “Long Cases,” which included three stages: (a) history-taking, (b) synthesizing and presenting the case, and (c) answering examiner questions.

Initially, five hypothetical cases were created by a consultant medical officer. These cases covered common “Long case” medical and psychosocial issues. The cases included conditions not explicitly mentioned in the trainee notes provided in the prompts (see Supplementary Information 1 – Hypothetical “Long Cases”).

Three sets of prompts were developed to guide the LLM through the history-taking, case presentation, and question-answering stages (see Supplementary Information 2 – Prompts). As the LLM could not perform a physical examination, the cases were simulated as if conducted via phone, similar to how exams were held during the COVID-19 pandemic, with physical examination details provided separately.

The evaluation of the LLM was undertaken in a stepwise process (see Figure 1). The LLM, Claude 3.5 Sonnet from *Anthropic*, received the “history” prompt along with basic patient details (name, age, gender, medication list). The LLM had 30 minutes—or until it began discussing management options—to ask any questions. A consultant medical officer, acting as the patient, provided answers that were direct but authentic, without extensive elaboration (see Supplementary Information 3 – History transcripts). Once the history was obtained, the LLM generated a case presentation, which was synthesized into a transcript (see Supplementary Information 4 – Case presentations). If the presentation was shorter than 1000 words, an additional prompt encouraged a more detailed, 2000-word response (standard for a 10-minute presentation). The presentation was recorded and shared with the examiners. Each case was assessed by two examiners: one an RACP fellow (FRACP), the other an advanced trainee or another FRACP. The examiners scored the presentation and provided feedback using standard DCE score sheets. Finally, the examiners each prepared five questions based on the case and presentation, which were sent to the LLM for written responses (see Supplementary Information 5 – Question answering). The “Long Case” scores were then finalized, with both descriptive statistics and narrative feedback provided.

**Figure 1.**
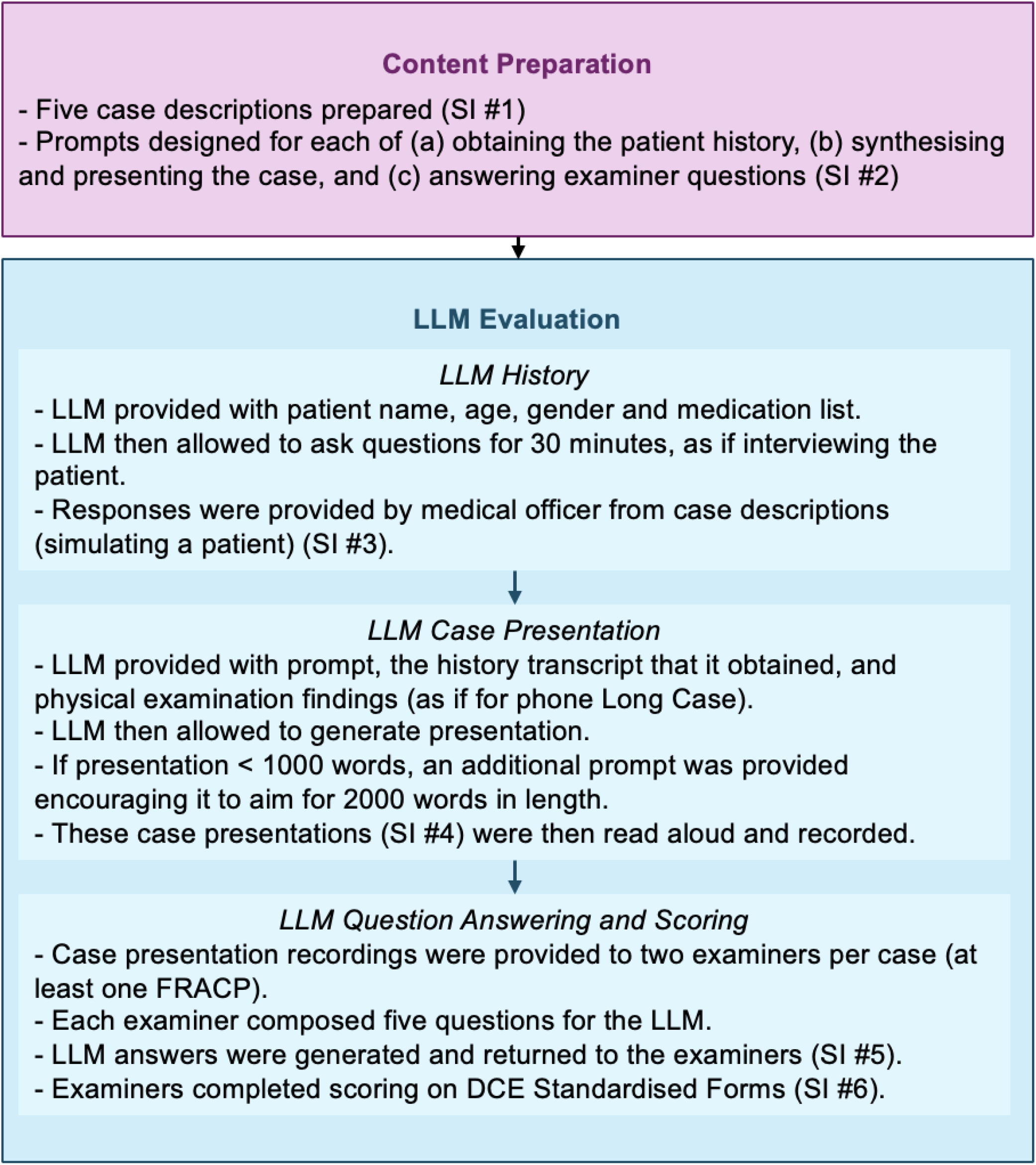
Flowchart detailing LLM “Long Case” evaluation process

## Results

The LLM achieved passing consensus scores of 4/6 on three cases and 5/6 on two cases (see Supplementary Information 6 – Scorer sheets). In the case that included a key condition not explicitly dealt with in the LLM prompt (myasthenia gravis) it scored 4/6. Narrative feedback included repetition in issues list and presenting too many options in response to questions, rather than only those specifically relevant to the patient.

## Discussion

These findings demonstrate a novel means to evaluate LLM performance, and the capabilities of contemporary AI. The performance of LLM on the RACP DCE are of particular significance given recent calls to certify AI similarly to a medical professional.^8^ While limited by the number and use of hypothetical cases (to protect patient confidentiality), this finding remains noteworthy, as in the RACP DCE only two “Long Cases” are typically undertaken by any given trainee on the day of assessment. The use of advanced trainees as examiners is not a limitation, as full case transcripts are available for readers to form their own opinions of the LLM’s performance. These findings give a preview to the type of functionality that such AI may perform in future and, in this context, highlights the need for dedicated education for clinicians on these topics.^9 10^

## Conclusion

This study demonstrates that LLMs can engage effectively in complex medical assessments, such as the RACP “Long Cases”. Further research is needed to refine these systems and explore their integration into real-world clinical settings.

## Data Availability

All data produced in the present work are contained in the supplementary information.

