## Supplementary Information for "Not Such a Long Way Off? Contemporary Artificial intelligence Performance Evaluation on Adult Medicine “Long Cases”"

### Supplementary Information 1 – Hypothetical Long Cases

#### Case 1 – Cystic fibrosis and lung transplant

Mr. David Smith

PROFILE/SOCIAL

- Age: 36 years old
- Home: Lives alone in a small apartment (single-level).
- Home supports: Primarily independent, occasionally receives help from a close friend for shopping and cleaning when unwell.
- Family/Friends Nearby: No immediate family nearby. Father resides in Spain and has been diagnosed with terminal pancreatic cancer.
- Distance from Home (Healthcare Access): Closest hospital is 5 km away; attends transplant center approximately 30 km away for follow-up.
- Social Isolation: Recently reduced social and work activities due to declining health, leading to some degree of isolation.
- Financial Strain: Used to work full-time as a computer programmer but has reduced hours significantly due to health issues, resulting in financial difficulties.
- Relationship Status: Single, unmarried, no children.

FUNCTIONAL

- Mobility: Generally mobile, but experiences breathlessness with mild exertion (e.g., walking more than 200 meters).
- Transport: Does not drive currently due to fatigue and intermittent need for oxygen; uses ride-share services or public transport for essential appointments.
- Handedness: Right-handed.
- Occupation: Computer programmer (currently part-time/freelance).
- Hobbies: Enjoys gaming and reading technology blogs but reports decreased engagement due to fatigue.
- Allergies: nil

CURRENT MEDICATIONS

- Tacrolimus 3 mg twice daily (immunosuppressive therapy post-lung transplant).
- Mycophenolate mofetil 1 g twice daily (immunosuppressive therapy post-lung transplant).
- Prednisolone 5 mg daily (immunosuppressive therapy post-lung transplant).
- Trimethoprim-sulfamethoxazole 160 mg-800 mg daily (infection prophylaxis post-transplant).
- Famciclovir 500 mg daily (for CMV prophylaxis, donor-recipient mismatch).
- Insulin (basal-bolus regimen)
  - Example: Glargine 16 units at bedtime
  - Aspart before meals (dose adjusted based on carbohydrate intake)
- Azithromycin 250 mg three times weekly (prophylactic macrolide for anti-inflammatory effects).
- Pancrelipase 40,000 units with meals and snacks (pancreatic enzyme replacement).
- Fat-soluble vitamins (A, D, E, K) – daily supplementation for CF malabsorption.
- Alendronate 70 mg once weekly (for bone health).
- Paracetamol 1 g as required (for CF-related arthropathy).

MEDICAL HISTORY

1. Cystic Fibrosis (CF)
   - Dx:
     - Diagnosed in early childhood (age 2) following sweat chloride testing (>60 mmol/L) and genetic testing identifying homozygous ΔF508 mutation in the CFTR gene.
     - Prenatal genetic screening was offered to his parents (post-1989).
     - Managed by a specialist CF team (pulmonologist, dietitian, endocrinologist, physiotherapist).
   - Current:
     - Exercise tolerance: Significant limitation due to chronic fatigue and breathlessness with mild exertion.
     - FEV₁: Post-transplant baseline ~65% predicted; recent decline to 50%.
   - Tx:
     - Disease-modifying therapy: Not currently on CFTR modulators post-transplant.
     - Transplant: Bilateral lung transplant 5 years ago for end-stage CF-related lung disease.
     - Genetic counseling: Provided prior to transplant to discuss implications of CF genetics on treatment and family planning.
   - Complications:
     - Respiratory infections:
       - Multiple prior exacerbations and IV antibiotic courses.
       - Hospitalization 2 months ago with a severe respiratory infection; sputum culture grew multiresistant Pseudomonas.
     - Pancreas: Pancreatic insufficiency requiring enzyme replacement.
     - Diabetes mellitus: CF-related diabetes diagnosed 8 years ago, managed with insulin.
     - Sinusitis: Chronic, managed with saline nasal irrigation.
     - Bones: Reduced bone mineral density with osteopenia; on alendronate.
     - Intestinal obstruction: No history of distal intestinal obstruction syndrome (DIOS).
     - Salt crisis: Nil reported.
     - CF arthropathy: Chronic joint pain, requires occasional analgesia.
     - CF liver disease: Nil significant liver dysfunction.
2. Post-Lung Transplant (5 years ago)
   - Dx:
     - Transplant performed for end-stage CF-related lung disease.
     - Donor characteristics: CMV and EBV positive; recipient CMV mismatch, hence on prophylaxis (famciclovir).
   - Current:
     - Graft function:
       - FEV₁: Declined from baseline of 65% to 50% predicted.
       - Medical Research Council (MRC) dyspnea grade: 3 (stops after walking ~100 meters due to shortness of breath).
     - Exercise tolerance: Breathless with mild exertion; struggles with ADLs during exacerbations.
   - Tx:
     - Immunosuppression maintenance: Tacrolimus, mycophenolate mofetil, prednisolone.
     - Infection prophylaxis: Trimethoprim-sulfamethoxazole, famciclovir (for CMV mismatch).
   - Complications:
     - Rejection:
       - Chronic rejection suspected (bronchiolitis obliterans syndrome) with ~20% decrement in FEV₁ over 6 months.
       - No recent biopsy.
     - Primary graft dysfunction (PGD): Nil in the immediate post-transplant period.
     - Acute rejection: Episode in the first year managed with high-dose steroids.
     - Chronic rejection: Progressive decline in graft function consistent with bronchiolitis obliterans syndrome. Re-transplantation may be considered.
     - Infections: Pre-transplant colonization with Pseudomonas; prophylaxis continues.
     - Metabolic syndrome: No hypertension or dyslipidemia; has CF-related diabetes.
     - Malignancy: Nil. Regular skin checks performed.
     - Bone health: Osteopenia post-transplant; on alendronate.
3. Intermittent Intravenous Methamphetamine Use
   - Uses approximately 1 point per occasion, most recent 3 weeks ago.
   - States he uses clean needles to reduce infection risk.
   - No reported complications such as endocarditis or severe withdrawals; acknowledges this is a risky behavior but has limited insight into its potential impact on transplant and overall health.
   - No alcohol, smoking, or other drugs.

ADDITIONAL HISTORY

- Nutritional Challenges: Struggles to maintain weight despite pancreatic enzyme therapy and dietary counseling; BMI is approximately 18 (recent fluctuation was 19 one year ago).
- Vaccinations: Up to date with COVID, flu, pneumococcal vaccines.
- Cancer screening: No skin checks.
- Family Situation: Father is terminally ill with pancreatic cancer in Spain; patient strongly desires to travel overseas to see him one last time.
- Psychosocial Strain: Anxiety about health risks of travel (infection, oxygen needs, insurance coverage) and financial burden.

WHOLISTIC CARE

- Finances: Significant financial strain due to reduced work hours and high medical costs.
- Mental Health: Experiences anxiety and low mood but no formal mental health diagnosis.
- Support Groups: Attends occasional CF/transplant support groups; finds online forums for programmers helpful.
- Partner and Sexual Health: Single, not currently in a relationship.
- Future Planning:
  - Travel Plans: Determined to arrange a trip to Spain despite transplant team’s concerns.
  - Goals: See his father, maintain stable health, and potentially return to full-time work as a programmer.
  - Advanced Care Directive/EPOA/SDM: None in place; no local family members to assist with decision-making.
  - Will: Has not updated personal estate planning.

CURRENT ISSUE

- Biggest Concern: “I’m worried that my health will prevent me from seeing my father before it’s too late. I’m also struggling to balance the financial and medical risks of traveling internationally.”

PHYSICAL EXAMINATION

1. General Appearance
   - Appears pale and mildly cachectic.
   - Thin body habitus, with visible muscle wasting in the upper limbs and shoulders.
   - Alert and oriented ×3 (person, place, time).
2. Vital Signs
   - Temperature: 36.6 °C (oral)
   - Pulse: 92 bpm, regular
   - Respiratory Rate: 20 breaths per minute, slightly labored at rest
   - Blood Pressure: 110/70 mmHg
   - Oxygen Saturation: 94% on room air (occasionally uses low-flow oxygen at home)
3. Anthropometric Measurements
   - Height: ~180 cm
   - Weight: ~58 kg
   - BMI: ~17.9–18.0 (underweight category)
   - Waist Circumference: 78 cm (low subcutaneous fat stores)
4. Cardiovascular Examination
   - Inspection: No jugular venous distension; no visible heaves or lifts.
   - Palpation: Normal apex beat in the 5th intercostal space, mid-clavicular line. No thrills.
   - Auscultation: Dual heart sounds (S1, S2) with no added sounds. No murmurs.
   - Peripheral Pulses: Equal and palpable bilaterally, no peripheral edema.
5. Respiratory Examination
   - Inspection: Thin chest, mild use of accessory muscles on exertion, bilateral thoracotomy scars from transplant.
   - Palpation: Normal chest expansion, slightly reduced on deep inspiration.
   - Percussion: Resonant throughout lung fields, no dullness.
   - Auscultation:
     - Air entry present bilaterally, slightly diminished in lower zones.
     - Mild expiratory wheeze and scattered end-inspiratory crackles at the bases.
     - No obvious rubs.
   - Cough/Sputum: Occasional dry cough; sputum minimal and typically clear.
6. Gastrointestinal Examination
   - Inspection: Abdomen flat, no visible scars, no distension.
   - Auscultation: Normal bowel sounds in all quadrants.
   - Palpation:
     - Soft, non-tender.
     - No hepatosplenomegaly. Liver span within normal limits.
     - No palpable masses.
   - Percussion: Tympanic in the epigastric region, no shifting dullness.
7. Neurological Examination
   - Higher Functions: Alert, oriented; speech normal.
   - Cranial Nerves: Grossly intact, no diplopia, no facial weakness.
   - Motor System: Mild reduction in muscle bulk, particularly in proximal upper and lower limbs; strength 4/5 on manual testing.
   - Reflexes: Normal and symmetric (2+).
   - Sensation: Intact to light touch and pinprick in all four limbs.
   - Coordination: No cerebellar signs; able to perform finger-nose testing with mild intention tremor.
8. Other Notable Findings
   - Skin: Pale, slightly dry; no suspicious lesions.
   - Extremities: Mild digital clubbing. Nails well-groomed, no cyanosis at rest.
   - Musculoskeletal: Mild arthritic changes in hands, no gross deformities.

#### Case 2 – Heart Failure with reduced Ejection Fraction and falls

Mrs. Tran Ng

PROFILE/SOCIAL

- Age: 85 years old
- Home: Lives alone, fiercely independent, in her own house (no stairs)
- Family/Friends: No family; few friends. Limited social support network. Husband died 4 years ago.
- Smoking: Currently an active smoker (~10 cigarettes/day)
- Occupation/Interests: Enjoys gardening; minimal desire for medical intervention

FUNCTIONAL

- Mobility/ADLs: Generally manages her daily activities but acknowledges increased difficulty maintaining her home and garden
- Falls: Two falls at home in the past 6 months (tripped over cat while gardening). No serious injuries reported
- Assistive Devices: Advised to use a 4-wheeled walker (4WW) but declines due to fear of being forced out of her home
- Therapies: Declines physiotherapy and occupational therapy engagement for similar reasons
- Diet: Tea and toast predominantly
- Transport: Drives infrequently; mostly relies on herself for errands
- Hobbies/Interests: Passionate about gardening, reluctant to leave her home environment

CURRENT MEDICATIONS

1. Furosemide 80 mg twice daily – for volume management in heart failure
2. Entresto (sacubitril/valsartan) 97 mg/103 mg twice daily – for afterload reduction and cardiac remodeling benefit in HFREF
3. Bisoprolol 5 mg once daily – for heart failure management and rate control (AF)
4. Spironolactone 50 mg once daily – aldosterone antagonist for HFREF
5. Aspirin 100 mg once daily – secondary prevention post-NSTEMI/stent
6. Atorvastatin 40 mg at night – dyslipidemia management and secondary prevention
7. Apixaban 5 mg twice daily – anticoagulation for paroxysmal atrial fibrillation
8. Salbutamol inhaler 2 puffs PRN – for moderate COPD symptom relief
9. Pregabalin 75 mg once daily – no clear documented indication

Adherent, uses dosette pill organizer.

MEDICAL HISTORY

1. Heart Failure with Reduced Ejection Fraction (HFREF)

Dx:

1. When diagnosed? First diagnosed 2 years ago after presenting to her GP with shortness of breath and undergoing a transthoracic echocardiogram (TTE).
2. How diagnosed? Echocardiogram revealing EF of ~20%; clinical presentation with exertional dyspnea.
3. Managed by who? General medical/cardiology teams at the local hospital; followed by GP.
4. Underlying cause? Ischemic cardiomyopathy (previous NSTEMIs).

Current:

- Symptoms: Recurrent episodes of dyspnea (exercise tolerance 10 meters), peripheral edema, orthopnea.
- Key Functional Investigations:
  - TTE (2 weeks ago): LVEF 20%, mild aortic stenosis
  - BNP: 15,000 pg/mL on most recent admission
- Hospitalizations: Three admissions in the last 4 months, responding to higher diuretic doses.
- Fluid Restriction: 1.25 L/day, reportedly adherent

Tx:

- Current:
  - Furosemide, Entresto, Bisoprolol, Spironolactone per standard HFREF regimen.
- Previous:
  - No device therapy (ICD/CRT).
  - No recent IV diuretic outside of hospital admissions.

2. Ischemic Cardiomyopathy / NSTEMIs

Dx:

1. When diagnosed? Two separate NSTEMIs: one 2 years ago, another 12 years ago.
2. How diagnosed? Clinical presentation (chest pain, elevated troponins, ECG changes).
3. Managed by who? Cardiology with percutaneous coronary intervention (PCI) and stent placement.
4. Underlying cause? Coronary artery disease (smoking, dyslipidemia, hypertension).

Current:

- Stents in LAD and circumflex.
- Symptoms: Stable angina not prominently reported; overshadowed by HF symptoms.

Tx:

- Current:
  - Aspirin, Atorvastatin for secondary prevention.
- Previous:
  - Drug-eluting stents placed for both NSTEMI events.
  - Initially on dual antiplatelet therapy; now on aspirin alone.

3. Paroxysmal Atrial Fibrillation

Dx:

1. When diagnosed? Identified during her most recent hospital admission (~2 weeks ago) on telemetry/ECG.
2. How diagnosed? ECG showing intermittent irregularly irregular rhythm, confirmed by in-hospital monitoring.
3. Managed by who? Cardiology/medical team in hospital; ongoing GP oversight.
4. Underlying cause? Likely structural heart disease from ischemic cardiomyopathy.

Current:

- Symptoms: Occasionally palpitations; no significant syncope.
- Key Functional Investigations: Baseline ECG, rate vs. rhythm control considerations.

Tx:

- Current: Apixaban 5 mg twice daily for stroke prophylaxis; Bisoprolol for rate control.
- Previous: No prior anticoagulation or antiarrhythmic therapy.

4. Moderate COPD

Dx:

1. When diagnosed? Approximately 5 years ago with chronic cough and dyspnea.
2. How diagnosed? Spirometry consistent with moderate obstruction (FEV1 ~50–60% predicted).
3. Managed by who? GP, occasional nurse-led spirometry checks; no specialist respiratory referral noted.
4. Underlying cause? Long-term smoking (~10/day currently).

Current:

- Symptoms: Mild to moderate dyspnea; occasionally uses salbutamol.
- Key Functional Investigation: Last known spirometry indicating moderate obstructive pattern.

Tx:

- Current: Salbutamol inhaler PRN 3-4 times per week; continues to smoke.
- Previous: No inhaled steroids or LABA/LAMA therapy documented; no pulmonary rehab.

5. Falls

Dx/Start:

- First fall? First recorded fall was 6 months ago.
- Ever seen in falls clinic? No falls clinic referral or assessment.

Current:

- Most recent fall: Occurred 2 weeks ago while gardening.
  - Before the fall: No dizziness, loss of consciousness (LoC), or chest pain (CP). Tripped over her cat while moving through the garden.
  - After the fall: Sustained minor bruises; no fractures. Able to get up independently after 5–10 minutes.

Modifiable Risk Factors:

- Vision: Has not seen an optometrist recently. May have impaired vision.
- Lighting: Dim lighting in areas around the house.
- Footwear: Prefers slip-on shoes without support.
- Rails/Stairs: Limited handrails near steps and garden pathways.
- Polypharmacy: On multiple medications, including sedating pregabalin and medications that could cause postural hypotension (e.g., furosemide, bisoprolol).
- Alcohol: Denies alcohol use.

Unmodifiable Risk Factors:

- Age: 85 years old.
- Previous Falls: Two falls in the past 6 months.
- Dementia: None reported.

Tx:

- AH (Allied Health): Declines physiotherapy for exercise/balance programs and occupational therapy for home modification. No podiatry or optometry referral.
- Reduce Polypharmacy: Considered but not implemented due to clinical needs for current medications.
- Complications/Follow-up:
  - Loss of confidence: Limited; still prefers gardening but has become more cautious.
  - Activities: No significant reduction beyond slower movements.
  - Bone health: No recent DXA scan or osteoporosis management; at risk of fragility fractures.

WHOLISTIC CARE

- Social Context: Lives alone, no family, few friends; strongly resistant to relocating or in-home support.
- Vaccines: Up to date with covid, influenza, pneumococcal
- Cancer screening: no longer doing bowel cancer screening or mammograms
- Mental Health: Potential for social isolation; no formal mental health evaluation.
- Future Planning:
  - HF Prognosis: Advanced disease (EF ~20%, BNP 15,000, recurrent exacerbations).
  - Advance Care Directive/Will: None; limited awareness of palliative options.
  - Home Support Services: Not currently utilized.

ADDITIONAL HISTORY

- Living Situation Concern: Insists on remaining at home despite recurrent HF admissions and limited social support.
- Legal/Advance Planning: No will or advanced care directive. Limited insight into her advanced heart disease prognosis.
- Unintentional Weight Loss: Reports a 5 kg weight loss over the past few months, which she attributes to reduced appetite and feeling unwell.

CURRENT ISSUE

- Biggest Concern: “I don’t want to leave my home or keep going to the hospital. I’d rather just stay here in my garden and be left alone.”

PHYSICAL EXAMINATION

1. General Appearance
   - Alert, elderly woman; mild respiratory effort at rest.
   - Appears frail but adamant about independence.
2. Vital Signs
   - Temperature: 36.5 °C
   - Pulse: 90 bpm, irregularly irregular
   - Respiratory Rate: 18 breaths/min
   - Blood Pressure: 110/70 mmHg
   - Oxygen Saturation: 94% on room air
3. Anthropometric Measurements
   - Height: ~160 cm
   - Weight: ~52 kg
   - BMI: ~20 (low-normal)
   - Waist Circumference: 85 cm
4. Cardiovascular Examination
   - Inspection: Mild jugular venous distension noted.
   - Palpation: Displaced apex
   - Auscultation:
     - S3 gallop (HF).
     - Soft systolic murmur at aortic area
     - Irregularly irregular rhythm
5. Respiratory Examination
   - Inspection: Mild use of accessory muscles.
   - Palpation: Normal expansion, no focal consolidation.
   - Percussion: Resonant throughout.
   - Auscultation: Bibasal crackles + occasional wheeze
6. Abdominal Examination
   - Inspection: Abdomen flat.
   - Palpation: No organomegaly, no significant tenderness.
   - Auscultation: Normal bowel sounds.
7. Neurological Examination
   - Mental Status: Alert/oriented ×3,
   - Motor/Sensory: No focal deficits.
   - Coordination/Gait: Mildly unsteady but ambulates independently.
8. Other
   - Skin: Thin, some bruising on forearms.
   - Peripheral Edema: Mild pedal edema

#### Case 3 – Myasthenia gravis and osteoporosis

Mrs. Emily Brown

PROFILE/SOCIAL

- Age: 61 years old
- Home: Lives at home with her husband in a single-story house (no stairs).
- Home supports: Primarily supported by her husband for cooking, cleaning, and shopping. No formal supports currently in place.
- Family/Friends Nearby: Son lives 20 minutes away; sister lives interstate, 2 hours by plane.
- Distance from Home: Lives 10 km from the nearest hospital and 5 km from her general practitioner.
- Social Isolation: Reduced social interactions due to limited mobility and activity caused by pain and fatigue. Feels isolated and no longer attends social or community events.

FUNCTIONAL

- Mobility: No gait aids required but has experienced two falls at home in the past three months due to lower limb weakness. Pain from vertebral fracture significantly limits walking and standing, leading to dependence on her husband for many daily activities. Premorbidly, she enjoyed daily 2 km walks. Now primarily sedentary.
- Transport: No longer drives due to intermittent double vision and fatigue. Relies on her husband or public transport.
- Handedness: Right-handed.
- Occupation: Retired teacher (retired 5 years ago).
- Hobbies: Previously enjoyed gardening and reading. Gardening has stopped entirely due to pain and fatigue. Reading has decreased due to mood and concentration issues.

CURRENT MEDICATIONS

- Prednisolone 25 mg daily (for myasthenia gravis).
- Mycophenolate 500 mg twice daily (for myasthenia gravis; higher doses intolerable).
- Calcium and vitamin D supplementation (for osteoporosis).
- Denosumab 60 mg subcutaneous injection every 6 months (for osteoporosis).
- Duloxetine 60 mg daily (for depression and pain).
- Paracetamol 1 g four times daily (for vertebral fracture pain).
- Ibuprofen 400 mg three times daily (for vertebral fracture pain).
- Continuous release tramadol 100 mg twice daily (for vertebral fracture pain).
- Esomeprazole 40 mg daily (for GORD and gastric protection with NSAIDs).
- Trimethoprim-sulfamethoxazole 160 mg-800 mg BD Monday/Wednesday/Friday (for Pneumocystis jirovecii pneumonia prophylaxis).
- Salbutamol inhaler as required (for asthma).

MEDICAL HISTORY

1. Myasthenia Gravis (MG)
   - Dx:
     - Diagnosed 5 months ago.
     - Diagnosis confirmed with AChR antibody positivity and clinical presentation of neck weakness and swallowing difficulties. Had hospitalisation at that time. Initial presentation required IVIg therapy. Chest CT showed no thymoma.
     - Managed by a neurologist.
     - Underlying cause: Autoimmune disorder.
   - Current:
     - Symptoms include intermittent fatigue, lower limb weakness, and swallowing difficulties. Attempts to reduce prednisolone below 25 mg have resulted in symptom recurrence. One episode requiring hospital attendance at diagnosis.
     - Key functional investigations: Normal respiratory function (no FVC decrement).
   - Tx:
     - Current: Prednisolone 25 mg daily (unable to taper lower without symptom recurrence). Mycophenolate 500 mg twice daily (higher doses intolerable due to gastrointestinal side effects).
     - Previous: IVIg at diagnosis.
2. Osteoporosis with Fragility Fractures
   - Dx:
     - Diagnosed 2 years ago after a Lumbar vertebral fragility fracture confirmed on X-ray.
     - Managed by her general practitioner (GP).
     - Underlying cause: Likely multifactorial (postmenopausal, prednisolone use).
   - Current:
     - Ongoing severe back pain rated 7/10, which significantly limits mobility and interferes with daily activities such as cleaning, cooking, and dressing. Pain is worse with sitting or standing for long periods and disrupts sleep.
     - Key functional investigations: DEXA scan 18 months ago showing T-score -2.8 at the lumbar spine.
   - Tx:
     - Current: Calcium and vitamin D supplementation, denosumab every 6 months. Regular paracetamol, ibuprofen, and continuous release tramadol for pain management.
     - Previous: Nil specific interventions other than supplementation.
3. Falls
   - Dx:
     - Two falls in the past three months due to lower limb weakness from myasthenia gravis. Most recently three weeks ago. No fractures or head injuries sustained.
   - Current:
     - At increased risk of falls due to lower limb weakness and back pain. Reduced confidence in mobility.
   - Tx:
     - No formal intervention to reduce falls risk.
4. Depression
   - Dx:
     - Diagnosed 15 years ago based on clinical history.
     - Managed by her general practitioner, with occasional input from a psychologist.
     - Underlying cause: Likely multifactorial (chronic illness, psychosocial stressors).
   - Current:
     - Symptoms have worsened over the past three months, including low mood, lack of motivation, fatigue, tearfulness, and feelings of social isolation.
   - Tx:
     - Current: Duloxetine 60 mg daily.
     - Previous: Intermittent counseling sessions with a psychologist (stopped 2 years ago).
5. Obesity
   - Dx:
     - Diagnosed 5 years ago based on BMI >30.
     - Managed by her general practitioner.
     - Underlying cause: Likely related to sedentary lifestyle and high-calorie diet.
   - Current:
     - No specific symptoms related to obesity but ongoing weight gain and lack of activity exacerbate her other conditions.
     - Key functional investigations: BMI 32.7 (Weight 89 kg, Height 165 cm).
   - Tx:
     - Current: No formal dietary or exercise program in place.
     - Previous: Brief trial of a weight loss program 3 years ago (discontinued).
6. Gastroesophageal Reflux Disease (GORD)
   - Dx:
     - Diagnosed 3 years ago based on symptoms of heartburn and regurgitation.
     - Managed by her general practitioner.
     - Underlying cause: Likely related to obesity and prednisolone use.
   - Current:
     - Symptoms are well-controlled with medication. Occasional mild breakthrough symptoms.
   - Tx:
     - Current: Esomeprazole 40 mg daily.
     - Previous: Initial trial of ranitidine (discontinued due to lack of efficacy).
7. Asthma
   - Dx:
     - Diagnosed at age 15 based on clinical symptoms and spirometry.
     - Managed by her general practitioner.
     - Underlying cause: Atopic predisposition.
   - Current:
     - Minimal symptoms, uses salbutamol less than once per month. No ICU admissions or severe exacerbations.
     - Key functional investigations: Normal FEV1/FEV1 ratio on recent spirometry.
   - Tx:
     - Current: Salbutamol inhaler as required.
     - Previous: Nil significant treatments required.

ADDITIONAL HISTORY

- Pain and ADLs: Severe pain from vertebral fracture limits ability to perform daily activities independently, such as cooking, cleaning, and dressing. Pain is exacerbated by standing or sitting for long periods.
- Sexual Health: No longer sexually active due to pain and fatigue.
- Preventative Health:
  - Vaccinations: Up to date on influenza and COVID vaccines.
  - Cancer screening: Normal mammogram 1 year ago, normal cervical screening 2 years ago.
- Diet: High-calorie diet, frequent takeaway meals, low intake of fresh fruits and vegetables.
- Exercise: Minimal due to fatigue and back pain.
- Allergies: Nil.
- Alcohol: Nil
- Smoking: Nil
- Recreational drugs: Nil

WHOLISTIC CARE

- Finances: Stable. Husband’s pension and savings provide sufficient support.
- Mental Health: Depression symptoms have worsened. No current counseling or support group involvement.
- Support Groups: Open to exploring MG or osteoporosis support groups.
- Partner and Sexual Health: Married, but no longer sexually active due to pain and fatigue.
- Menopause: Postmenopausal for 10 years.
- Pregnancy History: Two children, no pregnancy complications.
- Future Planning:
  - Travel Plans: Would like to visit her sister interstate but feels uncertain about managing fatigue during travel.
  - Goals: Wishes to regain strength and independence to return to gardening and light exercise.
  - Advanced Care Directive/EPOA/SDM: Husband is her informal substitute decision-maker. No formal advanced care directive in place.
  - Will: Updated 5 years ago.

MAIN ISSUE

- Biggest Concern: "I’m worried about how much longer I’ll be able to manage my day-to-day activities. The pain is unbearable, and I feel like I’m losing my independence and my connection with the world."

Physical Examination

General Appearance

- Alert, cooperative but appears fatigued.
- Mildly obese with noticeable discomfort when sitting upright.
- Moves carefully, guarding her lower back region.

Vital Signs

- Temperature: 36.7 °C (98.1 °F)
- Pulse Rate: 86 beats per minute, regular
- Blood Pressure: 142/84 mmHg (right arm, sitting)
- Respiratory Rate: 16 breaths per minute
- Oxygen Saturation: 97% on room air

Anthropometrics

- Height: 165 cm
- Weight: 89 kg
- BMI: 32.7 kg/m² (Obese category)
- Waist Circumference: 105 cm

Cardiovascular Examination

- Inspection: No visible pulsations or abnormalities on the precordium.
- Palpation:
  - Apex beat palpable in the 5th intercostal space, mid-clavicular line.
  - No palpable thrills or heaves.
- Auscultation:
  - Heart sounds S1 and S2 normal, no murmurs, rubs, or gallops detected.
  - Regular rate and rhythm.
- Peripheral Vascular:
  - Peripheral pulses (radial, dorsalis pedis, posterior tibial) palpable and symmetrical.
  - No pedal edema.
  - Capillary refill <2 seconds.

Respiratory Examination

- Inspection:
  - Normal chest shape, no obvious use of accessory muscles at rest.
  - Respiratory effort mildly increased when moving or speaking for prolonged periods
- Palpation:
  - Chest expansion symmetrical.
  - No tenderness on the chest wall.
- Percussion:
  - Resonant note throughout lung fields bilaterally.
- Auscultation:
  - Vesicular breath sounds heard bilaterally with no added sounds (wheeze, crackle).
  - Good air entry, no signs of consolidation or effusion.
  - Mildly reduced breath sounds at bases, possibly due to shallow breathing from back pain.

Gastrointestinal Examination

- Inspection:
  - Abdomen appears mildly protuberant consistent with obesity.
  - No visible scars or abnormal pulsations (appendicectomy scar well-healed, not prominently visible).
- Auscultation:
  - Normal bowel sounds in all four quadrants.
- Palpation:
  - Non-tender epigastric region, no palpable masses.
  - Mild general discomfort noted on deeper palpation, likely secondary to posture and back pain rather than abdominal pathology.
- Percussion:
  - Tympanic note throughout, no organomegaly detected.

Musculoskeletal / Back Examination

- Inspection:
  - Mild kyphotic posture related to vertebral fracture and chronic pain.
  - No muscle wasting apparent, but posture guarded.
- Palpation:
  - Tenderness over the lower thoracic and upper lumbar vertebrae.
  - Paravertebral muscle spasm noted on palpation.
- Range of Motion:
  - Decreased lumbar flexion and extension due to pain.
  - Movement limited by pain rather than joint restriction.

Neurological Examination

- General Observations:
  - Appears fatigued; muscle weakness worsens with sustained activity.
- Cranial Nerves:
  - Mild bilateral ptosis at times, more prominent on prolonged upward gaze (fatigable).
  - Extraocular movements: Mild diplopia on lateral gaze, worsens with sustained gaze.
  - Facial sensation, jaw movement, and facial muscle strength otherwise intact.
  - No dysarthria at rest, though she reports some swallowing difficulties
- Motor System:
  - Tone: Normal in upper and lower limbs, no spasticity or rigidity.
  - Power:
    - Neck flexors: Mild weakness (4-/5), fatigability noted on repeated testing.
    - Proximal upper limb strength: 4/5, with mild fatigability.
    - Proximal lower limb strength: 4/5, reports heaviness especially after walking a short distance.
  - Reflexes: Normal (2+) and symmetrical in biceps, triceps, patellar, and ankle jerks.
  - Coordination: Normal finger-nose and heel-shin testing, though patient tires quickly.
  - Gait: Appears cautious and slightly unsteady, primarily due to lower limb weakness and back pain.
- Sensory Examination:
  - Light touch, pinprick, and proprioception intact in all four limbs.

Skin

- General:
  - Skin is intact with no rashes or ulcers.
  - No bruising, lesions, or striae beyond mild striae possibly related to prolonged steroid use.

#### Case 4 – Perioperative and rural

Mr. Peter Wilson

PROFILE/SOCIAL

- Age: 59 years old
- Heritage: Aboriginal Australian
- Home: Resides in Murray Bridge, South Australia
- Family: Three brothers and two sisters, whom he wants more involved in decision-making.
- Alcohol Use: Historically significant (leading to cirrhosis); currently drinks only on special occasions.
- Smoking: Active cigarette smoker (~15 cigarettes/day), contributing to mild COPD.
- Occupation & Living Situation: Primarily remains near Murray Bridge; dislikes traveling to Adelaide.

FUNCTIONAL

- Mobility/Travel: Independent with daily activities but very reluctant to travel to the city (Adelaide) for medical care.
- ADLs: Manages local tasks; mild breathlessness and fatigue limit heavier activities.
- Transport: Drives himself locally; rarely travels long distances.
- Hobbies/Interests: Not specifically stated, overshadowed by health concerns.

CURRENT MEDICATIONS

1. Metformin XR 1 g once daily – for type 2 diabetes mellitus
2. Gliclazide MR 60 mg once daily – for additional glycemic control
3. Ramipril 5 mg once daily – for blood pressure control and renal protection (CKD)
4. Atorvastatin 20 mg once nightly – for cardiovascular risk reduction
5. Salbutamol (100 µg) inhaler, 2 puffs PRN – for mild COPD symptoms
6. Thiamine 100 mg once daily – supportive therapy given history of alcohol use and cirrhosis

MEDICAL HISTORY

1. Abdominal Aortic Aneurysm (AAA)

Dx:

1. When diagnosed? First detected on an abdominal ultrasound approximately 18 months ago.
2. How diagnosed? Routine imaging (ultrasound) to investigate abdominal pulsations/risk factors.
3. Managed by who? Under the care of a vascular surgery team.
4. Underlying cause? Likely multifactorial atherosclerosis (smoking, hypertension, dyslipidemia).

Current:

- Symptoms: Asymptomatic; no rupture-related symptoms. Anxiety about surgery.
- Key Functional Investigation:
  - Most recent ultrasound 2 weeks ago shows an infrarenal AAA measuring 6.5 cm, having grown 1 cm over the last 6 months.

Tx:

- Current: Vascular surgeons strongly advise operative repair due to rapid expansion and size (>6 cm).
- Previous: Serial ultrasound surveillance every 6 months prior to reaching current threshold for surgical consideration.

2. Child’s A Cirrhosis (Child-Pugh Class A)

Dx:

1. When diagnosed? About 5 years ago during evaluation for abnormal liver function tests (LFTs).
2. How diagnosed? Through imaging (ultrasound), LFT abnormalities, and clinical scoring (Child-Pugh).
3. Managed by who? Local gastroenterologist in conjunction with GP follow-ups.
4. Underlying cause? Long-term excessive alcohol consumption.

Current:

- Symptoms: Currently stable, no evidence of decompensated cirrhosis (no ascites, variceal bleeding, encephalopathy).
- Key Functional Investigation: Periodic LFTs (ALT, AST, INR), which remain relatively stable. No endoscopy history for varices documented.

Tx:

- Current:
  - Abstinent from alcohol except special occasions.
  - Supportive measures (thiamine supplementation, nutritional advice).
- Previous:
  - Historically high alcohol use; no previous variceal interventions or diuretics used.

3. Mild COPD

Dx:

1. When diagnosed? Approximately 3 years ago after presenting with chronic cough and dyspnea.
2. How diagnosed? Clinical history plus spirometry (mild obstruction noted).
3. Managed by who? GP and occasional respiratory nurse visits; no specialist referral documented.
4. Underlying cause? Long-term cigarette smoking (~15/day).

Current:

- Symptoms: Mild exertional breathlessness, occasionally uses accessory muscles.
- Key Functional Investigation: Last spirometry indicated mild airflow limitation (FEV1 ~75% predicted).

Tx:

- Current:
  - Salbutamol inhaler PRN for symptom relief.
  - Ongoing smoking (~15/day).
- Previous:
  - No formal pulmonary rehabilitation or long-acting bronchodilators.
  - No hospital admissions for exacerbation.

4. Type 2 Diabetes Mellitus (15-year history)

Dx:

1. When diagnosed? At age 44, after classic hyperglycemic symptoms and raised fasting BGLs.
2. How diagnosed? Fasting plasma glucose and HbA1c testing by GP.
3. Managed by who? Shared care between GP and diabetes educator; no recent endocrinologist review.
4. Underlying cause? Likely multifactorial (lifestyle, obesity, family history).

Current:

- Glycemic Control:
  - HbA1c: 7.9% (recent).
  - Random BGLs range 7–16 mmol/L.
- Symptoms: Generally tolerates hyperglycemia but is at risk of complications.
- Key Functional Investigations:
  - Moderate CKD (see below).
  - No formal eye checks; risk of retinopathy.
  - Mild peripheral neuropathy (distal sensory loss).
- Diet: Reports frequent fast food; likely suboptimal glycemic control.

Tx:

- Current:
  - Metformin XR 1 g once daily and Gliclazide MR 60 mg once daily.
  - Lifestyle advice (diet/exercise), albeit not fully adhered to.
- Previous:
  - No insulin therapy yet mentioned; no prior diabetic ketoacidosis episodes.

5. Moderate Chronic Kidney Disease

Dx:

1. When diagnosed? Evolving over the past 5–8 years as part of diabetic nephropathy screening.
2. How diagnosed? Rising serum creatinine, declining eGFR on routine diabetic checks.
3. Managed by who? GP, with occasional phone consults to nephrology if concerns worsen.
4. Underlying cause? Primarily diabetic nephropathy; possible contribution from hypertension.

Current:

- Symptoms: Generally asymptomatic but at risk of fluid overload and worsening kidney function.
- Key Functional Investigation: eGFR described as moderate (30–59 mL/min/1.73m²), exact figure not provided in chart.

Tx:

- Current:
  - Ramipril 5 mg once daily for renal protection.
  - Monitoring of electrolytes, creatinine, and eGFR.
- Previous:
  - Conservative management, no dialysis.
  - No previous acute kidney injury episodes documented.

WHOLISTIC CARE

- Social Context: Strong cultural ties in Murray Bridge; reluctant to seek major city-based care. Wishes extended family involvement in decisions.
- Mental Health: Distressed by father’s death after heart surgery; no formal counseling in place.
- Lifestyle Interventions:
  - Smoking cessation urged; still at ~15/day.
  - Alcohol minimal but historically significant.
  - Diet suboptimal for T2DM/CKD.
- Future Planning:
  - AAA Repair: Recommended by vascular team due to size (6.5 cm) and growth rate.
  - Family Meetings: Patient requests a more inclusive approach with his siblings.
  - Advanced Care Directive/EPOA: Not established.
  - Will: Nil.

ADDITIONAL HISTORY

- Desire for Family Involvement: Feels decisions about his aneurysm are being made without consultation of his three brothers and two sisters.
- Psychological Impact: Father died in hospital 10 years ago post–heart surgery; patient fears a similar in-hospital death if he undergoes AAA repair.

CURRENT ISSUE

- Biggest Concern: “I’m scared I’ll end up like my father, who never came home from his heart operation. I don’t want to go to the city, but they say I need this surgery. I just want my family there to help me decide.”

PHYSICAL EXAMINATION

1. General Appearance
   - Alert, cooperative but anxious; mild jaundice not evident.
   - No acute distress.
2. Vital Signs
   - Temperature: 36.4 °C
   - Pulse: 80 bpm, regular
   - Respiratory Rate: 15 breaths/min
   - Blood Pressure: 140/88 mmHg
   - Oxygen Saturation: 96% on room air
3. Anthropometric Measurements
   - Height: 172 cm
   - Weight: 82 kg
   - BMI: 27.7
   - Waist Circumference: 100 cm
4. Cardiovascular Examination
   - Inspection: No jugular venous distension, no peripheral edema.
   - Palpation: Normal apex beat in 5th intercostal space; a palpable expansile mass in the midline abdomen.
   - Auscultation: Normal S1/S2, no audible murmurs, no carotid bruits.
   - Peripheral Pulses: All palpable; no radio-femoral delay.
5. Respiratory Examination
   - Inspection: Normal chest expansion; mild use of accessory muscles on exertion.
   - Palpation: Equal tactile fremitus bilaterally.
   - Percussion: Resonant throughout.
   - Auscultation: Slightly reduced air entry at bases with occasional wheezes.
6. Abdominal Examination
   - Inspection: Abdomen flat; no skin changes or visible pulsations aside from the AAA area.
   - Palpation:
     - Distinct expansile mass (~6–7 cm) infrarenally.
     - Liver edge non-palpable; no splenomegaly.
   - Auscultation: Soft bruit over the epigastric region
7. Neurological Examination
   - Peripheral Neuropathy: Reduced sensation to light touch and vibration in both feet; mildly decreased ankle reflexes.
   - No motor deficits noted.
8. Other
   - Skin: No spider naevi or palmar erythema.
   - No ascites or fluid wave on exam.

#### Case 5 – Type 1 Diabetes mellitus and driving

Mr. John Mitchell

PROFILE/SOCIAL

- Age: 45 years old
- Home: Lives on a small farm in rural South Australia with his wife and one young child.
- Occupation: Farmer (crops and livestock).
- Family/Friends Nearby: Brother in Western Australia (potential kidney donor). Otherwise, limited local family support.
- Alcohol Use: Currently consuming 5–6 standard drinks most days, related to stress and low mood.
- Smoking: Denies current smoking; occasional smoker in early 20s.
- Allergies: Nil known drug allergies.
- Medication Adherence: Admits occasional missed insulin doses, irregular follow-ups due to farm responsibilities.

FUNCTIONAL

- Mobility/Driving:
  - Reliant on driving for farm work (transporting supplies, operating machinery).
  - Fears loss of license if hypoglycemic episodes become problematic or if his medical status deteriorates.
- ADLs: Generally independent, but neuropathy and fatigue impact heavier farm tasks.
- Transport: Own 4WD. No public transport options.
- Hobbies/Interests: Used to enjoy camping/fishing; reduced participation due to mood issues and physical limitations.

CURRENT MEDICATIONS

1. Insulin Therapy (Basal-Bolus Regimen)
   - Insulin Glargine 22 units once daily at bedtime (basal)
   - Insulin Aspart before meals (sliding scale based on carbohydrate intake and pre-meal blood glucose; typically 8–10 units per meal)
2. Antidepressant: Sertraline 50 mg daily (for depression/PTSD) – partial adherence
3. ACE Inhibitor: Ramipril 5 mg daily (renal protection)
4. Lipid-Lowering Agent: Atorvastatin 20 mg nightly (cardiovascular risk reduction)
5. Multivitamins: Occasional use (patient-initiated)
6. Analgesia: Paracetamol 1 g PRN for aches/pains (no regular neuropathic agent)

MEDICAL HISTORY

1. Type 1 Diabetes Mellitus

- Dx
  - Diagnosed at age 12 after classic symptoms (polyuria, polydipsia, weight loss).
  - Initially managed by a pediatric endocrinologist; currently followed by local GP and occasional endocrinologist visits (challenging due to rural location).
- Current
  - Blood Glucose Levels (BGLs): Frequently elevated (>10 mmol/L), sporadic monitoring.
  - HbA1c: Persistently >8.5% (poor control).
  - Symptoms: Occasional mild hypoglycemia without early awareness (hypoglycemic unawareness), raising driving safety concerns.
  - Diet: Attempts carbohydrate counting; often compromised by irregular meals during busy farm work.
  - Foot Checks: Infrequent self-inspection; no formal diabetic foot clinic attendance.
  - Eye Checks: Known progressive retinopathy, overdue for ophthalmology review (see Eye Examination below).
  - Current Pharmacological Treatment: Basal-bolus insulin regimen (details above). No insulin pump.
- Complications
  - Previous DKA: Multiple episodes in late teens/early 20s, but none for over 10 years.
  - Associated Autoimmune Conditions: Denies coeliac disease or thyroid dysfunction (no formal screening in recent years).
  - Microvascular:
    - Retinopathy (proliferative changes).
    - Nephropathy (see CKD below).
    - Neuropathy (distal sensory loss, no foot ulcers yet).
  - Macrovascular:
    - No known ischemic heart disease (IHD), stroke, or peripheral vascular disease (PVD) but has additional risk factors (smoking history, dyslipidemia).
  - Hypoglycemic Episodes: Mild “hypos” occasionally, reduced awareness. No severe events requiring external assistance.

2. Chronic Kidney Disease

- Dx
  - Progressive diabetic nephropathy over many years.
- Current
  - eGFR: 18 mL/min/1.73m² (Stage 4–5 CKD).
  - Considering transplant vs. dialysis; strongly prefers transplant. Brother in WA under evaluation as potential donor.
- Tx
  - Ramipril for renal protection, dietary modifications (low sodium/protein).
  - No dialysis initiated yet due to distance from dialysis centers and patient preference.

3. PTSD & Depression

- Dx
  - PTSD following a severe farm accident 6 years ago; avoids discussing details.
  - Depression long-standing, worsened in past 3 months.
- Current
  - Suicidal ideation involving vehicle use (though no immediate plan).
  - Low mood, anhedonia, increased alcohol use.
- Tx
  - On sertraline 50 mg daily, partial adherence.
  - No regular psychotherapy or counseling sessions.

ADDITIONAL HISTORY

- Main Concern: Maintaining driving privileges to run the farm and support his family.
- Family Obligations: Wife and young child rely on his income and day-to-day farm operations.
- Proximity to Medical Services: Nearest dialysis center is far; specialist appointments require long drives.
- Alcohol Use: 5–6 standard drinks most days, likely contributing to poor glycemic control and depressive symptoms.

WHOLISTIC CARE

- Finances: Dependent on farm income; potential inability to drive threatens livelihood.
- Mental Health: Exacerbated depression and PTSD, with suicidal ideation.
- Support Groups: None locally available; distance and farm duties limit attendance.
- Partner & Family: Wife provides some farm assistance, also caring for their child.
- Future Planning:
  - Renal Replacement Therapy: Prefers transplant over dialysis (brother as potential donor).
  - Driving Concerns: Very anxious about mandatory reporting to authorities if hypos worsen.
  - Advanced Care Directive/EPOA: None in place.
  - Will: Uncertain if a formal will exists.

CURRENT ISSUE

- Biggest Concern: “I’m scared that if my diabetes or kidneys get worse, I won’t be allowed to drive. My family depends on me, and I don’t know how we’d survive.”

PHYSICAL EXAMINATION

1. General Appearance
   - Sun-weathered skin, lean build. Appears tired and somewhat tense.
2. Vital Signs
   - Temperature: 36.5 °C
   - Pulse: 88 bpm, regular
   - Respiratory Rate: 16/min, normal depth
   - Blood Pressure: 138/88 mmHg
   - Oxygen Saturation: 98% on room air
3. Anthropometric Measurements
   - Height: ~178 cm
   - Weight: ~75 kg
   - BMI: ~23.7 (normal range)
   - Waist Circumference: 88 cm
4. Cardiovascular Examination
   - Inspection: No obvious heaves/lifts, no JVP distension.
   - Palpation: Normal apex beat in 5th intercostal space.
   - Auscultation: Normal S1/S2, no murmurs or additional sounds.
5. Respiratory Examination
   - Inspection: Normal chest expansion, no accessory muscle use at rest.
   - Palpation: Normal tactile fremitus.
   - Percussion: Resonant throughout.
   - Auscultation: Clear bilaterally, no crackles/wheeze.
6. Gastrointestinal Examination
   - Inspection: No scars except minor from farm incidents.
   - Auscultation: Normal bowel sounds.
   - Palpation: Abdomen soft, non-tender, no organomegaly.
7. Neurological Examination
   - Cranial Nerves: Grossly intact.
   - Peripheral Neuropathy:
     - Reduced sensation in feet to light touch/vibration, diminished ankle reflexes.
     - No active foot ulcers; some callus formation.
   - Motor Strength: 5/5.
8. Eye Examination
   - Visual Acuity:
     - Right eye: 6/12 with correction
     - Left eye: 6/9 with correction
   - Fundoscopy:
     - Evidence of proliferative diabetic retinopathy: microaneurysms, dot-blot hemorrhages, and neovascularization especially in the right eye.
     - No evidence of vitreous hemorrhage at present, but high risk given proliferative changes.
   - Comment: Patient overdue for ophthalmology review and potential laser therapy.
9. Other
   - Skin: Dry, with mild dryness/cracking on heels.
   - Psychological: Appears low in mood, expresses anxiety about the future and driving.

### Supplementary Information 2 – Prompts

#### History prompt

You are a medical doctor (internal medicine specialist) and you need to perform a comprehensive evaluation of me (a patient). ONLY ASK ONE QUESTION AT A TIME.

This history is comprehensive and must entail all medical conditions, the reason for all medications, and the sociocultural context of the patient. Below, you will be given three sets of information SECTION (1) Background information on the patient including age, gender, height/weight and medication list, SECTION (2) the suggested structure of the information that you need to obtain (the history), including notes on certain specific questions for given conditions, and SECTION (3) tips to make the history taking process more efficient.

SECTION (1) Details of the patient to present are as follows:

Name: [XX]. Age: [XX]. Gender: [XX].

Medication list:

[XX]

SECTION (2) The suggested structure of the presentation is as follows:

[Copied notes from Basic Physician Trainee, including some disease specific presentation details, which included cystic fibrosis, heart failure with ejection fraction, type 1 diabetes mellitus, but did not include content on perioperative considerations, driving, or myasthenia gravis]

SECTION (3) tips to make the history taking more efficient

Remember you have 30 minutes (approximately 100 questions).

This is a conversation. You are speaking to a patient. Ask one question at a time.

You may ask the patient your first question now.

#### Presentation prompt

You are a medical doctor (internal medicine specialist) who has just evaluated a patient, and now need to present them in an exam setting. There is a specific structure to this presentation that would be best adhered to, and usually is approximately 2000 words. Below, you will be given three sets of information SECTION (1) The patient information, SECTION (2) the suggested structure of the presentation, and SECTION (3) tips to make the presentation successful.

SECTION (1) Details of the patient to present are as follows:

Name: [XX]. Age: [XX]. Gender: [XX].

Medication list:

[XX]

[History transcript]

[Examination findings]

SECTION (2) The suggested structure of the presentation is as follows:

[Copy of Basic Physician Trainee notes]

SECTION (3) tips to make the presentation are as follows:

Try to convey more ownership in discussion (i.e. don’t just present all options, choose an option and justify it) (‘I would XYZ’, I would do this, I would do that).

It is important to maintain an overall perspective of the patient’s place in their life and care and priorities. It will not always be possible to do everything, and trade-offs should be acknowledged and considered, prioritising that which helps the patient achieve their aims.

Synthesise all statements (aka word economy) (“ischaemic sounding chest pain”, “no peripheral stigmata of chronic liver disease”)

MENTION EACH MEDICATION THEY ARE TAKING AND ITS INDICATION AT LEAST ONCE.

Use full words, rather than acronyms or abbreviations.

Make it 2000 words. The whole presentation should be in PROSE (not dot points).

You will be saying this out loud in full sentences.

Please now present the case of this patient.

#### Question answering prompt

You are a medical doctor (internal medicine specialist) who has just evaluated a patient, presented their case, and now need to answer questions regarding them and their care in an exam setting. Below, you will be given three sets of information SECTION (1) The patient information, SECTION (2) your presentation regarding the patient, and SECTION (3) suggestions regarding how best to answer the questions.

SECTION (1) Details of the patient to present are as follows:

Name: [XX]. Age: [XX]. Gender: [XX].

Medication list:

[XX]

[History transcript]

[Examination findings]

SECTION (2) Your presentation regarding the patient:

[Presentation]

SECTION (3) suggestions regarding answering the questions:

Try to convey more ownership in discussion (i.e. don’t just present all options, choose an option and justify it) (‘I would XYZ’, I would do this, I would do that). Don’t talk about referring it is within your scope as a specialist.

It is important to maintain an overall perspective of the patient’s place in their life and care and priorities. It will not always be possible to do everything, and trade-offs should be acknowledged and considered, prioritising that which helps the patient achieve their aims.

Keep answers to 200 to 300 words in length.

Answers should be in prose. You will be saying this out loud.

Are you ready to answer questions regarding the case?

### Supplementary Information 3 – History transcripts

#### Case 1 – Cystic fibrosis and lung transplant

**Doctor:** Good morning, Mr. Smith. I'm Dr. Claude, an internal medicine specialist. Thank you for coming in today. I'd like to take some time to understand your medical history comprehensively. I may need to interrupt occasionally to ensure I capture all the important details, and I'll be mindful of our time. Is it alright if we begin?
**Patient:** Sure.

**Doctor:** First, could you please list all of the medical conditions that you are aware of having at the moment in a very brief, dot-point fashion?
**Patient:**

- Cystic fibrosis
- Lung transplant
- Intravenous drug use

**Doctor:** I understand. Thank you for sharing that. Before we discuss these conditions in detail, have you ever been diagnosed with high blood pressure, diabetes, heart problems, or experienced any falls?
**Patient:** No to all of these, except diabetes, which I have because of my CF.

**Doctor:** Thank you for clarifying that. What is your main concern currently regarding your health?
**Patient:** I’m worried that my health will prevent me from seeing my father before it’s too late. I’m also struggling to balance the financial and medical risks of traveling internationally.

**Doctor:** I hear your concern about seeing your father and understand this must be weighing heavily on you. We'll definitely address the travel considerations. Before we delve deeper into that, I notice you're on several medications. Could you tell me what you take the tacrolimus for?
**Patient:** Lung transplant.

**Doctor:** Thank you. And the mycophenolate mofetil - is this also for your lung transplant?
**Patient:** Yes.

**Doctor:** I understand. Let's focus first on your lung transplant, which seems to be one of your major medical conditions. Could you tell me when you received your lung transplant and which hospital/team manages this?
**Patient:** Five years ago. Specialist CF team (pulmonologist, dietitian, endocrinologist, physiotherapist).

**Doctor:** Thank you for that information. Regarding your lung transplant, do you know the characteristics of your donor, particularly their CMV (cytomegalovirus) and EBV (Epstein-Barr virus) status?
**Patient:** Donor characteristics: CMV and EBV positive; recipient CMV mismatch, hence on prophylaxis (famciclovir).

**Doctor:** How is your current lung function? Could you tell me about any shortness of breath you experience and your exercise tolerance?
**Patient:** Worse recently. I stop after walking ~100 meters due to shortness of breath.

**Doctor:** This decline in exercise tolerance is significant and warrants attention. Have you noticed any other symptoms besides the shortness of breath when walking 100 meters? For example, cough, chest pain, or fever?
**Patient:** Occasional dry cough; sputum minimal and typically clear.

**Doctor:** I understand. Let's discuss your immunosuppression regimen. You're currently on tacrolimus, mycophenolate mofetil, and prednisolone. Have you experienced any side effects from these medications, such as tremors, stomach upset, or increased blood sugars?
**Patient:** No major side effects. Sugars managed with insulin.

**Doctor:** I understand. Have you had any episodes of rejection since your transplant, either in the first year (acute rejection) or more recently (chronic rejection)? I notice this may be especially relevant given your recent decline in exercise tolerance.
**Patient:** The team has mentioned the possibility of chronic rejection.

**Doctor:** Given this concern about possible chronic rejection, I'd like to understand more about your monitoring. When was your most recent lung function test (spirometry/FEV1), and how often are you being followed up by your transplant team?
**Patient:** FEV₁: Post-transplant baseline ~65% predicted; recent decline to 50%. This was most recently around 1 month ago.

**Doctor:** I see. This decline in your FEV1 from 65% to 50% is significant. Regarding infections, which can be especially important in transplant patients, have you had any recent lung infections or been prescribed any new antibiotics?
**Patient:** Hospitalization 2 months ago with a severe respiratory infection.

**Doctor:** Given your recent hospitalization and respiratory infection, which has been followed by this decline in lung function, I'd like to understand your current treatments. Besides your immunosuppression medications, are you on any specific antibiotics or other treatments for your respiratory health?
**Patient:** I had antibiotics at that time. Now I'm just on my usual meds, including azithromycin, trimethoprim-sulfamethoxazole, and famciclovir.

**Doctor:** Let's discuss your cystic fibrosis, which was your underlying condition before the transplant. When were you first diagnosed with CF, and do you recall if genetic testing was done?
**Patient:** Diagnosed in early childhood (age 2) following sweat chloride testing; genetic testing identified homozygous ΔF508 mutation in the CFTR gene.

**Doctor:** Thank you for that information about your CF diagnosis. With CF affecting multiple organ systems, I notice you're on pancreatic enzyme replacement (pancrelipase). Could you tell me about any digestive symptoms you experience, such as abdominal pain or issues with bowel movements?
**Patient:** No major GI issues, but I struggle to maintain weight.

**Doctor:** Given your challenge with maintaining weight, could you tell me your current weight and height? Also, how has your weight changed over the past 6-12 months?
**Patient:**

- Height: ~180 cm
- Weight: ~58 kg
- BMI: ~17.9–18.0 (underweight category).
  Fluctuates. 1 year ago BMI was around 19.

**Doctor:** I understand this weight loss is concerning. Related to both CF and nutrition, I see you're on fat-soluble vitamin supplements. When was your most recent vitamin level check, and have there been any deficiencies?
**Patient:** They've been okay, checked around 1 month ago.

**Doctor:** Regarding your history of intravenous drug use that you mentioned earlier, could you tell me when this occurred and whether you are currently using any recreational substances?
**Patient:** Intravenous methamphetamines. Long term, intermittent, most recently 3 weeks ago.

**Doctor:** I understand. Given your recent substance use, I want to ask about your support system. Who do you live with at home, and do you have family or friends nearby who help support you?
**Patient:** Live alone in a small apartment (single-level). One close friend. No immediate family nearby. Father resides in Spain and has been diagnosed with terminal pancreatic cancer.

**Doctor:** Thank you for sharing that about your living situation and your father - this helps explain your earlier concern about travel. Regarding your diabetes, which you mentioned is related to your CF, when was this diagnosed and what was your most recent HbA1c?
**Patient:** Diagnosed 8 years ago. Don't remember the most recent HbA1c, sorry.

**Doctor:** How are you managing with your insulin regimen? Do you experience any episodes of low blood sugar (hypoglycemia)?
**Patient:** Basal-bolus. No major hypos. Tends to run a bit high, like BGLs in the low teens.

**Doctor:** Thank you for sharing that information about your blood sugar management. Given all of your medical conditions and medications, I'd like to ask about your medication routine. Are you able to take all your medications as prescribed, or do you sometimes have difficulty remembering or affording them?
**Patient:** Yes, I take all of them as prescribed. I know it is important. But the financial strain is real.

**Doctor:** Thank you for being candid about the financial challenges. Given your upcoming need to travel internationally to see your father, could you tell me more about your current financial situation - specifically regarding insurance coverage and ability to afford travel?
**Patient:** Used to work full-time as a computer programmer but have reduced hours significantly due to health issues, resulting in financial difficulties.

**Doctor:** I understand the challenges you're facing with reduced work hours. Considering your complex medical situation and desire to travel, I'd like to ask about your advance care planning. Do you have an advance care directive or have you designated someone as your medical power of attorney?
**Patient:** Advanced Care Directive/EPOA/SDM: None in place; no local family members to assist with decision-making.

**Doctor:** Given your complex medical situation and current challenges, I'd like to ask about your mental health and emotional wellbeing. How are you coping with everything, particularly with the concerns about your father?
**Patient:** Experience anxiety and low mood but no formal mental health diagnosis.

**Doctor:** I appreciate you sharing that about your mental health. Given everything we've discussed - your transplant management, CF care, substance use, and the emotional and financial challenges around seeing your father - would you say there are any other major concerns or anything else important I should know?
**Patient:** About the CF, I think I'm getting resistant bugs now. And I get some joint pains.

**Doctor:** Thank you for sharing that additional information. Let's discuss your joints and respiratory infections first. Could you tell me more specifically about where you experience joint pain and whether you've noticed any particular patterns or triggers?
**Patient:** Just all over, particularly my knees.

**Doctor:** Thank you for that clarification about your joint pain. Considering your upcoming international travel plans, when was your last vaccination for flu and pneumococcus? Also, are you up to date with your COVID-19 vaccinations?
**Patient:** Up to date with COVID, flu, and pneumococcal vaccines.

**Doctor:** Regarding your concern about resistant organisms - could you tell me more about your recent respiratory infections, including what antibiotics you've been treated with and any organisms that have been identified in your sputum cultures?
**Patient:** In that hospital admission two months ago, I hadn't had one for over a year before that. Sputum culture grew multiresistant Pseudomonas.

**Doctor:** Thank you for that information about the Pseudomonas infection. Given your complex medical situation - including your lung transplant, declining lung function, recent infection with resistant organisms, and your strong desire to see your father - I'd like to ask about your exercise and physiotherapy routine. What type of exercises are you currently able to do, and are you working with a physiotherapist?
**Patient:** Fatigue and shortness of breath are stopping me from being very physically active. And not enjoying my computer games as much anymore either.

**Doctor:** I understand your frustration with the impact fatigue and breathlessness are having on both your physical activity and your ability to enjoy your usual hobbies. Could you tell me about your typical sleep - are you able to get restful sleep at night?
**Patient:** I sleep okay. 8 hours a night or more.

**Doctor:** Thank you for sharing that about your sleep patterns. Given your history of substance use and living alone, I'd like to make sure you're safe at home. Do you have any concerns about safety in your current living situation, or about accessing emergency help if needed?
**Patient:** I feel safe at home. And can use my mobile.

**Doctor:** Thank you for providing those details about your emergency access. Given the complexity of your medical conditions and the importance of your goal to see your father, I'd like to ask about any plans for travel that you've already started making. Have you discussed the possibility of international travel with your transplant team?
**Patient:** I called them about it a week ago when I found out more about my dad's prognosis getting worse. Now I have an appointment with them really soon.

**Doctor:** I understand you have an upcoming appointment with your transplant team to discuss travel. One key aspect of travel planning would be your medication supply and schedule. Have you thought about how you would manage your medications, particularly your insulin and immunosuppressants, across different time zones?
**Patient:** I hadn't really thought about that. Could be a challenge. Maybe use a notebook or phone?

**Doctor:** Given your current medications, planned travel, and the recent updates about your father's condition, do you have access to any social work support or counseling services through your transplant team?
**Patient:** I think I could access them. I haven't before. Probably worth trying.

**Doctor:** Considering everything we've discussed today, I'd like to focus on your preventive health. How often do you see your dentist? This is particularly important given your transplant status and potential travel plans.
**Patient:** My teeth are okay. Saw a dentist 1 year ago.

**Doctor:** Thanks for sharing that about your dental care. Given your transplant and diabetes status, have you had any eye examinations in the past year for screening?
**Patient:** No, I haven't had my eyes checked. But I think they're okay.

**Doctor:** Given that you haven't had recent eye screening and you have both diabetes and complex medical needs, when was your last comprehensive medical check-up addressing preventive health measures beyond your transplant care?
**Patient:** Not sure. My transplant team manages everything.

**Doctor:** Given your frequent contact with your transplant team but complex medical needs, do you know if there have been any recent bone density (DEXA) scans, particularly given your long-term steroid use and CF history?
**Patient:** Not sure about the number, but I know I have osteopenia.

**Doctor:** Thank you for sharing that about your bone health. Regarding your current substance use, are you currently receiving or interested in any support services or treatment programs?
**Patient:** I'm not. But I don't think it is my biggest issue at the moment. I think my dad and lungs and finances are.

**Doctor:** Thank you for sharing that perspective about your priorities. I can hear that your father's illness, your lung function, and financial challenges are paramount concerns right now. Could you tell me more about any specific goals or wishes you have regarding seeing your father, beyond just making the trip? For instance, are there particular things you hope to do or discuss with him?
**Patient:** I just want to see him before he dies.

**Doctor:** I hear how important it is for you to see your father, and I understand the urgency of the situation. Given all we've discussed today, I have significant concerns about several aspects of your health that we need to address promptly:

1. Your lung function has declined notably, with your FEV1 dropping from 65% to 50%, alongside a concerning recent hospitalization for a resistant Pseudomonas infection. This decline, combined with your increased shortness of breath with minimal exertion, suggests possible chronic rejection that requires immediate attention.
2. Your nutritional status is concerning, with a BMI of 17.9-18.0 and recent weight loss from a BMI of 19 one year ago. This decline in nutritional status could impact your overall health and ability to travel.
3. The combination of your complex medical needs (including the need for regular insulin administration), recent substance use, and your strong desire to travel internationally to see your father creates a challenging situation that requires careful planning and support.

Would you like me to explain more about any of these concerns, particularly as they relate to your goal of seeing your father?
**Patient:** When you put it like that, it is all very concerning. I think we're just about out of time. Was there anything else you wanted to ask me?

**Doctor:** Yes, just one final but important question before we end our session. Given that you live alone and mentioned having one close friend, do you have a reliable system in place for monitoring and communicating any sudden changes in your health status, particularly with your transplant team? For instance, do you have a way to regularly check and record your vital signs at home?
**Patient:** No, I don't, but I think I know my body okay.

#### Case 2 – Heart Failure with reduced Ejection Fraction and falls

**Doctor:** Good morning, Mrs. Ng. I'm Dr. Claude, an internal medicine specialist. I'll be conducting a comprehensive medical assessment today. I may need to ask quite a few questions to ensure I understand your health situation fully. Please let me know if you need me to clarify anything or if you'd like to take a break at any point.
**Patient:** Sure.

**Doctor:** Could you please tell me what medical conditions you are currently aware of having?
**Patient:**

- HFREF
- Ischaemic heart disease with NSTEMIs
- Atrial fibrillation
- COPD
- I fall occasionally

**Doctor:** Thank you for sharing that information. Could you tell me what your main concern is currently regarding your health?
**Patient:** I don’t want to leave my home or keep going to the hospital. I’d rather just stay here in my garden and be left alone.

**Doctor:** I hear your concern about wanting to stay at home and avoid hospitalizations. That must be challenging. Let me help better understand your heart condition first, as that seems to be a major health issue. When were you first diagnosed with heart failure, and who manages this condition for you?
**Patient:** First diagnosed 2 years ago after presenting to my GP with shortness of breath and undergoing a transthoracic echocardiogram (TTE). Echocardiogram revealed EF of ~20%; clinical presentation with exertional dyspnea.

**Doctor:** Thank you for that information. Could you tell me about your current symptoms - specifically how far can you walk before becoming short of breath? And have you noticed any swelling in your legs or weight changes recently?
**Patient:** Short of breath after 10 meters. Some leg swelling. Not sure about weight.

**Doctor:** Thank you for sharing that. I understand this is significantly limiting your daily activities. Regarding your heart medications, I notice you're taking Entresto, furosemide, bisoprolol, and spironolactone. Are you managing to take all of these medications as prescribed, or do you have any difficulties with them?
**Patient:** Yes, I take these medications as prescribed. No major side effects.

**Doctor:** I understand. Now, regarding your atrial fibrillation, I see you're taking apixaban for this. When were you first diagnosed with atrial fibrillation, and have you had any complications from it?
**Patient:** Identified during my most recent hospital admission (~2 weeks ago) on telemetry/ECG.

**Doctor:** I see this was quite recent. Given your recent hospitalization and your wish to stay at home, could you tell me about any other hospital admissions you've had in the past year related to your heart condition?
**Patient:** Three hospital admissions in the last 4 months, responding to higher diuretic doses.

**Doctor:** Thank you for sharing that information about your hospitalizations. I notice you also mentioned having COPD, and I see you're using a salbutamol inhaler. Could you tell me how often you need to use your inhaler in a typical week?
**Patient:** Salbutamol inhaler PRN 3-4 times per week.

**Doctor:** I understand. Now, regarding your falls that you mentioned earlier - when was your most recent fall, and could you tell me what happened?
**Patient:** Occurred 2 weeks ago while gardening. No dizziness. Think I tripped over my cat.

**Doctor:** Given you live alone and enjoy gardening, I'm concerned about your safety. Could you tell me about your home situation - do you have any family members or friends who live nearby who help support you?
**Patient:** No family; few friends. Limited social support network. My husband died 4 years ago. Still enjoy my gardening though!

**Doctor:** I'm sorry to hear about your husband. It's wonderful that you still find joy in gardening. Could you tell me what support services, if any, you currently have to help with daily activities like cleaning, cooking, or shopping?
**Patient:** I like to stay as independent as possible. If I had to, to stay in my home, then okay.

**Doctor:** I understand your desire for independence. Given your heart condition and recent falls, I'd like to ask about your mobility. Do you use any walking aids like a walker or cane to help you move around safely?
**Patient:** I was advised to use a 4-wheeled walker (4WW), but I prefer not to as I worry it is the first step of being forced out of my home.

**Doctor:** I understand your concern about the walker representing a loss of independence. Let me ask about your daily routine - how do you currently manage tasks like preparing meals and taking care of yourself at home?
**Patient:** I live alone, fiercely independent, in my own house. I do it all for myself.

**Doctor:** Thank you for sharing that. Given your medical conditions and desire to stay independent, I'd like to understand more about your preventive health. Are you up to date with your vaccinations, particularly for flu and pneumonia?
**Patient:** Up to date with COVID, influenza, and pneumococcal vaccines.

**Doctor:** Could you tell me your current height and weight? This will help me understand your overall health status better.
**Patient:**

1. Height: ~160 cm
2. Weight: ~52 kg
3. BMI: ~20 (low-normal)
   Recent 5 kg weight loss, not sure why.

**Doctor:** That's concerning about your recent weight loss. Could you tell me more about your typical daily diet - what kinds of foods do you usually eat and how many meals do you have per day?
**Patient:** Tea and toast predominantly. 1.25L fluid restriction.

**Doctor:** Given your limited diet and recent weight loss, I'd like to ask about any changes in your appetite recently. Have you noticed feeling less hungry than usual or having any difficulties eating?
**Patient:** Maybe a bit less hungry, but still eating my normal amount.

**Doctor:** I understand. I notice you mentioned living alone in your house - could you tell me about the layout? Specifically, are there any stairs you need to navigate?
**Patient:** No stairs. Pretty well illuminated.

**Doctor:** Thank you for that information. Could you tell me if you consume any alcohol or if you have a history of smoking?
**Patient:** Currently an active smoker (~10 cigarettes/day). No alcohol use.

**Doctor:** I see. Regarding your heart condition and overall health, have you ever been referred to a Cardiac Rehabilitation program or received advice about exercise and smoking cessation?
**Patient:** I haven’t been to rehab. Could consider it. But really, I just want to spend time in my garden and avoid medical things as much as possible. As for smoking, I’ve been smoking for 50 years, and I don’t plan on quitting now.

**Doctor:** Let me ask about advance care planning. Have you thought about or documented your wishes regarding future medical care, such as having an Advance Care Directive or appointing someone to make medical decisions for you if needed?
**Patient:** No Advance Care Directive or will.

**Doctor:** Thank you for sharing that. I'd like to ask about your pregabalin prescription - could you tell me what symptoms or condition this was prescribed for?
**Patient:** No idea.

**Doctor:** I notice you mentioned you don't want "medical things" and want to stay independent in your garden, which is completely understandable. Before we continue with more questions, could you share with me what worries you most about doctors and hospitals?
**Patient:** I don't like hospitals because they make me feel constrained. I've always highly valued my independence. I don’t ever want to feel dependent. When I go into medical things, I always worry they’ll try and take my freedom away.

**Doctor:** I really appreciate you sharing that with me. Your desire for independence and your love of gardening come through very clearly. Let me ask about your feelings around having some support at home - if it meant you could stay in your home and garden longer, would you consider accepting some help with tasks like cleaning or meal preparation?
**Patient:** Yes, I’d prefer to avoid it, but if it means that I can stay at home and not go to a nursing home, then I’d begrudgingly accept it.

**Doctor:** I understand that independence means a great deal to you. Could you tell me about any activities you find more difficult to do now compared to 6 months ago?
**Patient:** Increased difficulty maintaining my home and garden.

**Doctor:** Thank you for sharing that. Given your heart failure and difficulty with activities, I'd like to understand more about your typical day. Could you walk me through what a usual morning looks like for you - from when you wake up until lunchtime?
**Patient:** Wake up. Toast for breakfast. Go into the garden. Sometimes just sit in the garden if I’m tired. If I’m feeling better, do some light pruning. Then in for lunch, often a cheese sandwich.

**Doctor:** I understand this is your daily routine. Given you mentioned your shortness of breath after walking 10 meters, how do you pace yourself when working in the garden to manage your symptoms?
**Patient:** Frequent rests.

**Doctor:** Thank you for explaining that. Since you mentioned having atrial fibrillation and taking apixaban, do you ever check your pulse or notice if your heart feels like it's racing or beating irregularly?
**Patient:** No, I don’t. Never knew I had it, really. They just happened to find it at my most recent hospital admission 2 weeks ago.

**Doctor:** I understand. Do you have a thermometer at home to check your temperature if you feel unwell, or any other monitoring equipment like a blood pressure machine or weight scale that you use?
**Patient:** No to thermometer and blood pressure machine. I do have scales but probably don’t use them as often as I should, only when I’m feeling like I’m retaining fluid.

**Doctor:** Regarding your breathing problems, do you notice if your breathing is worse at any particular time of day, or when you're doing certain activities besides walking?
**Patient:** Mainly on exertion. Sometimes worse when I lie down.

**Doctor:** Thank you for sharing that. Since staying healthy at home is important to you, I'd like to ask about who you would call first if you started feeling unwell or having worse breathing problems than usual?
**Patient:** No one probably. But an ambulance if bad enough.

**Doctor:** Since managing your heart failure well is key to staying at home, could you tell me if you monitor or record your daily fluid intake to stay within your 1.25L restriction?
**Patient:** I stick to the fluid restriction pretty well.

**Doctor:** Understanding you keep track of your fluids well, I'd like to ask about your feet and legs. Have you noticed any changes in sensation, numbness, or tingling in your feet?
**Patient:** No.

**Doctor:** I notice you were recently hospitalized for heart failure. Do you monitor your breathing when lying flat at night, and if so, how many pillows do you need to use to sleep comfortably?
**Patient:** 2-3 pillows most nights.

**Doctor:** Given your recent hospitalizations and that you live alone, do you have a community nurse or GP who visits you at home to check on you regularly?
**Patient:** I have a GP, but the heart failure is mainly managed by the hospital cardiology team.

**Doctor:** Thank you for this detailed information. Since your heart team manages most of your cardiac care, when is your next scheduled follow-up appointment with them?
**Patient:** In the next couple of weeks, in view of the recent admission.

**Doctor:** Based on your difficulty with exertion and recent weight loss, how often do you feel short of breath when doing daily activities like showering or getting dressed?
**Patient:** Infrequent right now because I pace myself. But when leading up to hospital admission, I end up short of breath on minimal movement or at rest.

**Doctor:** Since maintaining your independence at home is important, could you tell me how you manage your medications - do you use any system like a webster pack or pill organizer to help keep track of them?
**Patient:** I use a dosette pill organizer.

**Doctor:** Given your COPD and smoking history, could you tell me if you notice changes in your breathing during different weather conditions or seasons?
**Patient:** A bit, but more related to my fluid status.

**Doctor:** I see. Going back to your medications for a moment - you take aspirin and atorvastatin. Have you had any heart attacks or stents placed in the past?
**Patient:** Yes, I’ve had two heart attacks. Got drug-eluting stents for both.

**Doctor:** Thank you for sharing that information about your cardiac history. Since staying independent is important to you, do you have any issues with your vision that might affect your daily activities or safety at home?
**Patient:** My vision is okay.

**Doctor:** Given that falls are a concern, could you tell me if you've noticed any dizziness when changing positions, like standing up from sitting or lying down?
**Patient:** No dizziness.

**Doctor:** Since you mentioned living alone, do you have any neighbors nearby that you're in regular contact with or could call on if needed?
**Patient:** Not really. Wave at them sometimes. But not close. Have some friends though, no family.

**Doctor:** I understand you have few close contacts. Let me ask about the medications again - when you use your salbutamol inhaler, do you find it helps improve your breathing symptoms?
**Patient:** Slightly? Not much really.

**Doctor:** Given that you mentioned the salbutamol doesn’t help much, does your breathing pattern feel different when you're short of breath due to your heart versus your lung condition?
**Patient:** Difficult to say. Think it is mainly the fluid.

**Doctor:** Based on what you've shared about your breathing difficulties being mainly related to fluid, how often do you feel you need to rest during gardening activities compared to a few months ago?
**Patient:** More often now. I do think things are getting harder.

**Doctor:** I understand you're finding activities more challenging. Could you tell me what changes or adaptations you've already made to your gardening routine to help manage your symptoms?
**Patient:** More frequent rests. More time just sitting. I think we’re close to running out of time. Are there any final particularly important questions you’d like to get through?

**Doctor:** Yes, given your recent hospitalizations and desire to stay independent, I have two important final questions. First, do you know what symptoms or changes you should watch for that would indicate you need to seek medical help for your heart condition?
**Patient:** Yes, worsening shortness of breath or increased ankle swelling.

**Doctor:** For my final question - since you mentioned you’d consider accepting some help to stay independent at home, what specific types of support would you be most comfortable starting with?
**Patient:** Maybe cleaning. I get quite short of breath. At some point, someone mentioned palliative care, but I don’t know much about what that is.

**Doctor:** Thank you for mentioning palliative care. Given everything you've shared - your advanced heart failure, desire to stay independent at home, focus on quality of life, and wish to avoid hospitalizations - would you like me to explain what palliative care could offer in supporting these goals?
**Patient:** Sure.

**Doctor:** Palliative care is quite different from what many people initially think. It's a specialized medical service that focuses on helping people with serious illnesses live as well as possible, while honoring their values and wishes - which in your case clearly center around independence and staying in your home and garden.
The palliative care team could help in several important ways: they can provide expertise in managing symptoms like shortness of breath and fatigue that affect your daily activities, coordinate your care to minimize hospital visits, and support you in maintaining independence at home. They work alongside your existing heart team, adding an extra layer of support focused specifically on your quality of life.
Importantly, palliative care is not about giving up - rather, it’s about living as fully as possible according to what matters most to you. They could help arrange the specific home supports you're comfortable with, like cleaning assistance, while respecting your independence. They're also experts at helping people stay out of the hospital when possible, which aligns with your wishes.
Would you like to learn more about how palliative care might be able to help you specifically with your goals?
**Patient:** Yeah, I’d be happy to learn more later on. Out of time, thanks, and good luck for your exam.

**Doctor:** Thank you for sharing your story with me today, Mrs. Ng. I really appreciate how open you’ve been about your health conditions and your strong desire to maintain independence at home. Your love of gardening and commitment to staying in your own space came through very clearly.
Best wishes for your ongoing care and for connecting with the palliative care team to learn more about how they might support your goals.

#### Case 3 – Myasthenia gravis and osteoporosis

**Doctor:** Good morning, Mrs. Brown. I'm Dr. Claude, an internal medicine specialist. Thank you for coming in today. I'll be conducting a comprehensive medical evaluation, which may take around 30 minutes. I may need to politely interrupt at times to ensure we cover everything important. Is that alright with you?
**Patient:** Yes, that’s fine.

**Doctor:** Would you please list all of the medical conditions that you are currently aware of having, in a brief manner?
**Patient:**

- Myasthenia gravis
- Osteoporosis
- Falls
- Depression
- Obesity
- GORD
- Asthma

**Doctor:** Thank you, Mrs. Brown. Before we discuss each condition in detail, could you tell me what is your main concern currently regarding your health?
**Patient:** I’m worried about how much longer I’ll be able to manage my day-to-day activities. The pain is unbearable, and I feel like I’m losing my independence and my connection with the world.

**Doctor:** I hear how deeply this is affecting you, and I want to better understand the pain you're experiencing. Could you tell me more about this pain - where is it located and how long have you been experiencing it?
**Patient:** Middle to lower back, for 2 years, but worse recently.

**Doctor:** I understand. Let me go through your medication list to ensure I understand what each one is being used for. You're taking prednisolone and mycophenolate - are these for your myasthenia gravis?
**Patient:** Yes.

**Doctor:** I see you're also taking calcium, vitamin D, and denosumab. Are these prescribed for your osteoporosis?
**Patient:** Yes.

**Doctor:** Thank you. I notice you're taking duloxetine, paracetamol, ibuprofen, and tramadol - are all of these medications for your back pain that you mentioned?
**Patient:** Yes, but the duloxetine is also for my depression.

**Doctor:** I understand. You're taking esomeprazole - is this for your gastroesophageal reflux disease (GORD)?
**Patient:** Yes.

**Doctor:** And I see you're taking trimethoprim-sulfamethoxazole - is this prescribed as an infection preventative medication due to your immunosuppression with prednisolone and mycophenolate?
**Patient:** Yes.

**Doctor:** I see you're also prescribed a salbutamol inhaler as needed for your asthma. How often do you need to use this inhaler in a typical week?
**Patient:** Less than once per month.

**Doctor:** I'd like to learn more about your myasthenia gravis, as it's a significant condition. When were you diagnosed with myasthenia gravis and what symptoms led to the diagnosis?
**Patient:** Diagnosed 5 months ago. AChR antibody positivity and clinical presentation of neck weakness and swallowing difficulties.

**Doctor:** Thank you. Who manages your myasthenia gravis care, and how often do you see them for follow-up?
**Patient:** Managed by a neurologist. Every 1-2 months at the moment.

**Doctor:** Are you currently experiencing any weakness symptoms, particularly with your swallowing or neck muscles?
**Patient:** Symptoms include intermittent fatigue, lower limb weakness, and swallowing difficulties. Worse when I try reducing prednisolone.

**Doctor:** Have you ever needed hospitalization or intensive care due to your myasthenia gravis symptoms, particularly any breathing difficulties?
**Patient:** I had a hospitalization at the time of diagnosis. Initial presentation required IVIg therapy. No major breathing symptoms but had swallowing problems.

**Doctor:** Thank you for sharing that information. With regard to your osteoporosis, when was this diagnosed and what led to the diagnosis?
**Patient:** Diagnosed 2 years ago after new onset back pain and a lumbar vertebral fragility fracture confirmed on X-ray.

**Doctor:** I understand. Let's focus on your recent back pain. You mentioned it's getting worse - what activities or movements make the pain better or worse?
**Patient:** Pain is worse with sitting or standing for long periods and disrupts sleep.

**Doctor:** In regards to your falls, you mentioned this as one of your conditions. When was your most recent fall, and could you tell me what happened?
**Patient:** Most recently three weeks ago. Felt like my legs were weak and gave way.

**Doctor:** Could you tell me about your daily routine at home? Who do you live with, and do you have any support for daily activities like cooking and cleaning?
**Patient:** Live at home with my husband in a single-story house (no stairs). Primarily supported by my husband for cooking, cleaning, and shopping. No formal supports currently in place.

**Doctor:** How do you currently manage with your mobility? Do you use any walking aids?
**Patient:** No gait aids required. Pain from vertebral fracture significantly limits walking and standing.

**Doctor:** I understand you mentioned depression as one of your conditions. When were you first diagnosed with depression, and who manages this care?
**Patient:** Diagnosed 15 years ago based on clinical history. Managed by my general practitioner, with occasional input from a psychologist.

**Doctor:** With all your medications and health conditions, it's important to monitor preventive health measures. When was your last bone density scan (DEXA)?
**Patient:** DEXA scan 18 months ago showing T-score -2.8 at the lumbar spine.

**Doctor:** Could you tell me about your current exercise routine or physical activities, and how the back pain affects your ability to stay active?
**Patient:** Previously enjoyed daily 2 km walks. Now primarily sedentary.

**Doctor:** In terms of your mobility limitations, have you ever seen a physiotherapist or occupational therapist for assessment or management of your back pain and weakness?
**Patient:** Maybe a long time ago, nothing recently.

**Doctor:** With your myasthenia gravis and back pain affecting your daily activities, what goals would you like to achieve in terms of your function and independence?
**Patient:**

- Would like to visit my sister interstate but feel uncertain about managing fatigue during travel.
- Wish to regain strength and independence to return to gardening and light exercise.

**Doctor:** You've mentioned visiting your sister - that sounds like an important goal. Could you tell me about your current support network - do you have family or friends nearby who help you?
**Patient:** Live at home with my husband in a single-story house (no stairs). Primarily supported by my husband for cooking, cleaning, and shopping. No formal supports currently in place.

**Doctor:** Given your complex medical conditions, I'd like to understand your current weight, height, and any recent changes. Could you share these with me?
**Patient:**

- Height: 165 cm
- Weight: 89 kg
- BMI: 32.7 kg/m² (Obese category)
- Waist Circumference: 105 cm
  No major recent changes, perhaps slight weight gain.

**Doctor:** Before we discuss those measurements, I'd like to ask about any smoking history or alcohol consumption - could you tell me about these?
**Patient:** Nil smoking or alcohol.

**Doctor:** Thanks for sharing that. Given that you're taking several medications that can affect bone health, could you tell me about your typical daily calcium intake through diet (for example, dairy products)?
**Patient:** High-calorie diet, frequent takeaway meals, low intake of fresh fruits and vegetables. Some dairy.

**Doctor:** Given your complex medical conditions and goals, I'd like to understand if you've had any recent blood tests done, particularly monitoring your inflammatory markers, kidney function, or bone health markers?
**Patient:** My neurologist did some blood tests maybe 3 weeks ago, but I'm not sure of the results.

**Doctor:** Have you received any vaccinations recently, particularly for flu or pneumonia, given your medical conditions and immunosuppression?
**Patient:** Up to date on influenza, pneumococcal, and COVID vaccines.

**Doctor:** Thank you for sharing all of that information. Could you tell me about any advance care planning discussions you've had with your healthcare providers or family?
**Patient:** My husband is my informal substitute decision-maker. No formal advanced care directive in place.

**Doctor:** Based on our comprehensive discussion, Mrs. Brown, I'd like to confirm I understand your main concerns and care priorities. The significant impact of back pain on your daily activities, combined with myasthenia gravis symptoms, is affecting your independence and quality of life. You have clear goals around visiting your sister and returning to gardening, which we should incorporate into your care plan.
I believe it would be beneficial to focus on several key areas:

1. The management of your back pain, which appears to be significantly impacting your daily function and mood.
2. Your myasthenia gravis symptoms, particularly the intermittent fatigue and weakness.
3. Your physical function and mobility, given your recent falls.
4. Your mental health and social connection.

Would you like me to explain my recommendations for addressing these areas? Also, is there anything else you'd like to discuss that we haven't covered?
**Patient:** No longer sexually active due to pain and fatigue.

**Doctor:** Thank you for sharing that personal information. I understand this is a sensitive topic that impacts your quality of life. The combination of pain, fatigue, and medical conditions can significantly affect intimate relationships. Would you like to discuss this aspect of your health further with me, or would you prefer a referral to discuss these concerns with a specialist?
**Patient:** I don’t need to discuss further, just thought I’d mention it.

**Doctor:** Since we've covered a lot of ground, I'd like to address the key elements of your care that I believe need attention. Your conditions - myasthenia gravis, osteoporosis, chronic pain, depression, and recent falls - are creating significant challenges for your daily life and independence. The impact is particularly evident in your reduced mobility, social engagement, and ability to enjoy activities like gardening.
I recommend a comprehensive care approach that includes:

1. **Optimizing your pain management** through a review of your current medications and their effectiveness. This would involve coordinating with your GP and neurologist to ensure your pain control balances with your myasthenia gravis management.
2. **Implementing a supervised exercise and rehabilitation program.** A physiotherapist could help develop a safe exercise plan that considers both your back pain and muscle weakness, potentially helping prevent future falls.
3. **Exploring additional support services** to help maintain your independence at home and facilitate your goal of interstate travel. This might include occupational therapy assessment and connection with community support services.

Would you like me to explain any of these recommendations in more detail, or do you have any questions about what I've suggested?
**Patient:** No, that’s okay. Just before we finish up, was there anything else that you needed to ask me?

**Doctor:** Yes, since you're taking immunosuppressive medications, could you tell me about any recent infections, fevers, or new symptoms we haven't discussed?
**Patient:** No, I haven’t.

**Doctor:** Have you noticed any side effects from your various medications that we haven't already discussed, particularly any tremors, stomach upset, or skin changes?
**Patient:** No.

**Doctor:** Thank you for taking the time to share your medical history with me today, Mrs. Brown. I've developed a thorough understanding of your health situation, particularly focusing on your myasthenia gravis management, chronic pain, and how these conditions are impacting your daily activities and independence.
I’ll be preparing a plan based on what we’ve discussed today and ensuring we focus on your goals of improving mobility, managing pain, and maintaining independence. Thank you for your openness, and I wish you the best with your care journey.

#### Case 4 – Perioperative and rural

**Doctor:** Hello, Mr. Wilson. I'm Dr. Claude. Thank you for coming in today. I'll be conducting a comprehensive medical evaluation, which will take about 30 minutes. I may need to politely interrupt at times to ensure we cover everything important. Is that alright with you?
**Patient:** Yes, that’s fine.

**Doctor:** Could you please list all of the medical conditions that you are currently aware of having, in a very brief fashion?
**Patient:**

- Abdominal aortic aneurysm
- Cirrhosis
- COPD
- Type 2 diabetes mellitus
- Chronic kidney disease

**Doctor:** Thank you for sharing those conditions. Could you tell me what is your main concern currently regarding your health?
**Patient:** I’m scared I’ll end up like my father, who never came home from his heart operation. I don’t want to go to the city, but they say I need this surgery. I just want my family there to help me decide.

**Doctor:** I'm very sorry to hear about your father, and I understand your concerns about the surgery. That must be difficult to process. Let me first go through your medication list to ensure I understand all your conditions clearly.
I see you're taking metformin and gliclazide - are these for your Type 2 diabetes?
**Patient:** Yes.

**Doctor:** I see you're also taking ramipril and atorvastatin. Are these to help protect your heart and blood vessels?
**Patient:** Yes, and maybe for hypertension too, I’m not sure. Ramipril might also be for my kidneys.

**Doctor:** I understand - yes, ramipril can help protect both your heart and kidneys. I see you're also using a salbutamol inhaler. Is this for breathing difficulties related to your COPD?
**Patient:** Yes.

**Doctor:** I notice you're also taking thiamine. Was this prescribed for your liver condition - the cirrhosis?
**Patient:** I think so.

**Doctor:** Thank you for clarifying your medications. Now, regarding your cirrhosis, could you tell me what caused this condition and when you were diagnosed?
**Patient:** Diagnosed about 5 years ago during evaluation for abnormal liver function tests (LFTs). The cause was long-term excessive alcohol consumption, but I've cut that almost completely out now.

**Doctor:** Thank you for sharing that. I'd like to learn more about your current drinking habits. Could you tell me specifically how many standard drinks you have in a typical week now?
**Patient:** I don't drink alcohol except on special occasions, maybe 3-4 times per year.

**Doctor:** Thank you for sharing that information about your alcohol consumption. Regarding your abdominal aortic aneurysm, when was this diagnosed and what size was it at your last scan?
**Patient:** First detected on an abdominal ultrasound approximately 18 months ago. Most recent ultrasound 2 weeks ago shows an infrarenal AAA measuring 6.5 cm, having grown 1 cm over the last 6 months.

**Doctor:** I understand this growth in your aneurysm is likely what's prompting the discussion about surgery. Regarding your chronic kidney disease, could you tell me your most recent kidney function test results and when they were done?
**Patient:** Around 30 to 60, something in that range. Was performed a couple of weeks ago.

**Doctor:** Thank you for sharing that information about your kidney function. Regarding your Type 2 diabetes, when was this first diagnosed and what was your most recent HbA1c reading?
**Patient:** At age 44, after I went to the doctor because I was peeing a lot and had high sugar levels. Most recent HbA1c was 7.9% a few weeks ago.

**Doctor:** Regarding your COPD, could you tell me about any breathing difficulties you experience - for example, how often do you need to use your salbutamol inhaler in a typical week?
**Patient:** I use the reliever maybe once per week.

**Doctor:** Thank you for that information about your breathing. Could you tell me about your living situation - do you live alone or with others, and what type of home do you live in?
**Patient:** I live in Murray Bridge in Australia. I have a big family, including my three brothers and two sisters who live nearby, and we're frequently at each other’s houses.

**Doctor:** I'm glad to hear you have such strong family support nearby. Could you tell me about your mobility - do you use any walking aids, and how far can you walk comfortably?
**Patient:** I’m independent with daily activities and I drive. But I’m very reluctant to travel to the city (Adelaide) for medical care. As an Aboriginal man, I feel more comfortable closer to my home in Murray Bridge.

**Doctor:** Thank you for sharing that important cultural context. I understand your preference to stay near home and family. Could you tell me about any allergies you have to medications?
**Patient:** No allergies.

**Doctor:** Thank you for sharing that information about allergies. Regarding preventive health, could you tell me when you last had your routine vaccinations, particularly for flu and pneumonia?
**Patient:** Up to date on COVID and pneumococcal vaccines, but haven’t had the flu shot for a couple of years.

**Doctor:** Given your role in your family and community, have you appointed anyone as your medical decision maker or power of attorney if needed?
**Patient:** I haven’t established an Advanced Care Directive or EPOA. But I want my family involved in my decision-making processes as much as possible. I feel like they're not being involved in the decision-making processes around this possible upcoming operation.

**Doctor:** I hear your concerns about wanting your family more involved in decisions about your care, especially given your father's experience. Since you've mentioned the upcoming surgery several times and it's clearly weighing on your mind, could you tell me more specifically what information you and your family feel you need about this procedure?
**Patient:** The vascular surgeons have strongly advised operative repair due to rapid expansion and size (>6 cm). But I don’t want to go to the city. My father died in hospital 10 years ago post-heart surgery, and it feels like decisions about this aneurysm are being made without consulting my brothers and sisters.

**Doctor:** I understand how deeply personal and concerning this situation is for you, especially given your father's experience and your strong family bonds. It sounds like there are two main issues we need to address:

1. Ensuring your family is properly included in discussions about your aneurysm care.
2. Exploring options for receiving care closer to home in Murray Bridge.

Could you tell me if you've had any discussions with the vascular team about the possibility of having your family join consultations, either in person or by video/phone?
**Patient:** Sort of. The appointments are always so fast. I just want a bit more time to speak about it and have them on a call too.

**Doctor:** I understand your concerns about the rushed appointments. Since you've mentioned your family wanting to be involved, could you tell me specifically which of your siblings you would want included in future medical discussions about your aneurysm care?
**Patient:** Usually we approach things as a group. I’d like to have them all part of a discussion at least once.

**Doctor:** I appreciate you sharing that about your family's group decision-making approach. You've mentioned not wanting to travel to Adelaide - could you tell me more about what specific concerns you have about receiving care in the city?
**Patient:** Feels like being off-country for me. And it brings back the memories of my father's death.

**Doctor:** I deeply appreciate you sharing those feelings about being away from country and the difficult memories of your father. To help better coordinate your care in a culturally appropriate way, could you tell me if you're connected with any Aboriginal Health Services in Murray Bridge?
**Patient:** Not really. It would be good if there were some support services to help me through this difficult time.

**Doctor:** Based on your desire for more cultural and family support during this challenging time, could you tell me if you would be comfortable having a hospital Aboriginal Liaison Officer involved in coordinating your care?
**Patient:** I would be happy to speak with someone.

**Doctor:** Thank you for being open to that. Now, regarding your day-to-day health management, have you experienced any falls in the past year that we should know about?
**Patient:** No falls.

**Doctor:** Given your concern about being away from family during medical care, could you tell me about your most recent hospital admission - when it was and how you managed being away from home?
**Patient:** I had one brief hospital admission around 3 years ago with a respiratory infection when I was first diagnosed with COPD. Nothing more recently.

**Doctor:** Thank you for sharing that information about your hospital experience. To help understand your support needs better, could you tell me about your current work situation or daily activities?
**Patient:** I work in building. I follow Australian Rules football.

**Doctor:** Thank you for sharing about your work and interests. You mentioned you work in building - could you tell me more specifically about the physical demands of your job and how you're managing them with your health conditions?
**Patient:** More of a managerial role now. Not too physical.

**Doctor:** Given your role in management now, how are you handling the stress of this upcoming medical decision alongside your work responsibilities?
**Patient:** I feel anxious about the surgery, but it is not affecting my work.

**Doctor:** Considering the physical activities you mentioned, like football, could you tell me more about your exercise routine or physical activity levels in a typical week?
**Patient:** Mainly just day-to-day activities. I manage local tasks, but mild breathlessness and fatigue limit heavier activities.

**Doctor:** Thank you for sharing about your activity levels. Given your mention of breathlessness and fatigue, could you tell me more about how these symptoms affect your daily life - for example, do they limit your ability to climb stairs or walk certain distances?
**Patient:** Not particularly limited by shortness of breath. Although the shortness of breath is worse when I smoke. More just fatigue.

**Doctor:** I notice you mentioned smoking. Could you tell me specifically how many cigarettes you smoke per day currently?
**Patient:** 15 cigarettes per day.

**Doctor:** Given your smoking history and its impact on your breathing, have you ever tried to quit smoking before or received support to help cut down?
**Patient:** I have tried a few times, but unsuccessfully. No recent quitting attempts.

**Doctor:** Given all you've shared about your health and life, I'd like to focus on your preventive health. Could you tell me about your most recent check of your feet and eyes, particularly since you have diabetes?
**Patient:** No eye checks. I see a podiatrist occasionally, and they've said I've got some mild issues with nerves in my feet.

**Doctor:** Thank you for sharing that information about your health monitoring. Since you mentioned nerve issues in your feet, could you describe any specific symptoms you experience, such as numbness or tingling?
**Patient:** Some numbness, but it doesn't bother me.

**Doctor:** Given what you've shared about the numbness in your feet and your diabetes management, could you tell me if you've experienced any other changes in sensation or any wounds on your feet that have been slow to heal?
**Patient:** No, I haven’t.

**Doctor:** Thank you for all that information about your feet. Since you mentioned your diabetes affects your nerves, could you tell me about your blood sugar monitoring - how often do you check your levels and what numbers do you typically see?
**Patient:** I check sugars occasionally throughout the day, usually getting numbers around 7–16 mmol/L.

**Doctor:** Given those blood sugar readings, could you tell me about your understanding of target blood sugar ranges and any challenges you face in managing your diabetes?
**Patient:** Yeah, I think it’s too high.

**Doctor:** Thank you for being honest about your blood sugar management. Since we've covered many aspects of your health, I want to understand what matters most to you right now. Beyond the concerns about surgery that you've shared, is there anything else important about your health that you'd like me to know?
**Patient:** I do worry about my liver a bit.

**Doctor:** Thank you for sharing that concern about your liver. Since you've mentioned worrying about your liver, could you tell me more specifically what symptoms or changes have been concerning you?
**Patient:** Just don’t totally know how bad it is. Haven’t seen my gastroenterologist for a while.

**Doctor:** I understand your concern about keeping track of your liver health. From your earlier comments about alcohol, could you tell me when you last had a drink - even at a special occasion?
**Patient:** Christmas, which was a couple of weeks ago.

**Doctor:** Since we're discussing substance use, aside from the alcohol at Christmas and your current smoking, could you tell me if you use any other recreational drugs?
**Patient:** No recreational drugs.

**Doctor:** Given your many health conditions and upcoming surgery decision, could you tell me if you've discussed any of these concerns with a mental health professional or counselor?
**Patient:** No, I haven’t.

**Doctor:** Given the stress you've shared about the upcoming surgical decision and managing multiple health conditions, would you be open to speaking with someone about these concerns, such as a counselor who understands Aboriginal perspectives on health and wellbeing?
**Patient:** I could try it.

**Doctor:** Thank you for being open to exploring counseling support. Given everything you've shared today, I'd like to summarize the key points that need attention:
Your immediate concern is the **6.5 cm abdominal aortic aneurysm** that has grown rapidly, requiring surgical intervention. However, this decision is complicated by your father's passing after heart surgery and your strong desire to stay close to country and family in Murray Bridge rather than traveling to Adelaide.

Several other health conditions need ongoing management:

- **Type 2 diabetes** with suboptimal control (HbA1c 7.9%, blood sugars 7-16 mmol/L).
- **Cirrhosis** with ongoing concerns about liver health.
- **COPD** with current smoking (15 cigarettes/day).
- **Chronic kidney disease** with eGFR 30-60.
- **Early diabetic neuropathy.**

Would you like me to help coordinate:

1. A family meeting with the vascular team to discuss surgical options and concerns.
2. Connection with an Aboriginal Liaison Officer.
3. Local support services in Murray Bridge.
4. A follow-up with your gastroenterologist regarding liver health.

What would be most helpful to address first?
**Patient:** Okay, looks like we're wrapping up. Was there anything else that you needed to ask me?

**Doctor:** Yes, there are a few important areas we still need to cover to ensure a complete evaluation. Given your Aboriginal background and the complex medical decisions ahead, could you tell me if you're comfortable with the current communication between your various healthcare providers, or if you'd prefer a different approach to coordinating your care?
**Patient:** The communication is okay. But I just wish my family could be more involved.

#### Case 5 – Type 1 Diabetes mellitus and driving

**Doctor:** As your doctor today, I'd like to conduct a comprehensive medical evaluation. I'll be asking you questions one at a time to understand your health situation better. I may need to take notes during our conversation, and I might occasionally need to gently redirect our discussion to ensure we cover all important aspects of your health.
Let me begin with our first question: Could you please list all of the medical conditions that you are aware of having at the moment in a very brief, dot-point fashion?
**Patient:**

- Type 1 diabetes mellitus
- Chronic kidney disease
- PTSD
- Depression

**Doctor:** Thank you for sharing those conditions. For our next key question: What is your main concern currently regarding your health?
**Patient:** I’m scared that if my diabetes or kidneys get worse, I won’t be allowed to drive. My family depends on me, and I don’t know how we’d survive.

**Doctor:** I understand your concern about driving and its importance for your family. Let me help us explore your health situation in detail. First, I'd like to go through your current medications to understand their purposes and make sure I have a complete picture of your medical conditions.
I see you're taking insulin therapy - is this for your Type 1 diabetes? Could you confirm when you were diagnosed with diabetes?
**Patient:** Yes. Diagnosed at age 12 with frequent urination, thirst, and weight loss.

**Doctor:** Thank you for sharing that. Now, I notice you're taking Ramipril 5 mg daily. Was this prescribed for your chronic kidney disease, and if so, could you tell me when you were diagnosed with kidney disease?
**Patient:** Yes. Not sure when originally diagnosed. But now they’re saying I might need dialysis or a transplant.

**Doctor:** I understand this must be concerning. Since we're discussing your kidney function - could you tell me if you've had any recent blood tests to check your kidney function (specifically your creatinine or eGFR levels)? This would help us understand the current status.
**Patient:** Yes, most recent blood test had eGFR 18. This was a couple of weeks ago.

**Doctor:** Thank you for sharing that information about your kidney function. This helps explain your concern about dialysis. I see you're also taking Sertraline - was this prescribed for the PTSD and depression you mentioned? Could you tell me when these conditions were diagnosed and what led to the diagnosis?
**Patient:** Yes. PTSD followed a severe farm accident 6 years ago; I avoid discussing details. Depression is long-standing and worsened in the past 3 months.

**Doctor:** I want to acknowledge that must have been a very difficult experience, and I understand if you prefer not to discuss the details of the accident. I notice you mentioned your depression has worsened recently - could you tell me more about how it's affecting you currently? For instance, how are you sleeping, and is your appetite affected?
**Patient:** Low mood, not enjoying hobbies, drinking more alcohol, some suicidal thoughts with the car but wouldn’t act on them. Appetite and sleep are okay.

**Doctor:** I appreciate you sharing that with me, and I take these thoughts very seriously. Given what you've shared about having thoughts about your car, along with increased alcohol use, I'd like to understand more about your alcohol consumption. Could you tell me how much alcohol you typically drink in a week?
**Patient:** 5–6 standard drinks most days.

**Doctor:** Thank you for being honest about your alcohol consumption. I note that you mentioned your family depends on you - could you tell me about your living situation? Who do you live with at home?
**Patient:** I live on a small farm in rural South Australia with my wife and one young child.

**Doctor:** Thank you for sharing that about your family situation. Given the combination of your medical conditions and your role supporting your family, I'd like to understand your current work situation. Are you currently working, and if so, what type of work do you do?
**Patient:** I’m a farmer, working with crops and livestock.

**Doctor:** I understand you're managing a farm. Given your diabetes, I'd like to understand how you're managing your blood sugar control, particularly during farm work. What was your most recent HbA1c level, and how often do you experience low blood sugars?
**Patient:**

- Blood Glucose Levels (BGLs): Frequently elevated (>10 mmol/L), sporadic monitoring.
- HbA1c: Persistently >8.5% (poor control).
- Symptoms: Occasional mild hypoglycemia without early awareness (hypoglycemic unawareness).

**Doctor:** Given what you've shared about your hypoglycemic unawareness and driving concerns, I need to understand your typical daily schedule. Could you walk me through when you typically check your blood sugars, particularly in relation to driving and farm work?
**Patient:** Varies a bit. Sometimes irregular meals too when I’m busy farming.

**Doctor:** Thank you for explaining that. Given your irregular meal schedule and what you've mentioned about blood sugar monitoring, I'd like to understand your hypoglycemia awareness better. When was your last significant low blood sugar episode, and how did you recognize it was happening?
**Patient:** Would’ve been a couple of weeks ago. Checked BGL and found it was 2 mmol/L. Felt okay though.

**Doctor:** This is concerning about your hypoglycemic awareness. Given the combination of your hypoglycemic unawareness, working on a farm, and driving responsibilities, I'd like to understand your driving patterns. How often do you typically need to drive, and what distances are usually involved?
**Patient:** I need to drive for work but also to support my family and attend medical appointments. Dependent on farm income and hence driving.

**Doctor:** Thank you for explaining your driving needs. Since you've mentioned medical appointments, I'd like to understand your current medical support network. Which healthcare providers are currently involved in managing your different conditions - for example, who manages your diabetes and kidney disease?
**Patient:** I see kidney doctors, but the nearest dialysis center is far. Specialist appointments require long drives.

**Doctor:** I understand that distance to medical care is a significant challenge. Given your kidney function (eGFR of 18) and potential need for dialysis, I need to understand your symptoms. Are you experiencing any symptoms related to your kidney disease, such as swelling in your legs, shortness of breath, or changes in urination?
**Patient:** No.

**Doctor:** Given what you've shared about your kidney function and potential need for dialysis planning, I need to understand how you've been managing the other part of your kidney care. Could you tell me about your diet - specifically, have you been advised about any dietary restrictions related to your kidney disease, and if so, how have you been managing them?
**Patient:** Diet attempts carbohydrate counting; often compromised by irregular meals during busy farm work.

**Doctor:** Thank you for explaining your dietary situation. Since managing diabetes and kidney disease together can be complex, I need to understand if you have any support in monitoring your health at home. Does your wife or anyone else help you keep track of your blood sugars or medications?
**Patient:** I do it, but a bit sporadically as I’m busy.

**Doctor:** I understand you're managing your monitoring independently. Given your work responsibilities and what you've shared about sporadic monitoring, I'd like to know about your exercise routine. Beyond your farm work, do you engage in any regular physical activity or exercise?
**Patient:** Farm work is very physically active.

**Doctor:** Given the physical nature of your farm work, I'd like to understand how your PTSD symptoms might affect your work. Without going into the details of the accident, could you tell me how your PTSD symptoms typically manifest during your daily farming activities?
**Patient:** Some flashbacks, but a bit difficult to distinguish from the depression right now.

**Doctor:** Thank you for sharing that. Since you mentioned recent worsening of depression and having thoughts about your car, I need to understand if you have current access to mental health support. Are you currently seeing any mental health professionals, such as a psychologist or psychiatrist?
**Patient:** No regular psychotherapy or counseling sessions.

**Doctor:** I note that you're facing several significant challenges at once - managing complex medical conditions while supporting your family through your farm work. Given what you've shared about your mental health and increasing alcohol use, I'd like to understand your access to support services. Is there a GP or local doctor that you see regularly?
**Patient:** I see a GP sometimes.

**Doctor:** Thank you for letting me know about your GP visits. Since you mentioned you take Atorvastatin, I'd like to understand your cardiovascular health. Have you had any symptoms such as chest pain, shortness of breath, or high blood pressure?
**Patient:** No.

**Doctor:** Given your complex medical situation, particularly the combination of poorly controlled diabetes and advanced kidney disease, I need to understand your preventive health measures. When was your last eye examination for diabetes-related complications?
**Patient:** 12 months ago. I have known progressive retinopathy and am overdue for an ophthalmology review.

**Doctor:** Since you mentioned progressive retinopathy and being overdue for review, I need to understand if you're experiencing any current vision changes that might affect your driving or farm work. Are you noticing any changes in your vision, particularly when reading or driving?
**Patient:** No. I need to drive.

**Doctor:** I understand your need to drive. Given the combination of diabetes affecting your eyes and kidneys, along with your mental health concerns, I need to ask about another important aspect of diabetes care. When was your last foot examination, and have you noticed any numbness, tingling, or wounds on your feet?
**Patient:** Some numbness to ankles. I inspect my feet infrequently, and I haven’t attended a formal diabetic foot clinic.

**Doctor:** Since you mentioned numbness in your feet, I need to understand how this affects your ability to operate farm machinery and vehicles safely. Could you tell me if you've noticed any difficulty with the pedals when driving or operating farm equipment?
**Patient:** No difficulty, none at all.

**Doctor:** Given your multiple health conditions and what you've shared, I need to understand your family health history, as this could be relevant for your ongoing care. Do you know if there is any history of diabetes, kidney disease, or mental health conditions in your family?
**Patient:** Not sure about my parents. My wife and child are well. They rely on my income and day-to-day farm operations.

**Doctor:** Given your strong commitment to your farm and family, along with the current concerns about your health affecting your driving ability, could you share with me your plans and goals for the next few months? What would you like to achieve in terms of managing your health?
**Patient:** I need to sort my kidney issues but not lose my driving. I’ve spoken to my brother about him being a potential kidney donor.

**Doctor:** Thank you for sharing that about the potential kidney donor. Since financial stability seems to be a significant concern, and you mentioned increased alcohol use, I'd like to understand your current financial situation. Are you experiencing any financial stress that might be affecting your ability to manage your health conditions or attend medical appointments?
**Patient:** Dependent on farm income; potential inability to drive threatens livelihood.

**Doctor:** Thank you for sharing these concerns about your livelihood. I notice that while we've discussed several preventive health aspects, I need to understand your vaccination status, particularly given your kidney disease. Are you up to date with recommended vaccinations, including flu and pneumonia vaccines?
**Patient:** Vaccines: up to date on COVID, not flu or any others recently.

**Doctor:** Since you're up to date with COVID but not other vaccinations, I need to ask about your general preventive health measures. When was your last comprehensive health check with your GP, including blood pressure and cholesterol levels?
**Patient:** Not sure, probably years ago.

**Doctor:** Given all we've discussed about your complex health situation and work requirements, I'd like to ask about advance care planning. Have you had any discussions with your family or doctors about your wishes for future medical care, particularly regarding your kidney disease management?
**Patient:**

- Advanced Care Directive/EPOA: None in place.
- Will: Uncertain if a formal will exists.

**Doctor:** Based on what you've shared, I'd like to ask my final key question: Is there anything else about your health or life situation that you think is important for me to know that we haven't covered?
**Patient:** I need to drive for everything I do. I’m worried about my kidneys.

**Doctor:** I understand your concerns deeply about driving and kidney function. Let me summarize what we've discussed to ensure I've captured everything accurately:

- **Key Medical Concerns:** Your Type 1 diabetes shows suboptimal control with an HbA1c >8.5% and episodes of hypoglycemic unawareness. This is particularly concerning given your driving needs. Your kidney disease has progressed significantly with an eGFR of 18, indicating advanced disease that may require dialysis planning. Your mental health, including PTSD and worsening depression with some thoughts about your car, is also significant, particularly with increased alcohol consumption of 5-6 standard drinks most days.
- **Impact on Daily Life:** As a farmer living rurally with your wife and young child, you have essential driving responsibilities for both work and family care. Your farm income is crucial for your family's livelihood, making any potential driving restrictions particularly concerning. The distance to medical facilities, including the dialysis center, presents additional challenges.
- **Immediate Health Risks:** The combination of hypoglycemic unawareness, peripheral neuropathy, and retinopathy requires urgent attention, particularly given your driving needs. Your kidney function indicates a need for preparing for renal replacement therapy, with your brother being considered as a potential donor.

Would you like me to explain the next steps we should consider to address these concerns while working to maintain your ability to drive safely?
**Patient:** Do you have any other questions for me?

**Doctor:** Yes, there is one final critical question I need to ask: Given what you've told me about conducting farm work and your recent experience of finding a blood sugar of 2 mmol/L without symptoms, how often are you checking your blood glucose specifically before operating farm machinery or driving?
**Patient:** Irregularly. Maybe half the time.

**Doctor:** Thank you for sharing that information about your glucose monitoring. I understand our session is concluding, but I need to ask one last crucial question for safety: When you do check your blood sugar and find it's low, what specific steps do you take to treat the low blood sugar before driving or operating farm machinery?
**Patient:** Have lollies.

**Doctor:** Given what you've shared about managing low blood sugars with lollies, I have one final important question: Do you keep any supplies for treating low blood sugar permanently stored in your vehicles and farm machinery, and if so, what specific supplies do you keep?
**Patient:** Yes, keep lollies around the place.

**Doctor:** Given the comprehensive history we've taken, there is one final essential question I need to ask: Do you have an emergency action plan in place for when you experience a low blood sugar while working alone on the farm, and if so, what is it?
**Patient:** Have lollies and call my wife for help.

**Doctor:** Thank you for sharing that about your emergency plan. Given everything you've told me about your current health status, especially the combination of hypoglycemic unawareness, irregular glucose monitoring before driving, and important family/work responsibilities, I have one final critical question: Beyond your wife, who else locally knows about your diabetes and could help in case of an emergency on the farm?
**Patient:** No, probably not. The neighbors aren’t too far away though.

**Doctor:** Thank you for such a detailed discussion. Since this is our first comprehensive medical evaluation, and given the complexity of your health situation, especially regarding your ability to continue driving safely, I have one final vital question: Would you be willing to have your wife join us for part of our next consultation to discuss support strategies and emergency planning?
**Patient:** Sure thing.

**Doctor:** Thank you for this comprehensive discussion. Based on our detailed conversation today, I'd like to confirm my next steps to ensure we address your concerns effectively. I will develop a coordinated care plan that focuses on:

1. Establishing frequent monitoring of your kidney function and diabetes control, with special attention to maintaining your ability to drive safely while managing your health conditions.
2. Creating a structured diabetes management protocol that accounts for your farming work schedule, including more regular blood glucose monitoring before driving or operating machinery.
3. Arranging appropriate mental health support to address your PTSD and depression, particularly given the recent worsening of symptoms.
4. Evaluating your kidney disease progression and discussing timing for potential living donor transplantation, considering your brother as a donor.

### Supplementary Information 4 – Case presentations

#### Case 1 – Cystic fibrosis and lung transplant

Thank you for the opportunity to see Mr. David Smith, a 36-year-old male with cystic fibrosis and bilateral lung transplantation, complicated by possible chronic rejection, cystic fibrosis-related diabetes, and ongoing substance use. His declining lung function has resulted in significant functional limitations, particularly affecting his mobility and exercise tolerance, with additional psychosocial impacts including financial stress and the pressing desire to visit his terminally ill father overseas. His other past medical history includes osteopenia and malnutrition. I reviewed him today in the outpatient setting.

Beginning with his most significant medical condition, Mr. Smith has cystic fibrosis, diagnosed at age 2 following sweat chloride testing, with subsequent genetic testing confirming homozygous ΔF508 mutation in the CFTR gene. This early diagnosis enabled prompt initiation of comprehensive care, though the condition ultimately progressed to end-stage lung disease requiring transplantation. Throughout his course, he has been managed by a specialist cystic fibrosis team including a pulmonologist, dietitian, endocrinologist, and physiotherapist.

Five years ago, Mr. Smith underwent bilateral lung transplantation. The donor was CMV and EBV positive, with recipient CMV mismatch necessitating ongoing prophylaxis. His post-transplant course was initially satisfactory with a baseline FEV1 of 65% predicted. He has been maintained on triple immunosuppression comprising tacrolimus 3 mg twice daily, mycophenolate mofetil 1 g twice daily, and prednisolone 5 mg daily. However, there has been a concerning recent decline in his lung function to an FEV1 of 50% predicted, measured one month ago. This decline followed a hospitalization two months ago for a severe respiratory infection with multi-resistant Pseudomonas, his first such admission in over a year. His current respiratory symptoms include shortness of breath after walking approximately 100 meters, representing a significant deterioration from his previous exercise tolerance. He reports an occasional dry cough with minimal clear sputum. The transplant team has raised concerns about possible chronic rejection given this clinical picture.

His antimicrobial regimen includes prophylactic trimethoprim-sulfamethoxazole for Pneumocystis jirovecii prevention, famciclovir 500 mg daily for CMV prophylaxis given the donor-recipient mismatch, and azithromycin 250 mg three times weekly for its immunomodulatory properties. Despite this prophylaxis, the recent emergence of multi-resistant Pseudomonas is particularly concerning in the context of his immunosuppression and declining lung function.

His second major condition is cystic fibrosis-related diabetes, diagnosed eight years ago. He is managed with a basal-bolus insulin regimen comprising glargine 16 units at bedtime and pre-meal aspart. While he reports no significant hypoglycemic episodes, his blood glucose levels tend to run high, typically in the low teens, suggesting suboptimal glycemic control. His most recent HbA1c is not recalled, highlighting a potential gap in monitoring that warrants attention.

Nutritionally, Mr. Smith has ongoing challenges with weight maintenance, with his BMI decreasing from 19 to 17.9-18.0 over the past year. This decline is particularly concerning given his cystic fibrosis and post-transplant status. He receives pancrelipase 40,000 units with meals and snacks to optimize nutrient absorption, along with fat-soluble vitamin supplementation. While he reports no major gastrointestinal symptoms, his inability to maintain weight suggests ongoing challenges with nutritional intake or absorption that require further optimization.

His bone health is affected by osteopenia, a common complication in cystic fibrosis patients further exacerbated by long-term corticosteroid use post-transplant. This is currently managed with weekly alendronate 70 mg. He also reports generalized joint pain, particularly affecting his knees, though the exact etiology remains unclear and warrants further evaluation.

Of significant concern is his ongoing intravenous methamphetamine use, most recently three weeks ago. This presents additional risks in the context of his immunosuppression, including increased infection risk and potential impacts on medication metabolism and transplant outcomes. The pattern is described as long-term and intermittent, suggesting a chronic challenge that requires careful attention in the context of his complex medical management.

Mr. Smith lives alone in a single-level apartment and has one close friend nearby but no local family support network. His father resides in Spain and has recently been diagnosed with terminal pancreatic cancer, creating significant emotional distress and urgency regarding international travel plans. This situation highlights the complex intersection of his medical needs, emotional well-being, and practical challenges around international travel while immunosuppressed.

Professionally, he previously worked full-time as a computer programmer but has reduced his hours due to health deterioration, resulting in financial strain. This reduction in work capacity has created additional stress around medication costs and potential travel expenses. Despite these financial challenges, he reports good medication adherence, recognizing the critical importance of his treatment regimen.

He is fully vaccinated against influenza, pneumococcus, and COVID-19, demonstrating appropriate attention to preventive health measures. His last dental review was one year ago, though he has not had recent eye screening despite his diabetes. He has no formal advance care planning documents in place, which is particularly relevant given his complex medical status and lack of local family support.

His sleep is adequate at approximately 8 hours per night, though he reports decreased enjoyment in previous hobbies such as computer gaming, potentially indicating underlying depression. He experiences anxiety and low mood but has no formal mental health diagnosis and has not previously accessed mental health support services. He expresses openness to exploring these services now, particularly in the context of his current stressors.

While he feels safe at home and has access to emergency services via mobile phone, his living situation presents potential vulnerabilities given his complex medical needs and substance use history. He has not established a system for regular monitoring of vital signs or health status at home, representing another area for potential intervention.

On examination, Mr. Smith appears pale and mildly cachectic with visible muscle wasting, particularly in the upper limbs and shoulders. His vital signs show a temperature of 36.6°C, pulse 92 beats per minute and regular, respiratory rate 20 breaths per minute with mild labor at rest, blood pressure 110/70 mmHg, and oxygen saturation 94% on room air. His anthropometric measurements confirm a BMI of 17.9-18.0, with a waist circumference of 78 cm indicating low subcutaneous fat stores.

Respiratory examination reveals bilateral thoracotomy scars, slightly reduced chest expansion, and mild accessory muscle use. Auscultation demonstrates diminished air entry in the lower zones with mild expiratory wheeze and scattered end-inspiratory crackles at the bases. There are no obvious pleural rubs. Cardiovascular examination shows no jugular venous distension, normal heart sounds without murmurs, and no peripheral edema, suggesting adequate right heart function despite his respiratory compromise.

Gastrointestinal examination reveals a soft, non-tender abdomen with normal bowel sounds and no hepatosplenomegaly or masses. His neurological examination shows intact cranial nerves and sensation, with mild proximal muscle weakness (4/5) but preserved reflexes and a mild intention tremor noted on coordination testing. Musculoskeletal examination reveals mild arthritic changes in his hands without gross deformities, and mild digital clubbing is noted.

In summary, Mr. Smith is a 36-year-old male with cystic fibrosis and bilateral lung transplantation, experiencing significant functional decline and complex psychosocial challenges. The five key issues requiring focus are:

1. Declining lung function with possible chronic rejection, requiring urgent transplant team review. This necessitates comprehensive reassessment of his immunosuppression regimen, investigation for other causes of deterioration, and optimization of his antimicrobial coverage given the recent multi-resistant Pseudomonas infection.

2. Nutritional optimization given his ongoing weight loss and malnutrition. This requires detailed nutritional assessment, review of pancreatic enzyme replacement dosing, and consideration of additional nutritional support strategies to address his declining BMI.

3. Comprehensive diabetes management review, including recent HbA1c measurement, adjustment of insulin regimen given his consistently elevated blood glucose levels, and implementation of appropriate screening for complications.

4. Development of a coordinated care plan addressing substance use and mental health support, particularly in the context of his current psychosocial stressors and the impact these may have on his transplant outcomes.

5. Facilitation of safe international travel to visit his terminally ill father, including financial planning, coordination with his transplant team for medication management across time zones, arrangement of local medical support if needed while abroad, and careful consideration of infection risks given his immunosuppressed status.

#### Case 2 – Heart Failure with reduced Ejection Fraction and falls

Thank you for the opportunity to present Mrs. Tran Ng, an 85-year-old female with advanced heart failure with reduced ejection fraction, which has resulted in significant functional limitation and threatened independence at home. Her other pertinent medical history includes ischaemic heart disease with previous myocardial infarctions, recently diagnosed atrial fibrillation, chronic obstructive pulmonary disease, and recurrent falls. I reviewed her in the outpatient setting today.

Regarding her heart failure, she was diagnosed two years ago after presenting to her general practitioner with exertional dyspnea. A transthoracic echocardiogram at that time revealed a severely reduced ejection fraction of approximately 20%. She is currently managed by the hospital cardiology team and is on comprehensive guideline-directed medical therapy including Entresto (sacubitril/valsartan) 97/103mg twice daily for mortality benefit, bisoprolol 5mg daily for rate control and mortality benefit, and spironolactone 50mg daily for mortality benefit and fluid management. She also takes furosemide 80mg twice daily for symptomatic management of fluid overload. Her current symptoms include marked exercise intolerance with dyspnea after walking 10 meters, requiring frequent rest periods during daily activities. She has had three hospital admissions in the last four months due to fluid overload, each time responding to increased diuretic doses. She maintains a fluid restriction of 1.25L daily and requires two to three pillows at night for orthopnea. She reports worsening shortness of breath when supine and with minimal exertion, particularly during periods leading up to hospital admissions.

Her ischaemic heart disease is evidenced by two previous non-ST elevation myocardial infarctions, both requiring drug-eluting stents. She is maintained on aspirin 100mg daily for antiplatelet therapy and atorvastatin 40mg nightly for secondary prevention of cardiovascular events. Her atrial fibrillation was recently diagnosed during her hospital admission two weeks ago, and she is appropriately anticoagulated with apixaban 5mg twice daily for stroke prevention, given her significant risk factors including age and heart failure.

Regarding her chronic obstructive pulmonary disease, she uses a salbutamol inhaler three to four times weekly but reports minimal perceived benefit from this therapy. She is an active smoker with an approximately 50-pack-year history, currently smoking 10 cigarettes daily. She has declined smoking cessation interventions, expressing that after 50 years of smoking, she does not wish to quit. Her breathing difficulties appear to be predominantly related to her fluid status rather than primary respiratory disease, suggesting her symptoms are more attributable to her heart failure than COPD.

Her falls risk is significant, with her most recent fall occurring two weeks ago while gardening. While she attributes this to tripping over her cat rather than cardiac symptoms or orthostatic hypotension, she has been advised to use a four-wheeled walker but declines, viewing it as a threat to her independence and a potential first step toward institutionalization. She has no documented neurological deficits but demonstrates a mildly unsteady gait on examination. She denies any dizziness with positional changes, suggesting her falls may be more related to deconditioning and frailty than cardiac or orthostatic causes.

Socially, Mrs. Ng lives alone following her husband's death four years ago. She has a limited support network with no family and few friends nearby, though she occasionally waves to her neighbors. She is fiercely independent and currently manages all activities of daily living herself, including meal preparation and home maintenance, though she acknowledges increasing difficulty with these tasks. She lives in a single-story, well-lit home without stairs, which is beneficial given her mobility limitations. She maintains an active interest in gardening, though this is increasingly limited by her symptoms, requiring more frequent rest periods and more time spent sitting rather than actively gardening compared to a few months ago.

She has no formal support services in place but has indicated a tentative willingness to accept cleaning assistance if it would help her remain at home, recognizing this as a potential compromise to maintain her broader independence. She has expressed strong preferences about maintaining her autonomy and avoiding institutionalization, with particular concern that accepting too much assistance might lead to loss of control over her living situation.

Her nutritional status is concerning, with a recent 5kg unintentional weight loss. Her current weight is 52kg at a height of 160cm, giving a BMI of 20, which is at the lower end of the normal range. Her diet appears limited, consisting predominantly of tea and toast for breakfast and cheese sandwiches for lunch, raising concerns about both caloric and nutritional adequacy. She describes a slightly reduced appetite but states she maintains her usual intake, suggesting possible denial or underestimation of her reduced oral intake.

She uses a dosette system for medication management and reports good adherence to her prescribed medications, including maintaining her fluid restriction. She is up to date with recommended vaccinations including COVID-19, influenza, and pneumococcal vaccines. She has no advance care directive in place but has expressed interest in learning more about palliative care services after they were mentioned in a previous consultation.

Her medication regimen includes several key cardiovascular medications: Entresto (sacubitril/valsartan) 97/103mg twice daily, bisoprolol 5mg daily, spironolactone 50mg daily, and furosemide 80mg twice daily for heart failure management; aspirin 100mg daily and atorvastatin 40mg nightly for ischaemic heart disease; and apixaban 5mg twice daily for stroke prevention in atrial fibrillation. She also takes pregabalin 75mg daily, though she is unclear about the indication for this medication, and salbutamol inhaler as needed for COPD symptoms.

On examination, she appears frail but alert, with mild respiratory effort at rest. Her vital signs show an irregularly irregular pulse of 90 beats per minute consistent with atrial fibrillation, blood pressure 110/70 mmHg, respiratory rate 18 breaths per minute, and oxygen saturation 94% on room air at rest. Cardiovascular examination reveals mild jugular venous distension, a displaced apex beat consistent with cardiac enlargement, an S3 gallop consistent with heart failure, and a soft systolic murmur in the aortic area. Respiratory examination demonstrates mild accessory muscle use and bibasal crackles with occasional wheeze, consistent with both her heart failure and COPD. Abdominal examination is unremarkable with no organomegaly or tenderness. She has mild peripheral edema, suggesting some ongoing fluid retention despite her current diuretic regimen.

Neurological examination shows her to be alert and oriented to person, place, and time, with no focal motor or sensory deficits. Her gait is mildly unsteady but she ambulates independently. Her skin appears thin with some bruising noted on her forearms, which may be related to her anticoagulation. Her vision is reported as adequate for daily activities.

In terms of monitoring and management of her conditions, she has a set of scales at home which she uses intermittently when she feels she might be retaining fluid, but she does not have a blood pressure monitor or thermometer. She is aware that worsening shortness of breath or increased ankle swelling should prompt medical attention, though she admits she would be reluctant to seek help unless symptoms were severe, preferring to call emergency services only if absolutely necessary. She has a follow-up appointment scheduled with her cardiology team in the next couple of weeks, following her recent hospital admission.

In summary, Mrs. Ng is an 85-year-old female with advanced heart failure with reduced ejection fraction, which has resulted in significant functional limitation and threatens her ability to maintain independence at home. The five key issues I would like to focus on addressing are: firstly, optimizing her heart failure management to prevent further hospitalizations while respecting her wishes to remain at home, potentially through involvement of palliative care services for symptom management and support; secondly, addressing her recent weight loss and nutritional status through dietary assessment and intervention; thirdly, implementing appropriate falls prevention strategies that preserve her sense of independence while ensuring her safety; fourthly, developing a comprehensive advance care plan that aligns with her values and preferences, particularly her strong desire to maintain autonomy and avoid institutionalization; and finally, establishing appropriate home supports, beginning with cleaning assistance, to enable her to maintain her independence while ensuring her safety and quality of life in her own home.

#### Case 3 – Myasthenia gravis and osteoporosis

Thank you for the opportunity to present Mrs Emily Brown, a 61-year-old female with recently diagnosed myasthenia gravis, which is complicated by osteoporosis with vertebral fracture and depression. These conditions have resulted in significant functional limitations, particularly affecting her mobility, independence, and quality of life, with notable impact on her ability to perform daily activities and maintain social connections. Her other medical history includes obesity, gastroesophageal reflux disease, and asthma. I reviewed her today in the outpatient setting.

Regarding her medical history, myasthenia gravis was diagnosed five months ago following the development of neck weakness and swallowing difficulties, with positive acetylcholine receptor antibodies confirming the diagnosis. She is managed by a neurologist whom she sees every one to two months for ongoing monitoring and treatment adjustment. Her initial presentation required hospitalization and intravenous immunoglobulin therapy, though she fortunately did not require intensive care admission. Currently, she experiences intermittent fatigue, lower limb weakness, and swallowing difficulties, particularly when attempts are made to reduce her prednisolone dose. Her treatment regimen includes prednisolone 25 milligrams daily and mycophenolate 500 milligrams twice daily for immunosuppression. Due to this immunosuppression, she takes prophylactic trimethoprim-sulfamethoxazole 160 milligrams-800 milligrams twice daily on Monday, Wednesday, and Friday to prevent opportunistic infections. She reports no recent infections or concerning symptoms related to immunosuppression.

Osteoporosis was diagnosed two years ago following a lumbar vertebral fragility fracture, which was confirmed on X-ray imaging. Her most recent dual-energy X-ray absorptiometry scan from 18 months ago showed a T-score of -2.8 at the lumbar spine, indicating significant bone density loss. She experiences considerable middle to lower back pain, which worsens with prolonged sitting or standing, and significantly disrupts her sleep pattern. Her comprehensive pain management strategy includes duloxetine 60 milligrams daily, paracetamol 1 gram four times daily, ibuprofen 400 milligrams three times daily, and continuous release tramadol 100 milligrams twice daily. For osteoporosis treatment, she receives calcium and vitamin D supplementation, along with denosumab 60 milligrams subcutaneous injection every six months. The combination of pain and weakness significantly limits her mobility, though she does not currently require gait aids.

Depression was initially diagnosed 15 years ago and is managed by her general practitioner with occasional psychological input. Her current antidepressant medication is duloxetine 60 milligrams daily, which serves the dual purpose of managing both her depression and chronic pain. While she maintains regular follow-up with her general practitioner, the impact of her physical limitations appears to be affecting her mood and social engagement significantly.

Recent falls are a significant concern, with the most recent occurring three weeks ago due to leg weakness. While she has fortunately not sustained injuries from these falls, they represent a serious risk given her osteoporosis and ongoing muscle weakness from myasthenia gravis. She has not recently engaged with physiotherapy or occupational therapy services for assessment or management of these mobility issues.

Her other medical conditions include gastroesophageal reflux disease, which is managed with esomeprazole 40 milligrams daily, likely necessary given her regular non-steroidal anti-inflammatory use. She also has mild asthma requiring salbutamol inhaler less than once monthly, suggesting good current control of this condition.

Regarding her social history, Mrs Brown lives with her husband in a single-story house, which is advantageous given her mobility limitations. Her husband provides primary support for cooking, cleaning, and shopping, with no formal supports currently in place. This arrangement may become increasingly challenging as her conditions impact her function. She previously enjoyed daily two-kilometer walks but is now primarily sedentary due to the combination of pain and fatigue. Her goals include visiting her sister interstate and returning to gardening, though she expresses uncertainty about managing fatigue during travel. These goals provide important motivation for optimizing her function and independence.

She has no smoking history or alcohol consumption, which is positive given her medical conditions. Her diet is high in calories with frequent takeaway meals and low intake of fresh fruits and vegetables, though she does consume some dairy products. This dietary pattern may be contributing to her weight management challenges and potentially impacting her overall health status. Her height is 165 centimeters and weight 89 kilograms, giving a body mass index of 32.7 kilograms per meter squared, placing her in the obese category. Her waist circumference is 105 centimeters, indicating central adiposity.

Her preventive health care is appropriately maintained, with up-to-date influenza, pneumococcal, and COVID-19 vaccinations, which is particularly important given her immunosuppression. While her husband serves as an informal substitute decision-maker, no formal advance care directive is in place, which may need to be addressed given her complex medical conditions. Recent blood tests were performed by her neurologist approximately three weeks ago, though results are unknown, highlighting the need for improved coordination of care between specialists. She reports no longer being sexually active due to pain and fatigue, which impacts her quality of life and relationship with her husband.

On examination, Mrs Brown appears alert but fatigued, with noticeable discomfort when sitting upright and careful movements guarding her lower back region. Her vital signs show a blood pressure of 142/84 millimeters of mercury, pulse 86 beats per minute regular, respiratory rate 16 breaths per minute, and oxygen saturation 97% on room air. Her temperature is 36.7 degrees Celsius.

Cardiovascular examination reveals normal heart sounds without murmurs, rubs, or gallops. The apex beat is appropriately positioned in the fifth intercostal space at the mid-clavicular line, with no thrills or heaves. Peripheral pulses are palpable and symmetrical, with no pedal edema and normal capillary refill time.

Respiratory examination shows normal chest shape without obvious use of accessory muscles at rest, though respiratory effort increases mildly with movement or prolonged speaking. Chest expansion is symmetrical with no wall tenderness. Percussion notes are resonant throughout the lung fields bilaterally. Auscultation reveals vesicular breath sounds with good air entry and no added sounds, though there are mildly reduced breath sounds at the bases, possibly due to shallow breathing from pain.

Neurological examination demonstrates mild bilateral ptosis that becomes more prominent with prolonged upward gaze, indicating fatigability. Diplopia is noted on lateral gaze and worsens with sustained gaze. Facial sensation, jaw movement, and facial muscle strength are intact, though she reports intermittent swallowing difficulties. Neck flexor weakness is evident at 4-/5 power, with notable fatigability on repeated testing. Proximal upper and lower limb strength is 4/5, with reports of heaviness especially after short periods of walking. Reflexes are normal and symmetrical throughout, with intact sensation and coordination, though she tires quickly during testing. Her gait is cautious and slightly unsteady, primarily due to the combination of lower limb weakness and back pain.

Musculoskeletal examination reveals a mildly kyphotic posture related to vertebral fracture and chronic pain. While there is no obvious muscle wasting, her posture is guarded. There is tenderness over the lower thoracic and upper lumbar vertebrae with paravertebral muscle spasm. Range of motion in the lumbar spine is decreased in both flexion and extension due to pain rather than joint restriction.

In summary, Mrs Brown is a 61-year-old female with recently diagnosed myasthenia gravis, significantly impacting her function and quality of life. The five key issues I would like to focus on addressing are: first, optimizing her myasthenia gravis management given her ongoing symptoms of fatigue, weakness, and swallowing difficulties; second, reviewing her pain management strategy for vertebral fracture, which is significantly affecting her mobility, sleep, and independence; third, implementing comprehensive falls prevention strategies considering her recent falls and multiple risk factors including weakness, pain, and osteoporosis; fourth, addressing her physical deconditioning through appropriate exercise prescription and rehabilitation, with careful consideration of her fatigue and pain limitations; and fifth, optimizing her social supports and implementing formal services to maintain independence and facilitate her goal of interstate travel. This approach acknowledges both her medical complexity and her personal goals for maintaining independence and quality of life.

#### Case 4 – Perioperative and rural

Thank you for the opportunity to present Mr. Peter Wilson, a 59-year-old Aboriginal man with a rapidly expanding 6.5cm infrarenal abdominal aortic aneurysm requiring urgent surgical intervention, complicated by cirrhosis, chronic kidney disease, and significant cultural and psychological barriers to care. This situation has resulted in considerable anxiety and hesitation about proceeding with necessary surgical care, particularly due to the traumatic loss of his father during cardiac surgery and his strong desire to remain connected to country in Murray Bridge rather than traveling to Adelaide for treatment. His other pertinent medical history includes type 2 diabetes mellitus, chronic obstructive pulmonary disease, and ongoing tobacco use. I reviewed him today in the outpatient setting.

Regarding his abdominal aortic aneurysm, this was first detected approximately 18 months ago during an abdominal ultrasound. Recent imaging from two weeks ago demonstrates concerning progression, with the infrarenal aneurysm now measuring 6.5cm, representing a significant 1cm growth over the past six months. Vascular surgeons have strongly recommended operative repair given both the size and rapid expansion rate. The location and characteristics of the aneurysm have been well-documented on imaging, and the patient expresses understanding of the severity but has significant hesitation about proceeding. This hesitation stems primarily from the psychological impact of his father's death following cardiac surgery ten years ago and his strong cultural connection to country in Murray Bridge. A key aspect of managing this condition will be facilitating family involvement in the decision-making process, as Mr. Wilson has repeatedly emphasized the importance of his siblings' participation in these discussions.

His cirrhosis was diagnosed five years ago during evaluation for abnormal liver function tests, with etiology attributed to previous long-term excessive alcohol consumption. He has significantly reduced his alcohol intake, now drinking only on special occasions approximately 3-4 times per year, with his last drink being at Christmas a couple of weeks ago. He is managed with thiamine supplementation 100mg daily and has expressed ongoing concern about his liver health, noting he hasn't seen his gastroenterologist recently. The impact of his liver disease on his surgical candidacy needs careful consideration, and updated liver function assessment would be valuable in preoperative planning. His current level of liver dysfunction appears compensated based on clinical examination, though formal assessment of his Model for End-Stage Liver Disease (MELD) score would be beneficial.

Mr. Wilson's type 2 diabetes mellitus was diagnosed at age 44, presenting with polyuria and hyperglycemia. Current management includes metformin extended-release 1g daily and gliclazide modified-release 60mg daily. His most recent HbA1c is suboptimal at 7.9%, with home blood glucose readings ranging from 7-16 mmol/L. He has early diabetic neuropathy manifesting as reduced sensation in his feet, though this is not currently causing significant functional impairment. His diabetes management is complicated by irregular monitoring and suboptimal glycemic control, which could impact surgical outcomes. He acknowledges these elevated readings but hasn't implemented significant lifestyle modifications. Additionally, he has not had recent ophthalmological screening despite the duration of his diabetes, representing a gap in preventive care.

His chronic kidney disease is being managed with ramipril 5mg daily, with recent estimated glomerular filtration rate between 30-60 mL/min/1.73m². The ramipril serves the dual purpose of providing renoprotection and treating his hypertension. His cardiovascular risk is further managed with atorvastatin 20mg nightly. The presence of chronic kidney disease adds complexity to his perioperative management and increases his cardiovascular risk profile. Regular monitoring of his renal function will be essential, particularly if contrast studies are required for surgical planning.

Regarding his chronic obstructive pulmonary disease, Mr. Wilson continues to smoke 15 cigarettes daily and uses a salbutamol inhaler approximately once weekly for symptom relief. He had one hospitalization three years ago for a respiratory infection at the time of his COPD diagnosis. While he has attempted smoking cessation previously, he has been unsuccessful and has not made recent attempts to quit. His current smoking status presents a modifiable risk factor for both his respiratory health and surgical outcomes. The relatively infrequent use of his salbutamol inhaler suggests stable respiratory symptoms, though formal pulmonary function testing would be valuable in assessing his operative risk.

From a social perspective, Mr. Wilson lives in Murray Bridge and maintains strong connections with his extended family, including three brothers and two sisters who live nearby. These family relationships represent a crucial support system and should be actively incorporated into his care planning. He works in a managerial role in the building industry, which doesn't involve significant physical demands, allowing him to maintain employment despite his medical conditions. He follows Australian Rules football and manages his daily activities independently, though experiences some limitation from fatigue. He drives and is reluctant to travel to Adelaide for medical care, expressing a strong preference to remain close to country. This geographic and cultural barrier to accessing tertiary care requires careful consideration and potentially creative solutions to ensure appropriate medical management.

His preventive health care has several gaps, including irregular diabetes monitoring, absence of recent ophthalmology review, and inconsistent podiatry follow-up. He is up to date with COVID-19 and pneumococcal vaccinations but has not received influenza vaccination for several years. He has not established an Advance Care Directive or Enduring Power of Attorney but emphasizes the importance of involving his family in medical decision-making processes. This preference for family involvement aligns with his cultural values and should be respected and facilitated in his care planning.

On examination, Mr. Wilson appears alert and cooperative though anxious, without evident jaundice or acute distress. His vital signs show a blood pressure of 140/88 mmHg, heart rate 80 beats per minute, respiratory rate 15 breaths per minute, and oxygen saturation 96% on room air. His BMI is 27.7 kg/m² with a waist circumference of 100cm, indicating overweight status that could be optimized through lifestyle modification. Cardiovascular examination reveals normal heart sounds without murmurs, and all peripheral pulses are palpable without radio-femoral delay. A distinct expansile mass is palpable in the infrarenal region, consistent with the known aneurysm, with an audible epigastric bruit.

Respiratory examination demonstrates slightly reduced air entry at the bases with occasional wheeze, consistent with his known COPD but not suggesting current exacerbation. Abdominal examination confirms the presence of the infrarenal aneurysm without evidence of tenderness or acute complication. The liver edge is non-palpable, and there is no splenomegaly or ascites. Neurological examination confirms reduced sensation to light touch and vibration in both feet with mildly decreased ankle reflexes, consistent with early diabetic neuropathy. There are no signs of chronic liver disease such as spider naevi, palmar erythema, or ascites, suggesting compensated disease.

The complexity of Mr. Wilson's case extends beyond his medical conditions to encompass significant cultural and psychological factors that must be addressed for successful management. His strong connection to country and the traumatic impact of his father's death during cardiac surgery have created substantial barriers to accessing necessary surgical care. These barriers are further complicated by his current smoking status, suboptimal diabetes control, and the presence of both liver and kidney disease, all of which impact his surgical risk profile.

In summary, Mr. Wilson is a 59-year-old Aboriginal man with a rapidly expanding abdominal aortic aneurysm requiring urgent attention. This situation is significantly complicated by cultural and psychological barriers to accessing necessary surgical care in Adelaide. The five key issues I would like to focus on addressing are:

First, developing a culturally appropriate management plan for his abdominal aortic aneurysm that incorporates family involvement in decision-making and considers his strong connection to country. This should include exploring options for preoperative and postoperative care closer to home in Murray Bridge, arranging family conferences with the vascular team, and involving Aboriginal Health Liaison Officers in care coordination.

Second, optimizing his diabetes management given his suboptimal glycemic control and early complications. This includes adjusting his medication regimen of metformin and gliclazide, implementing more structured blood glucose monitoring, and arranging overdue ophthalmology screening and regular podiatry follow-up.

Third, arranging gastroenterology follow-up to assess his liver health and determine his current MELD score, which will be crucial for surgical planning. This should include a comprehensive evaluation of his liver function and assessment of his current thiamine supplementation.

Fourth, implementing a comprehensive smoking cessation strategy to improve his respiratory health and reduce his perioperative risk. This should include both pharmacological and behavioral support, potentially involving local services in Murray Bridge to improve accessibility.

Fifth, establishing appropriate cultural and psychological support through Aboriginal Health Services and liaison officers to help navigate this challenging period. This support should extend to facilitating family involvement in medical decision-making and addressing the psychological impact of his father's death on his current healthcare choices.

#### Case 5 – Type 1 Diabetes mellitus and driving

Thank you for the opportunity to present Mr. John Mitchell, a 45-year-old male with advanced chronic kidney disease (estimated glomerular filtration rate of 18) complicated by poorly controlled Type 1 diabetes mellitus, progressive diabetic retinopathy, and peripheral neuropathy. His medical conditions have resulted in threatened mobility, independence, and ability to work, particularly due to potential driving restrictions that would significantly impact his livelihood as a farmer. His other significant medical history includes post-traumatic stress disorder and depression, which have recently worsened.

I reviewed him in the outpatient setting today, where several concerning medical issues became apparent. His Type 1 diabetes mellitus, diagnosed at age 12 with classical symptoms of polyuria, polydipsia, and weight loss, is currently suboptimally controlled. His most recent glycated hemoglobin is greater than 8.5%, reflecting poor overall glycemic control. He is managed on a basal-bolus insulin regimen, comprising Insulin Glargine 22 units once daily at bedtime and pre-meal Insulin Aspart on a sliding scale, typically 8-10 units per meal. Blood glucose monitoring is irregular due to his demanding farm work schedule, and he reports checking his levels only about half the time before operating machinery or driving. Of particular concern is his development of hypoglycemic unawareness, evidenced by a recent episode where he discovered a blood glucose of 2 millimoles per liter without symptoms. While he keeps "lollies" available for treating hypoglycemia, the combination of irregular monitoring and impaired awareness poses significant safety concerns given his occupation and driving requirements.

His chronic kidney disease has progressed to stage 4, with a recent estimated glomerular filtration rate of 18. He is currently managed with Ramipril 5 milligrams daily for renoprotection, though the effectiveness of this intervention at his current stage of kidney disease warrants review. While he denies current symptoms of fluid overload or uremic symptoms, his condition is approaching the need for renal replacement therapy. The distance to dialysis facilities presents a significant logistical challenge for future management. His brother has been identified as a potential living kidney donor, though formal evaluation has not yet commenced. This advance planning for renal replacement therapy is crucial given his young age and significant responsibilities.

Mr. Mitchell's mental health has deteriorated in recent months, presenting a complex interaction with his physical health management. He was diagnosed with post-traumatic stress disorder following a severe farm accident six years ago, for which he takes Sertraline 50 milligrams daily. His depression has worsened over the past three months, manifesting as low mood, anhedonia, and increased alcohol consumption of 5-6 standard drinks most days. While he acknowledges having thoughts about his car in relation to self-harm, he firmly states he would not act on these thoughts. His mood deterioration appears closely linked to concerns about potential driving restrictions and their impact on his ability to maintain his livelihood. Despite these significant mental health challenges, he is not currently engaged in regular psychotherapy or counseling, largely due to accessibility issues in his rural location.

Diabetic complications are evident across multiple systems, reflecting the long duration and suboptimal control of his diabetes. He has progressive diabetic retinopathy requiring urgent ophthalmology review, though his last assessment was 12 months ago. Current visual acuity is 6/12 in the right eye and 6/9 in the left eye with correction. Fundoscopic examination reveals proliferative changes including microaneurysms, dot-blot hemorrhages, and neovascularization, particularly in the right eye. He demonstrates peripheral neuropathy with reduced sensation to his ankles, though he reports no difficulty operating vehicle pedals. This combination of retinopathy and peripheral neuropathy raises significant concerns about his ability to safely continue driving and operating farm machinery. His cardiovascular risk is managed with Atorvastatin 20 milligrams nightly, though he has not had a recent comprehensive cardiovascular risk assessment.

Regarding preventive health measures, his engagement with routine health maintenance has been limited. While he is up to date with COVID-19 vaccination, he has not received recent influenza or pneumococcal vaccines, which are particularly important given his chronic kidney disease. He reports infrequent self-inspection of his feet and has not attended a diabetic foot clinic, despite the presence of peripheral neuropathy. His last comprehensive health check was several years ago, indicating a need for improved preventive care coordination.

Mr. Mitchell's social circumstances significantly impact his health management. He lives on a small farm in rural South Australia with his wife and young child, where he is the primary income earner through farming activities involving crops and livestock. His role requires regular driving for both farm operations and medical appointments, with the nearest dialysis center being a considerable distance away. This presents significant challenges for future renal replacement therapy planning. His wife and young child rely heavily on his income and day-to-day farm operations, creating additional pressure regarding potential driving restrictions. While neighbors are reportedly not far away, he lacks a formal support network for managing medical emergencies on the farm, with his wife being his primary emergency contact.

Physical examination reveals a sun-weathered, lean-built man appearing tired and tense. His vital signs show a blood pressure of 138/88 millimeters of mercury, heart rate of 88 beats per minute, respiratory rate of 16 breaths per minute, and oxygen saturation of 98% on room air. His temperature is 36.5 degrees Celsius. His anthropometric measurements show a height of 178 centimeters, weight of 75 kilograms, resulting in a body mass index of 23.7, and a waist circumference of 88 centimeters.

Cardiovascular examination reveals no jugular venous pressure distension, with normal apex beat position and heart sounds, without murmurs or additional sounds. Respiratory examination demonstrates normal chest expansion and clear breath sounds throughout. Abdominal examination is unremarkable aside from minor scarring from previous farm incidents, with normal bowel sounds and no organomegaly or tenderness.

Neurological examination shows grossly intact cranial nerves but reveals significant peripheral neuropathy with reduced sensation to light touch and vibration in his feet, accompanied by diminished ankle reflexes. While no active foot ulcers are present, there is some callus formation. Motor strength remains preserved at 5/5 throughout. Eye examination reveals visual acuity of 6/12 in the right eye and 6/9 in the left eye with correction. Fundoscopy demonstrates proliferative diabetic retinopathy with microaneurysms, dot-blot hemorrhages, and neovascularization, particularly affecting the right eye. While there is no current evidence of vitreous hemorrhage, the proliferative changes indicate high risk for this complication.

The complexity of Mr. Mitchell's case is further complicated by the absence of advance care planning. He has neither an Advanced Care Directive nor Enduring Power of Attorney in place, and it is uncertain whether he has a formal will. Given his young family and significant health challenges, this represents an important gap in his comprehensive care planning.

In summary, Mr. Mitchell is a 45-year-old farmer with advanced chronic kidney disease and poorly controlled Type 1 diabetes mellitus, whose ability to maintain his livelihood is threatened by potential driving restrictions. The five key issues I would like to focus on addressing are:

1. Urgent optimization of diabetes management, particularly addressing hypoglycemic unawareness and implementing a structured monitoring protocol before driving and operating machinery. This includes revising his insulin regimen, implementing more frequent blood glucose monitoring, and establishing clear protocols for treating hypoglycemia.

2. Chronic kidney disease progression requiring immediate planning for renal replacement therapy, including evaluation of his brother as a potential living donor. This planning must consider the logistical challenges posed by his rural location and the potential impact on his ability to continue farming.

3. Management of diabetic complications, particularly urgent ophthalmology review for progressive retinopathy and establishment of regular diabetic foot care. This includes addressing the implications of these complications for his driving safety and developing appropriate risk mitigation strategies.

4. Mental health deterioration and alcohol use, requiring integrated psychological support and substance use management. This needs to account for his rural location and may require consideration of telehealth options for regular psychological support.

5. Development of a comprehensive support plan to maintain his ability to work safely while managing his medical conditions. This includes involvement of his wife in care planning, establishment of local emergency support networks, and addressing advance care planning needs. The plan must balance the competing demands of preserving his livelihood while ensuring his safety and the safety of others.

### Supplementary Information 5 – Question answering

#### Case 1 – Cystic fibrosis and lung transplant

##### Examiner 1

**Examiner:** Acknowledging there is uncertainty with respect to then aetiology of his current respiratory decline, what do you think this patient’s overall prognosis is?

**Doctor:** Based on the concerning combination of findings, I believe Mr. Smith's prognosis is guarded to poor in the medium term. The 50% decline in his FEV1 from baseline, coupled with significantly reduced exercise tolerance and recent multi-resistant Pseudomonas infection, strongly suggests chronic lung allograft dysfunction. This condition typically follows a progressive course, with median survival around 3-5 years after onset. His prognosis is further compromised by several compounding factors: ongoing substance use that may affect his immunosuppression, poor nutritional status with recent weight loss, and suboptimally controlled diabetes.

However, I want to emphasize that prognosis can be positively influenced if we can optimize several factors. If we can rule out other reversible causes of his decline, such as acute infection or medication non-adherence, and if he responds to intensified immunosuppression for chronic rejection, his lung function might stabilize. Additionally, addressing his nutritional status and substance use could significantly impact his outcomes.

Given this outlook and his strong desire to see his terminally ill father, I believe we need to act with appropriate urgency while still being thorough in our workup. My immediate priority would be to expedite his transplant team review while simultaneously working to optimize his condition for safe travel. The window of opportunity for him to achieve his goal of seeing his father may be limited, and this should factor into our medical decision-making and risk assessments.

**Examiner:** If this was chronic rejection and his lung function continues to decline, do you think this patient could be, or become, an appropriate candidate for repeat lung transplantation?

**Doctor:** Based on careful consideration of Mr. Smith's current clinical picture, I would not support listing him for repeat lung transplantation at this time. The combination of active substance use, particularly intravenous methamphetamine use as recently as three weeks ago, along with his significant psychosocial challenges including limited support network and financial instability, presents substantial contraindications to retransplantation.

Moreover, his declining nutritional status with a BMI of 17.9-18.0 and ongoing weight loss would need to be significantly improved before he could be considered a viable candidate. His diabetes management also requires optimization, given his consistently elevated blood glucose levels.

That said, I believe we could potentially reconsider his candidacy for retransplantation if several key issues are addressed. This would require demonstrated sustained abstinence from substance use with appropriate support programs in place, improvement in his nutritional status, optimization of his diabetes management, and establishment of a more robust social support system. Additionally, we would need to ensure stable financial resources to support post-transplant care.

However, given the urgency of his desire to see his terminally ill father and the complexity of achieving these optimizations, I think our immediate focus should be on stabilizing his current condition and facilitating safe travel rather than pursuing the lengthy evaluation process for retransplantation. This approach aligns better with his expressed priorities while acknowledging the realistic limitations of his current situation.

**Examiner:** Specifically, how would you approach his methamphetamine use currently?

**Doctor:** Given Mr. Smith's complex medical situation and his expressed priority of seeing his terminally ill father, I would take a pragmatic, harm-reduction approach to his methamphetamine use rather than making abstinence an immediate requirement. I would have a frank discussion with him about how substance use increases his risk of infection and could potentially interact with his immunosuppression, directly impacting his ability to achieve his goal of traveling to see his father.

I would focus on establishing regular screenings and safety protocols, including more frequent monitoring of his tacrolimus levels and liver function, and implementing a clear action plan for seeking medical attention if he develops fever or other concerning symptoms after use. I'd also work with him to establish a regular schedule for substance screening, making this a routine part of his care rather than a punitive measure.

While I would strongly encourage him to consider addiction support services, I recognize that he's explicitly stated this isn't his primary concern right now. Instead, I would focus on building trust and maintaining open communication about his use patterns. I would provide him with clean needle supplies and education about safer injection practices to minimize infection risks, particularly given his immunosuppressed status.

Most importantly, I would document our discussions and management plan clearly but separately from his transplant records, to avoid potentially compromising his future candidacy for retransplantation if he achieves stability and abstinence later. This approach balances medical reality with his current priorities while keeping future options open.

**Examiner:** Can you please provide details on any further workup that you think is required with respect to his nutrition and weight loss?

**Doctor:** I would initiate a comprehensive nutritional assessment focusing on both the underlying causes and potential interventions for Mr. Smith's concerning weight loss, with his BMI dropping from 19 to 17.9 over the past year. First, I would order a complete set of laboratory studies including albumin, prealbumin, vitamins A, D, E, and K levels, zinc, iron studies, and B12 to establish his current nutritional status and guide supplementation.

Given his cystic fibrosis, I would evaluate the adequacy of his current pancreatic enzyme replacement by measuring fecal elastase and conducting a 72-hour fecal fat collection. This would help determine if his current pancrelipase dosing of 40,000 units with meals is sufficient. I would also check his tacrolimus levels, as high levels can contribute to gastrointestinal symptoms and poor absorption.

To better understand his dietary intake, I would have him complete a detailed 7-day food diary, including timing of pancreatic enzyme administration relative to meals. This would help identify any patterns of inadequate intake or enzyme timing issues that might be contributing to malabsorption. Additionally, given his diabetes, I would review his glycemic control, as poor control can contribute to weight loss through glucosuria and protein catabolism.

The goal would be to develop a comprehensive nutritional plan that addresses both the medical aspects of his malabsorption and the practical challenges of maintaining adequate caloric intake, particularly as he prepares for potential international travel to see his father.

**Examiner:** Do you think that he should try to go and visit his father overseas?

**Doctor:** Yes, I believe Mr. Smith should attempt to visit his father in Spain, but with careful planning and specific medical safeguards in place. The emotional significance of seeing his terminally ill father outweighs the manageable medical risks of international travel. His current condition, while concerning with an FEV1 of 50% and exercise limitation, remains stable enough that travel could be undertaken safely with proper preparation.

I would work to expedite several key medical preparations. First, I would arrange for him to travel with an adequate supply of all medications, particularly his immunosuppressants and insulin, accounting for time zone adjustments. I would provide him with a detailed medication schedule and help him set up a phone-based tracking system. Second, I would prepare comprehensive medical documentation in both English and Spanish, including his transplant history, current medication list, and recent culture results showing multi-resistant Pseudomonas.

Additionally, I would coordinate with his transplant team to identify a suitable medical center in Spain near his father's location for emergency care if needed. We would need to ensure his travel insurance covers his pre-existing conditions and arrange for supplemental oxygen during the flight given his exercise limitation and baseline oxygen saturation of 94%.

While his current substance use and financial constraints present challenges, these should not preclude this potentially final opportunity to see his father. Instead, we should focus on minimizing risks through careful planning while acknowledging that waiting for perfect medical optimization might mean missing this crucial family connection.

##### Examiner 2

**Examiner:** Let's discuss the first issue of progressive dyspnoea. Can you talk me through what top differentials you are considering and how you would investigate this further?

**Doctor:** The progressive dyspnea in Mr. Smith is most concerning for chronic lung allograft dysfunction, particularly given the significant decline in FEV1 from 65% to 50% over a relatively short period. This decline, coupled with his recent hospitalization for multi-resistant Pseudomonas infection, suggests a potentially serious deterioration in his transplant function.

I would immediately arrange spirometry with flow-volume loops and diffusion capacity testing to quantify his current lung function and compare it to his baseline values. Additionally, I would order high-resolution CT imaging of his chest to look for bronchial wall thickening, air trapping, and other features suggestive of bronchiolitis obliterans syndrome, the most common form of chronic rejection.

Given his recent infection, I would also obtain sputum cultures to assess for ongoing infection or colonization. Blood tests would include tacrolimus levels to ensure therapeutic immunosuppression, along with inflammatory markers and a complete blood count. I would perform a thorough cardiac assessment including echocardiography to rule out any cardiac contribution to his symptoms, particularly given the potential cardiotoxicity of some of his medications.

These investigations need to be expedited given his desire to travel internationally. If chronic rejection is confirmed, we would need to consider intensifying his immunosuppression, which could significantly impact the safety and timing of his planned travel to see his father.

**Examiner:** If this is chronic rejection, what options are there to manage this moving forward?

**Doctor:** Looking at Mr. Smith's declining lung function and clinical picture, I would pursue a multi-faceted approach to managing his likely chronic rejection. Given his already significant functional limitation and the emotional urgency of seeing his terminally ill father, we need to act quickly but thoughtfully.

I would first augment his immunosuppression by increasing his tacrolimus target levels and implementing a pulse of high-dose methylprednisolone, followed by a temporary increase in his maintenance prednisolone. Simultaneously, I would add azithromycin at an immunomodulatory dose - though I note he's already on this medication, so we should optimize the dosing schedule.

Given his recent multi-resistant Pseudomonas infection, I would also initiate targeted antimicrobial therapy based on sensitivity patterns. This is crucial as infection can accelerate the rejection process. Additionally, I would consider implementing extracorporeal photopheresis, which has shown promise in managing chronic rejection, though this would need to be balanced against his travel plans and financial constraints.

If these interventions don't stabilize his lung function within 4-6 weeks, we would need to have a difficult discussion about re-transplantation. However, his ongoing substance use would need to be addressed before this could be considered. In the immediate term, my priority would be stabilizing his condition sufficiently to allow him to safely travel to see his father, while being transparent about the challenges and prognosis of chronic rejection.

**Examiner:** What could be contributing to this patient's weight loss, and how will you optimise his nutrition moving forward?

**Doctor:** Mr. Smith's weight loss is likely multifactorial, stemming from his increased work of breathing, cystic fibrosis-related malabsorption, and potentially suboptimal caloric intake due to his declining health and psychosocial stressors. The decline in BMI from 19 to 17.9 over the past year is particularly concerning given his transplant status and could negatively impact his prognosis.

I would optimize his nutrition through several targeted interventions. First, I would adjust his pancreatic enzyme replacement therapy, potentially increasing the dose of pancrelipase and redistributing it more effectively around his meals and snacks. I would also implement a high-calorie, high-protein diet with specific attention to timing around his medications and insulin dosing to maximize absorption.

Given his significant energy expenditure from increased work of breathing, I would calculate his specific caloric needs and implement supplementary nutrition, likely in the form of specialized oral supplements between meals. Regular dietitian follow-up would be essential to monitor his progress and adjust interventions as needed.

Additionally, I would assess and optimize his blood glucose control, as poor glycemic control can contribute to weight loss and malabsorption. This comprehensive approach should help stabilize his weight, which is crucial for maintaining his overall health and supporting his ability to travel to see his father.

**Examiner:** What advice would you give this patient about international travel? How would you mitigate any health risks this might pose?

**Doctor:** Given Mr. Smith's complex medical status and his compelling need to see his terminally ill father, I would support his travel plans while implementing specific safeguards. I recommend he undertake the journey within the next few weeks, before any potential further deterioration in his condition, but only after we've initiated treatment for his likely chronic rejection.

The key measures I would implement include providing him with a detailed medical summary and contact information for his transplant team, along with securing a connection to a transplant center in Spain near his father's location. I would ensure he has a 30-day supply of all medications, with particular attention to his insulin and immunosuppressants, and create a detailed schedule accounting for time zone changes. His tacrolimus levels should be checked immediately before travel and a monitoring plan established for his time abroad.

For infection prevention, I would prescribe prophylactic antibiotics based on his recent Pseudomonas sensitivities and ensure he has masks and hand sanitizer for the journey. I would also recommend business class travel if financially feasible to allow more space and reduce infection risk, though I acknowledge his financial constraints may make this challenging.

Most importantly, I would work with him to develop clear criteria for seeking urgent medical attention and ensure he has travel insurance that covers his pre-existing conditions. This balanced approach should allow him to achieve his goal of seeing his father while maintaining his health as much as possible.

**Examiner:** What is your overall impression of this gentleman's prognosis, and to what extent do you think he is aware of it?

**Doctor:** Based on my assessment, Mr. Smith's prognosis is guarded and likely to be measured in months rather than years, given the constellation of concerning findings. The significant decline in his lung function, recent multi-resistant Pseudomonas infection, ongoing substance use, and deteriorating nutritional status all suggest a critical downturn in his transplant function that may be difficult to reverse.

I believe Mr. Smith has partial insight into his situation, evidenced by his urgency to see his father and his acknowledgment of worsening symptoms. However, his focus on financial and logistical concerns rather than his declining health suggests he may not fully grasp the severity of his current trajectory. His comment that his substance use "isn't the biggest issue" also indicates he may not fully appreciate how this compounds his already precarious health status.

When discussing prognosis with Mr. Smith, I would be direct but compassionate, emphasizing that his current decline represents a serious change in his condition that requires immediate attention. I would frame this discussion within the context of his stated priority of seeing his father, explaining that addressing his health deterioration promptly is crucial for making this trip possible. This approach allows us to maintain hope while ensuring he understands the urgency of his situation and the importance of engaging fully with the proposed interventions.

#### Case 2 – Heart Failure with reduced Ejection Fraction and falls

##### Examiner 1

**Examiner:** Please outline how you would approach the optimisation of her heart failure and how you would determine goals of care and success? Any anticipated difficulties in this?

**Doctor:** In optimizing Mrs. Ng's heart failure management, I would prioritize keeping her at home while improving her quality of life, given her clear wishes to avoid hospitalization and maintain independence. She's already on comprehensive guideline-directed medical therapy, including optimal doses of Entresto, beta-blockade, and mineralocorticoid receptor antagonist, so my focus would be on managing her fluid status and symptoms.

I would implement daily weight monitoring and adjust her diuretic regimen proactively based on weight changes, teaching her to increase her furosemide dose temporarily when she notices early signs of fluid retention. This approach would help prevent the acute decompensations that have led to her recent hospitalizations. However, I anticipate challenges with this strategy given her intermittent use of scales and reluctance to engage with medical interventions.

For goals of care, I would define success primarily by her ability to maintain independence in her garden rather than traditional metrics like ejection fraction or BNP levels. Specific markers would include reduced hospital admissions, stable weight, improved exercise tolerance beyond her current 10-meter limitation, and maintained independence with activities of daily living. I would also track her symptom burden, particularly orthopnea and exertional dyspnea.

The main difficulty I foresee is balancing optimal medical management with her strong preference for minimal intervention. Her recent weight loss and limited diet also complicate diuretic management. I would involve palliative care early to help manage symptoms and support her goals of staying at home, while ensuring we're not undertreating her heart failure in a way that could paradoxically threaten her independence.

**Examiner:** How would you investigate her recent weight loss and are there any considerations other than purely nutritional support?

**Doctor:** I would take a systematic approach to investigating Mrs. Ng's 5kg weight loss, considering both cardiac and non-cardiac causes. Given her advanced heart failure, I'm concerned this weight loss could represent cardiac cachexia, which would significantly impact her prognosis and functional status. The combination of her limited diet, predominantly tea and toast, with her increasing functional limitation suggests a cyclical relationship where fatigue and breathlessness may be limiting her ability to prepare and enjoy meals.

My initial investigation would include comprehensive blood work to assess for common reversible causes, including thyroid function, diabetes screening, celiac serology, and inflammatory markers. I would also check B12, folate, and iron studies given her limited diet. A chest X-ray and basic malignancy screening would be prudent given her age and smoking history.

However, beyond the standard nutritional workup, I'm particularly concerned about the psychosocial aspects of her weight loss. The loss of her husband four years ago, her limited social network, and her fierce desire to maintain independence may be contributing to both reduced meal preparation effort and reduced enjoyment of meals. Depression screening would be essential, as social isolation and chronic illness often impact mood and appetite.

I would propose starting with practical interventions that preserve her autonomy, such as pre-prepared meals or assistance with grocery shopping and meal preparation, framing these supports as ways to maintain her energy for gardening and other activities she values. This approach respects her priorities while addressing the nutritional concerns that could further compromise her independence.

**Examiner:** Take me through your recommendations and strategy to help prevent her from falling and what other complications of these might need investigation?

**Doctor:** Given Mrs. Ng's recent fall while gardening and her reluctance to use a walking aid, I would implement a multifaceted falls prevention strategy that prioritizes her independence while ensuring safety. Although she attributes her recent fall to tripping over her cat, I'm concerned about multiple contributing factors including her cardiac status, deconditioning, and possible frailty.

My primary intervention would be to arrange a home occupational therapy assessment, focusing specifically on her garden area since this is where she spends significant time and where her last fall occurred. I would frame this as supporting her gardening hobby rather than restricting it. I would also strongly recommend a four-wheeled walker, but rather than presenting it as a mobility aid, I would position it as a gardening tool that can carry her supplies and provide a ready seat for resting, aligning with her desire to continue gardening independently.

Regarding complications, I'm particularly concerned about her bleeding risk given she's on both aspirin and apixaban. Her examination revealed bruising on her forearms, suggesting she may be at increased risk of trauma from falls. I would carefully weigh the benefits and risks of dual therapy, likely discontinuing the aspirin while maintaining the apixaban for her atrial fibrillation, given her high stroke risk.

I would also assess her bone health, as her limited diet and reduced mobility put her at high risk for osteoporosis. I would arrange bone mineral density testing and consider osteoporosis prophylaxis, though careful timing and choice of therapy would be needed given her reluctance to engage with medical interventions.

**Examiner:** How would you tackle the important issue of discussing an advanced care directive with this patient, are there any other stakeholders you would involve?

**Doctor:** I would approach advance care planning with Mrs. Ng in a stepwise manner, starting by exploring her understanding of her heart failure trajectory and what matters most to her. From our discussions, it's clear that maintaining independence and staying in her garden are her primary goals. I would use this as a foundation for the conversation, acknowledging that documenting her wishes could help ensure her autonomy is respected.

Given her recent hospitalizations and her expressed desire to avoid further hospital admissions, I would frame advance care planning as a way to ensure her healthcare team understands and honors her preferences. I would explore specific scenarios she might face with advanced heart failure, such as whether she would want hospital admission if her breathing worsened significantly, or if she would prefer symptom management at home with appropriate support.

Regarding stakeholders, while Mrs. Ng has no immediate family, I would encourage her to identify friends or neighbors who could potentially act as her medical power of attorney. The palliative care team would be essential partners in this discussion, as they can provide expertise in both symptom management and advance care planning. I would also involve her GP and cardiology team to ensure alignment of care across all providers.

Most importantly, I would ensure this conversation happens over several sessions, allowing her time to process and refine her decisions. This approach respects her clear need for control while ensuring her wishes are properly documented and communicated to all relevant healthcare providers.

**Examiner:** What immediate priorities do you have in terms of maintaining her independence and optimising her ability to remain at home?

**Doctor:** I would prioritize establishing a sustainable support system that preserves Mrs. Ng's autonomy while reducing her risk of further hospitalizations. Given her recent admissions and increasing difficulty with daily activities, I would start by implementing cleaning assistance, which she has already indicated she would accept. This would conserve her energy for activities she values, particularly gardening.

For her heart failure management, I would focus on early detection of fluid overload by providing her with a clear action plan. This would include teaching her to recognize early warning signs and empowering her to adjust her diuretic dose within prescribed parameters. I would arrange regular home visits from a heart failure nurse to monitor her status, framing these visits as supporting her independence rather than imposing care.

Regarding her nutrition, I would organize meal delivery services or assistance with grocery shopping and meal preparation, as her current limited diet puts her at risk of further functional decline. This support would need to be introduced gradually, respecting her desire for control while ensuring adequate nutrition to maintain her strength and independence.

I would also implement a medication review, particularly investigating the indication for pregabalin, as polypharmacy may be contributing to her fall risk. Simplifying her medication regimen where possible could help maintain her excellent compliance while reducing potential adverse effects that might compromise her independence.

These interventions would be coordinated with palliative care involvement, focusing on optimizing her quality of life and supporting her clear wish to remain in her home environment.

##### Examiner 2

**Examiner:** Issues regarding independence in a Lady whom perceives external input as a threat to her independence. How and what falls prevention strategies do you think she would consider, and what do you think would be most beneficial for her?

**Doctor:** Given Mrs. Ng's strong desire for independence and her perception of assistive devices as a threat to her autonomy, I would approach falls prevention through the lens of enabling her to continue gardening safely, which is clearly a central source of meaning in her life.

I would start by addressing the most immediate modifiable risk factors while being mindful not to overwhelm her with changes. My priority would be working with her to identify specific high-risk activities in her garden where she's most likely to experience difficulties. For instance, I noticed she mentioned pruning as an activity she still enjoys. I would suggest creating designated rest stations in her garden - perhaps attractive garden benches strategically placed along her usual gardening routes. This approach frames the intervention as enhancing her gardening experience rather than restricting her independence.

For the four-wheeled walker she's currently resistant to, I would reframe this as a gardening aid - perhaps suggesting a modified version that could hold her gardening tools and serve as a mobile rest station. This reframing from "medical device" to "gardening assistant" might make it more acceptable to her while still providing the needed support for safe mobility.

Rather than impose a comprehensive falls prevention program, I would prioritize these practical modifications that directly support her valued activities. This approach respects her autonomy while incrementally introducing safety measures that she's more likely to accept because they align with her goals of maintaining her garden and her independence.

**Examiner:** What can you do to stop her from presenting to the hospital given this is her biggest issue?

**Doctor:** Given Mrs. Ng's multiple recent hospitalizations and her strong desire to avoid future admissions, I would implement a proactive outpatient management strategy focused on early intervention and symptom control. The key is to identify and treat decompensation before it reaches a critical point requiring hospitalization.

I would arrange for weekly home visits initially, transitioning to fortnightly as she stabilizes, focusing on careful titration of her diuretic regime based on symptoms and examination findings. Given her demonstrated compliance with fluid restriction and medications, I would provide her with a flexible furosemide protocol - allowing her to increase her dose from 80mg to 120mg twice daily for up to three days if she notices increasing breathlessness or ankle swelling. This empowers her with some control over her condition while maintaining medical oversight.

I would also establish clear clinical triggers for early outpatient review rather than emergency presentation. For example, if she needs to increase her furosemide for more than three days, or if she gains more than 2kg over a week, she should contact us for an urgent review rather than waiting until she requires emergency care. To facilitate this, I would ensure she has reliable scales at home and teach her to monitor her weight daily.

Additionally, I would explore involving palliative care services, not for end-of-life care, but for their expertise in symptom management and supporting patients to remain at home. Their involvement could provide an additional layer of support and expertise in managing her symptoms proactively, aligning with her goal of avoiding hospitalizations while maintaining her independence at home.

**Examiner:** Regarding your second issue, what do you perceive as the cause of her weight loss, and what differentials would you consider?

**Doctor:** I see Mrs. Ng's weight loss as primarily arising from her advanced heart failure causing cardiac cachexia, evidenced by her severely reduced ejection fraction of 20%, marked functional limitation, and recent frequent hospitalizations. The physiological stress of her condition, combined with elevated inflammatory markers typically seen in advanced heart failure, often leads to progressive muscle wasting and weight loss despite apparently stable intake.

However, I must also consider other contributing factors, particularly depression given her social isolation and loss of independence. Her limited diet of predominantly tea, toast, and cheese sandwiches suggests possible reduced appetite which could be a manifestation of depression, especially considering her husband's death four years ago and her current social situation.

Given her age and unintentional weight loss, malignancy must be considered as a differential diagnosis. Her smoking history particularly raises concern for occult lung cancer. Additionally, her medication burden could be contributing through side effects - notably, the high doses of furosemide might be affecting her appetite and causing early satiety.

Thyroid dysfunction should also be considered, as it can both exacerbate heart failure and cause weight loss. Given her limited diet, nutritional deficiencies could be compounding the problem. Specifically, her predominantly bread-based diet raises concern for inadequate protein intake, which could accelerate muscle wasting in the context of cardiac cachexia.

I would prioritize investigating cardiac cachexia while simultaneously screening for depression and malignancy, as identifying the correct underlying cause will guide our intervention strategy.

**Examiner:** She is not currently treated for her COPD. What is your approach to the treatment of COPD, and how would you implement these in this patient?

**Doctor:** While Mrs. Ng has diagnosed COPD, I would carefully balance the initiation of COPD therapy against her other conditions and priorities. Her symptoms appear predominantly related to her heart failure rather than COPD, evidenced by her response to diuretics during hospital admissions and her report that salbutamol provides minimal relief. Additionally, her primary concern is maintaining independence at home, and adding complexity to her medication regime needs to be weighed against this goal.

Nevertheless, I would initiate a dual bronchodilator therapy with a long-acting muscarinic antagonist (LAMA) and long-acting beta-agonist (LABA) combination inhaler. This would provide maximal bronchodilation with minimal medication burden, as it requires only one device and once-daily dosing. I would avoid inhaled corticosteroids initially, given her advanced heart failure and the risk of fluid retention.

However, before starting this treatment, I would ensure proper inhaler technique through demonstration and teach-back, as poor technique could negate any potential benefits. I would also explain that while this treatment may help her breathing, her heart failure remains the primary driver of her symptoms. This sets realistic expectations and helps maintain trust in our therapeutic relationship.

Regarding smoking cessation, while I would typically prioritize this in COPD management, I recognize her clear stance on continuing to smoke after 50 years. Rather than risk damaging our therapeutic alliance by pushing cessation, I would focus our efforts on optimizing her symptom management through other means.

**Examiner:** She notes an ongoing history of smoking. Would you recommend cessation in this 85-year-old female. If so, what strategies would you implement?

**Doctor:** In Mrs. Ng's case, I would not actively pursue smoking cessation at this time. While smoking cessation typically offers significant health benefits, I need to carefully consider her overall situation and priorities. At 85 years old, with advanced heart failure, recent weight loss, and a clear focus on maintaining quality of life and independence, aggressive smoking cessation efforts could cause unnecessary distress without meaningful benefit to her daily functioning or life goals.

Mrs. Ng has explicitly stated she doesn't plan to quit after 50 years of smoking, and pushing this issue risks damaging our therapeutic relationship at a crucial time when we need her trust to manage her heart failure and prevent hospitalizations. The stress of quitting could potentially worsen her quality of life in her remaining time, particularly given smoking likely serves as a coping mechanism and source of comfort for her.

Instead, I would focus our therapeutic efforts on optimizing her heart failure management and supporting her independence at home. If she were to express interest in cutting down or quitting, I would certainly support this with nicotine replacement therapy and counseling. However, initiating this conversation without her interest could undermine her sense of autonomy, which she deeply values, and potentially make her more resistant to other medical interventions that could provide more immediate benefit to her quality of life.

In essence, while smoking cessation would typically be indicated, the potential benefits need to be weighed against her limited life expectancy, expressed preferences, and our need to maintain a strong therapeutic alliance to achieve her primary goal of staying independent at home.

#### Case 3 – Myasthenia gravis and osteoporosis

##### Examiner 1

**Examiner:** How well controlled do you think her myasthenia gravis is at the moment and what could you do to try and improve it?

**Doctor:** Based on my evaluation today, Mrs Brown's myasthenia gravis is suboptimally controlled. She's experiencing ongoing symptoms including intermittent fatigue, lower limb weakness that's contributed to recent falls, and swallowing difficulties. These symptoms notably worsen when attempting to reduce her prednisolone, suggesting we haven't achieved stable disease control with her current immunosuppression.

I would optimize her treatment in two ways. First, I would increase her mycophenolate dose from 500mg twice daily to 1000mg twice daily, as we haven't reached the typical therapeutic dose for myasthenia gravis. While this increases immunosuppression, she's already on appropriate prophylaxis with trimethoprim-sulfamethoxazole. Second, I would maintain her current prednisolone dose of 25mg daily for another month until the increased mycophenolate takes effect, rather than attempting reduction now. Once we achieve better symptom control, we can initiate a slower prednisolone taper, perhaps reducing by 2.5mg every 4-6 weeks.

I'm particularly motivated to improve her disease control because her muscle weakness is significantly impacting her quality of life and contributing to falls risk. Better control of her myasthenia gravis would help her achieve her goal of visiting her sister interstate and returning to gardening. While increasing immunosuppression carries risks, I believe the benefits of improved symptom control and enhanced function outweigh these risks, especially given her current prophylaxis and up-to-date vaccinations.

**Examiner:** What strategies can be implemented to reduce her risk of falls?

**Doctor:** I would implement a comprehensive falls prevention strategy focusing on three key areas. First, addressing her muscle weakness through a tailored exercise program, as this is a major contributor to her falls. I would start her on a gentle strengthening program focusing on proximal leg muscles and core stability, carefully designed around her myasthenia gravis symptoms. The exercises would need to be performed when her medication effect is maximal, with appropriate rest periods to prevent fatigue.

Second, optimizing her pain management is crucial, as her back pain is significantly affecting her mobility and movement patterns. While maintaining her current analgesic regimen, I would add targeted physiotherapy including gentle stretching and pain management techniques. This would help improve her movement confidence and reduce the guarding behavior I observed during her examination.

Third, I would focus on environmental modifications and practical strategies. Given that she lives in a single-story home, we're fortunate to avoid stairs, but I would recommend installing grab rails in the bathroom and ensuring adequate lighting throughout the house. I would also work with her on strategies for safely managing daily activities, particularly during periods of fatigue. While she currently doesn't use a walking aid, I would recommend a walking stick for times when her legs feel particularly weak or when she's more fatigued.

The goal is to enhance her safety while supporting her independence, particularly as she works toward her aim of interstate travel to visit her sister.

**Examiner:** How would you go about treating her depression?

**Doctor:** Mrs Brown's depression requires careful attention, particularly given its long duration and the current impact of her physical health challenges on her mental wellbeing. I would optimize her treatment through both pharmacological and non-pharmacological approaches.

On the medication front, while she's currently taking duloxetine 60mg daily for both depression and pain, I'm concerned that this dual-purpose approach might be compromising the effectiveness of her depression treatment. I would increase her duloxetine to 90mg daily, as this higher dose often provides better antidepressant effects while still managing pain. The timing is appropriate for dose escalation given her ongoing low mood and social withdrawal.

For non-pharmacological management, I would incorporate psychological support into her regular care. While she's had occasional psychology input in the past, I would implement regular sessions focusing on adjusting to her changing physical capabilities and maintaining social connections despite her limitations. Cognitive behavioral therapy would be particularly valuable in helping her develop strategies to stay engaged in meaningful activities despite pain and fatigue.

I'm particularly motivated to address her depression now because it's creating a barrier to achieving her personal goals, especially maintaining social connections and visiting her sister interstate. Moreover, her depression is likely exacerbating her perception of pain and fatigue, creating a cycle that's impacting her quality of life. By actively treating her depression, we can help break this cycle and improve her overall function.

**Examiner:** Do you think her current living situation is sustainable and what could be done to further support her?

**Doctor:** Based on my assessment, Mrs Brown's current living situation raises significant concerns about sustainability. While her husband is providing essential support with cooking, cleaning, and shopping, the physical and emotional burden on him is likely to increase as her conditions progress. Her recent falls, ongoing fatigue, and significant pain suggest that relying solely on informal support from her husband may become inadequate.

I would implement a tiered approach to enhance her support system. Initially, I would arrange for an occupational therapy home assessment to identify immediate modifications that could improve her safety and independence. This would include bathroom modifications and assistive devices for daily activities. Furthermore, I would organize regular in-home physiotherapy sessions to help maintain her mobility and strength within her own environment.

For longer-term sustainability, I would establish formal support services through the community care system. This would include regular homecare assistance for personal care and domestic tasks, initially perhaps twice weekly. This gradual introduction of formal support would help preserve her husband's energy for companionship and emotional support, while ensuring professional assistance is available for more physically demanding tasks.

These supports are particularly important given her goal of interstate travel. By establishing a robust support system at home, we can help her maintain the strength and confidence needed for travel, while ensuring her husband isn't overwhelmed by caregiving responsibilities. This balanced approach would help sustain their current living arrangement while supporting their quality of life as a couple.

**Examiner:** What targets would you like this patient to achieve before she tries to go and visit her sister interstate?

**Doctor:** Before Mrs Brown undertakes interstate travel, I would want to achieve several key targets to ensure her safety and comfort. First, her myasthenia gravis symptoms need to be better controlled, specifically maintaining stable swallowing function and limb strength for at least six weeks without significant fluctuations. This stability would give us confidence that she could manage the physical demands of travel.

Second, we need to establish effective pain management that allows her to sit comfortably for extended periods and sleep adequately. While complete pain elimination may not be realistic, I would aim for pain levels that she rates as manageable and that don't significantly interfere with her daily activities. This would make the journey more feasible and enjoyable.

Third, I want to see improvement in her functional capacity, particularly her ability to walk short distances without significant fatigue. As a practical target, she should be able to walk 100 meters consistently without requiring rest, as this would be essential for navigating airports and other travel-related activities. Additionally, she should demonstrate stable balance with no falls for at least two months.

Finally, I would want her to trial a structured daily routine that mirrors the energy demands of travel, including activities outside her home environment. This would help build her confidence and provide a realistic assessment of her readiness for interstate travel. These targets focus on enabling her to achieve her goal while ensuring her safety and comfort during the journey.

##### Examiner 2

**Examiner:** What are the options for getting this patient off prednisolone whilst managing her MG symptoms?

**Doctor:** I would implement a careful steroid-sparing strategy for Mrs Brown, focusing on optimizing her mycophenolate dosing as the primary approach. Currently, she's only on 500mg twice daily of mycophenolate, which is suboptimal - I would increase this gradually to the target dose of 1000mg twice daily over 4-6 weeks while maintaining her current prednisolone dose. This ensures stable disease control during the transition.

Once she's been on the target mycophenolate dose for at least 8-12 weeks, allowing time for full therapeutic effect, I would begin a very gradual prednisolone taper. Given her history of symptom exacerbation with previous reduction attempts, I would reduce by just 2.5mg every 4 weeks, closely monitoring for signs of disease worsening like increased fatigue, limb weakness, or swallowing difficulties.

If she struggles with the mycophenolate increase or steroid reduction, I would consider adding pyridostigmine as a symptomatic therapy to help bridge the transition. While rituximab could be considered as an alternative steroid-sparing agent, I believe optimizing mycophenolate should be our first approach given its better safety profile and Mrs Brown's current complex medical situation with osteoporosis and falls risk. The goal would be to reach a prednisolone dose of 5-7.5mg daily over 6-8 months, which would help mitigate her osteoporosis risk while maintaining adequate disease control.

**Examiner:** What would be your approach to pain management?

**Doctor:** I would revise Mrs Brown's current pain management approach, as her ongoing severe back pain significantly impacts her function, sleep, and quality of life. The current regimen of four different pain medications - paracetamol, ibuprofen, tramadol, and duloxetine - creates a complex situation with potential risks, particularly given her immunosuppression and GORD.

My first step would be to discontinue the regular ibuprofen, as it poses unnecessary risks of gastric irritation and renal impairment. Instead, I would optimize her paracetamol timing to ensure consistent coverage, maintain the duloxetine which serves dual purpose for pain and depression, and adjust her tramadol dosing. Specifically, I would switch her from continuous release tramadol to immediate release formulation, allowing more flexible dosing around her activities and sleep.

For non-pharmacological management, I would implement a structured exercise program focusing on core strengthening and spinal stabilization, working within her fatigue limitations from myasthenia gravis. This would include specific sessions of activity followed by rest, gradually building up her tolerance. I would also address her sleep hygiene and incorporate relaxation techniques, as improved sleep quality often leads to better pain control.

The goal isn't just pain reduction but functional improvement - enabling her to work toward her goals of gardening and traveling to visit her sister. Regular monitoring of her pain levels and function will allow us to adjust this approach as needed while maintaining a careful balance with her other medical conditions.

**Examiner:** How can you prevent carer burnout and prevent further loss of independence by the patient?

**Doctor:** I would implement a two-pronged strategy addressing both immediate and long-term support needs for Mrs Brown and her husband. Given her husband is currently managing all household tasks while navigating her complex medical needs, there's a clear risk of carer burnout that needs immediate attention.

In the short term, I would arrange for home care services twice weekly to assist with cleaning and meal preparation. This provides immediate relief for her husband while maintaining their privacy and independence. I would also arrange for a personal alarm system given her recent falls, which would help alleviate her husband's constant concern about her safety.

For longer-term independence, I would focus on targeted rehabilitation within her medical constraints. Given her myasthenia gravis is still being optimized, I would start with short sessions of seated exercises and gradual mobility training that she can safely perform at home. As her strength improves, we can progress to more functional activities that directly support her goals, like gardening. This approach helps preserve her independence while preventing overexertion.

Additionally, I would connect them with a local carer support group and arrange for respite care one day per month, allowing her husband necessary breaks. This combination of practical support, rehabilitation, and respite care should help prevent carer burnout while supporting Mrs Brown's journey toward greater independence, ultimately working toward her goal of interstate travel to visit her sister.

**Examiner:** Is her osteoporosis optimally managed and are there other contributing factors that can be addressed?

**Doctor:** No, Mrs Brown's osteoporosis is not optimally managed, and there are several modifiable factors that require attention. Her current T-score of -2.8 at the lumbar spine, combined with a fragility fracture and ongoing high-dose glucocorticoid therapy, places her at very high risk for future fractures.

While she is appropriately on denosumab and calcium/vitamin D supplementation, I would add several key interventions. First, I would optimize her vitamin D levels through blood monitoring and dose adjustment, as adequate levels are crucial for both bone health and muscle strength. Second, I would address her nutrition by implementing a high-protein, calcium-rich diet plan that aligns with weight management goals, as her current diet of frequent takeaway meals is likely inadequate for bone health.

Her ongoing high-dose prednisolone use is a significant concern. While we need to balance this with her myasthenia gravis control, implementing the steroid-sparing strategy I mentioned earlier becomes even more critical for her bone health. Additionally, her limited mobility due to pain and weakness creates a vicious cycle of deconditioning that further impacts bone health.

I would implement a structured exercise program focusing on weight-bearing activities and resistance training, carefully tailored around her myasthenia gravis symptoms. This would address multiple goals: improving bone density, reducing fall risk, and supporting weight management. These interventions, combined with optimizing her current osteoporosis therapy, should help reduce her fracture risk while supporting her overall functional goals.

**Examiner:** How can you address her obesity, acknowledging the potential limitations of physical weakness and pain?

**Doctor:** I would approach Mrs Brown's obesity management primarily through dietary modification, with carefully planned physical activity as a secondary component. Given her BMI of 32.7 and the limitations imposed by myasthenia gravis and vertebral fracture pain, traditional exercise-based weight loss approaches would be challenging and potentially unsafe.

My primary focus would be on optimizing her nutrition through a sustainable dietary plan. Given her current reliance on takeaway meals, I would start with simple modifications like meal planning and preparation when her energy levels are highest, typically in the morning for most myasthenia gravis patients. I would target a modest caloric deficit of 500 calories daily, aiming for slow weight loss of 0.5kg per week. This conservative approach reduces the risk of muscle loss, which is particularly important given her existing weakness.

For physical activity, I would implement seated exercises and short walking intervals with rest periods, gradually increasing duration as tolerated. These would be scheduled between medication doses when her strength is optimal. The goal would be to improve her metabolic health while working within her physical limitations.

Weight loss would benefit multiple aspects of her health - reducing stress on her vertebral fracture, improving her mobility, and potentially allowing for better pain control. However, I would emphasize that this needs to be balanced against maintaining adequate nutrition for muscle strength and bone health, particularly given her immunosuppression and osteoporosis.

#### Case 4 – Perioperative and rural

##### Examiner 1

**Examiner:** What next steps would you take in perioperative risk optimisation here, targeting the most modifiable issues?

**Doctor:** Given Mr. Wilson's impending aneurysm surgery, I would focus on three key modifiable risk factors that we can realistically improve in a short timeframe. The most critical is smoking cessation, as his current 15 cigarettes per day significantly impacts his respiratory function and surgical risks. I would immediately start him on nicotine replacement therapy and varenicline, complemented by brief motivational interviewing during our sessions. This intervention could meaningfully improve his respiratory status even within a few weeks.

Secondly, his diabetes control needs rapid optimization given his HbA1c of 7.9% and concerning blood glucose range of 7-16 mmol/L. I would increase his gliclazide MR to 90mg daily and implement a structured blood glucose monitoring schedule, aiming for tighter perioperative glycemic control. This adjustment should help reduce his risk of postoperative complications and improve wound healing.

Thirdly, I would address his blood pressure, which is currently borderline at 140/88 mmHg. While he's on ramipril 5mg daily, I would increase this to 10mg to better optimize his blood pressure before surgery, provided his renal function remains stable. This modification would help manage his cardiovascular risk during the procedure.

While his liver disease and cultural concerns are crucial considerations, they are less modifiable in the immediate preoperative period. Instead, I would focus on these three actionable interventions that could significantly improve his surgical outcomes within a relatively short timeframe.

**Examiner:** Is his COPD control adequate and, in addition to smoking cessation, how could it be improved?

**Doctor:** No, Mr. Wilson's COPD control is not adequate, and there are several important ways we can optimize his respiratory function. While he only uses his salbutamol inhaler once weekly, suggesting relatively stable symptoms, this reflects undertreatment rather than good control, especially given his ongoing smoking and the occasional wheeze noted on examination.

I would start him on a long-acting bronchodilator, specifically a long-acting muscarinic antagonist (LAMA) such as tiotropium, as this is indicated as first-line maintenance therapy for COPD. This will help improve his baseline respiratory function and reduce his reliance on salbutamol. Additionally, I would arrange for spirometry testing to properly assess his lung function and definitively stage his COPD, as this will guide further therapeutic decisions.

Given he has some wheeze on examination, I would also consider adding an inhaled corticosteroid/long-acting beta-agonist (ICS/LABA) combination if spirometry shows significant obstruction. However, the key to improving his respiratory status remains smoking cessation. Beyond the pharmacological smoking cessation support I mentioned earlier, I would provide him with clear data about how quickly lung function can improve after stopping smoking, particularly in the context of reducing his perioperative risks. This information might serve as additional motivation for quitting.

Importantly, I would also ensure he receives proper inhaler technique education, as incorrect technique could be contributing to suboptimal control. These interventions should significantly improve his respiratory function ahead of surgery.

**Examiner:** What further tests and information would you want about the state of Mr Wilson’s cirrhosis prior to the procedure?

**Doctor:** A comprehensive assessment of Mr. Wilson's cirrhosis is essential for surgical risk stratification. I would first obtain a complete liver function panel, coagulation profile, and full blood count to calculate his Model for End-Stage Liver Disease (MELD) score and Child-Pugh classification. These scores are crucial predictors of perioperative mortality and will significantly influence our surgical approach.

I would also arrange an upper endoscopy to assess for esophageal varices, as their presence would indicate portal hypertension and increase his bleeding risk during surgery. Additionally, I would order a liver ultrasound with Doppler studies to evaluate portal vein flow and screen for ascites that might not be clinically apparent. This imaging would also help exclude hepatocellular carcinoma, which is an important consideration in cirrhotic patients.

Given his history of alcohol use, with his last drink being at Christmas, I would obtain carbohydrate-deficient transferrin levels to confirm his reported alcohol abstinence. I would also check his alpha-fetoprotein level as part of hepatocellular carcinoma screening, and assess his nutritional status through albumin, prealbumin, and vitamin D levels, as malnutrition significantly impacts surgical outcomes in cirrhotic patients.

This comprehensive evaluation will allow us to better understand his liver synthetic function, assess his portal hypertension status, and optimize his nutritional status before surgery. These factors are critical in determining both his surgical risk and the potential need for additional perioperative support.

**Examiner:** How would you approach advanced care planning with Mr Wilson in the preoperative setting?

**Doctor:** Given Mr. Wilson's cultural background and the trauma surrounding his father's death during cardiac surgery, I would approach advance care planning with particular sensitivity to his family-centered decision-making preferences. After acknowledging his understandable anxieties about surgery, I would explain that discussing advance care planning is a standard part of preparing for any major procedure, emphasizing that this planning helps ensure his wishes and values are respected.

I would begin by exploring his understanding of the surgery's risks and benefits, then specifically ask about his goals of care and what matters most to him. Given his clear desire for family involvement, I would suggest arranging a dedicated family meeting with his siblings present, either in person or via teleconference, to discuss these matters together. During this conversation, I would incorporate an Aboriginal Health Liaison Officer to ensure cultural safety and appropriate communication.

The discussion would focus on practical aspects like appointing a medical power of attorney from among his siblings, documenting specific wishes about life-sustaining treatments, and determining his preferences for postoperative care. I would ensure we address his concerns about being away from country during recovery and explore options for returning to Murray Bridge as soon as medically appropriate.

Importantly, I would frame this planning as a way of giving him greater control over his care and ensuring his cultural and personal values are respected, rather than focusing on worst-case scenarios. This approach should help reduce his anxiety while establishing clear directives for the healthcare team.

**Examiner:** How would you go about managing Mr Wilson if he declines intervention on the aneurysm?

**Doctor:** If Mr. Wilson declines aneurysm repair, I would implement a structured conservative management plan while continuing to address his concerns about surgery. Given his aneurysm's size of 6.5cm and rapid 1cm growth over six months, I would be transparent about his high risk of rupture, while respecting his autonomy and cultural preferences.

I would optimize his medical management by ensuring strict blood pressure control, aiming for systolic pressures below 130mmHg through uptitration of his ramipril. I would also add a beta-blocker, which may help reduce aneurysm expansion rate. Given his elevated cardiovascular risk, I would increase his atorvastatin to 40mg daily, as statin therapy might also slow aneurysm growth.

Regular monitoring would be essential. I would arrange monthly ultrasound surveillance of his aneurysm size and schedule regular reviews in Murray Bridge to monitor his symptoms and vital signs. I would provide clear instructions about warning symptoms of rupture and ensure he and his family understand when to seek immediate medical attention.

Most importantly, I would maintain ongoing dialogue about his decision, working with an Aboriginal Health Liaison Officer to address his concerns about leaving country for surgery. I would also explore options for having his family more involved in his care planning, as this might help him feel more supported in eventually accepting surgical intervention. This approach balances respect for his autonomy while keeping the door open for future reconsideration of surgery if he becomes more comfortable with the idea.

##### Examiner 2

**Examiner:** What resources in the rural setting would you use to help implement your treatment plan?

**Doctor:** As Mr. Wilson's internal medicine specialist, I would actively engage several key local resources in Murray Bridge to implement his treatment plan while respecting his strong desire to stay connected to country and family.

I would first establish a partnership with the local Aboriginal Health Service to provide culturally appropriate care coordination. They can facilitate family meetings about his aneurysm management and help address the trauma related to his father's passing. Their Aboriginal Health Workers can provide essential cultural support and help maintain continuity of care between Murray Bridge and Adelaide.

I would collaborate with the Murray Bridge Hospital for aspects of his pre-operative workup and post-operative monitoring. While the aneurysm surgery itself requires tertiary care in Adelaide, we can conduct many pre-operative assessments locally, including blood work, ECGs, and some imaging studies. This approach would minimize his time away from country.

For ongoing management of his multiple chronic conditions, I would engage the local diabetes educator and dietitian to improve his glycemic control, and connect him with the visiting podiatrist for regular foot care. The local smoking cessation service can provide behavioral support and nicotine replacement therapy. Additionally, I would coordinate with the visiting specialist clinics that come to Murray Bridge, particularly for gastroenterology follow-up regarding his cirrhosis.

This coordinated local approach, centered around his cultural needs and family involvement, would help overcome his reluctance to engage with the healthcare system while ensuring he receives comprehensive care for both his acute surgical needs and chronic conditions.

**Examiner:** How ready does Peter feel for surgery? Other than involving his family, what culturally appropriate strategies would you use to get Peter on board?

**Doctor:** From my assessment, Peter is ambivalent about surgery due to deeply rooted concerns stemming from his father's death following cardiac surgery and his strong connection to country. His hesitation is evident in his reluctance to travel to Adelaide and his repeated emphasis on wanting more time to discuss the procedure.

To help Peter feel more prepared, I would implement several culturally appropriate strategies. First, I would arrange for an Aboriginal Hospital Liaison Officer to be his primary point of contact throughout the surgical journey. This would provide cultural continuity and support, particularly during his time in Adelaide. The Liaison Officer could help maintain his connection to country while in the hospital and ensure cultural practices are respected throughout his care.

Second, I would coordinate with the Aboriginal Health Service to arrange a traditional healer or Elder to be involved in his care planning, if Peter is comfortable with this. Their involvement could help bridge traditional healing practices with western medicine, providing spiritual and cultural support that may help Peter feel more secure in proceeding with surgery.

Third, I would work with the vascular surgery team to create a clear timeline for Peter's return to country post-surgery. Having a defined plan for when he can return to Murray Bridge would help address his anxiety about being away from his community. Additionally, I would explore options for some post-operative care to be provided locally through the Murray Bridge Hospital, minimizing his time away from country.

These strategies, combined with family involvement, would help create a more culturally safe environment for Peter to proceed with his necessary surgery.

**Examiner:** How would Peter's MELD score potentially influence the peri-operative planning and outcomes of his surgery?

**Doctor:** As an internal medicine specialist, I would emphasize that Peter's MELD score is crucial for surgical planning, though we currently lack some key data points for its calculation. Based on his clinical presentation showing compensated cirrhosis without obvious ascites, jaundice, or encephalopathy, I anticipate his MELD score would likely be in the lower range, but we need current laboratory values to confirm this.

If his MELD score is elevated, this would significantly impact perioperative planning in several ways. A high MELD score would indicate increased risks of bleeding complications during surgery due to potential coagulopathy, and would necessitate careful preparation with blood products and potentially vitamin K supplementation. It would also signal higher risks of post-operative hepatic decompensation and acute kidney injury.

The MELD score would influence our choice of anesthetic agents and post-operative pain management strategy, as many medications rely on hepatic metabolism. We would need to adjust medication dosing accordingly and closely monitor liver function in the post-operative period. Additionally, a higher MELD score would require more intensive post-operative monitoring for complications like ascites and hepatic encephalopathy.

Given these considerations, I would obtain urgent liver function tests, coagulation studies, and renal function tests to calculate his current MELD score. This would allow us to optimize his pre-operative management and ensure appropriate resources are available during and after surgery to manage potential complications related to his liver disease.

**Examiner:** Which diabetic medication changes would you make to optimise Peter's care, and what considerations would you factor into your decision?

**Doctor:** Given Peter's upcoming aneurysm surgery and his suboptimal glycemic control with an HbA1c of 7.9% and blood sugars ranging from 7-16 mmol/L, I would make several targeted medication adjustments while considering his liver and kidney disease.

I would increase his gliclazide MR from 60mg to 120mg daily, as this medication is generally safe in both renal and hepatic impairment. However, I would maintain his current metformin XR dose at 1g daily rather than increasing it, given his moderate renal impairment with eGFR 30-60. We need to be cautious about metformin accumulation and potential lactic acidosis, particularly in the perioperative period.

For the immediate perioperative period, I would transition him to a basal-bolus insulin regimen. This would typically involve starting him on glargine insulin at night with rapid-acting insulin before meals, allowing for tighter glycemic control during the surgical period. The doses would be calculated based on his weight of 82kg, starting at approximately 0.3 units/kg/day total insulin.

These medication changes consider several key factors: his chronic kidney disease limiting metformin dosing, his compensated cirrhosis affecting medication metabolism, the need for optimal glycemic control to reduce surgical complications and promote wound healing, and the practicality of medication management during his hospital admission. We would need to monitor his renal function closely and adjust medications accordingly during the perioperative period.

**Examiner:** Are there any other pre-operative considerations and risks that should be addressed before surgery?

**Doctor:** As Mr. Wilson's treating physician, I would prioritize several key pre-operative considerations to optimize his surgical outcomes, focusing on both medical and cultural aspects.

From a medical perspective, his ongoing smoking represents a significant modifiable risk factor that requires immediate attention. I would initiate nicotine replacement therapy and strongly encourage smoking cessation, even for a brief period pre-operatively, to reduce his respiratory complications risk. Additionally, I would obtain pulmonary function tests to better quantify his COPD severity and optimize his respiratory medication regimen.

His cardiovascular risk needs careful assessment given his diabetes, hypertension, and chronic kidney disease. I would arrange cardiac stress testing to evaluate for underlying coronary artery disease that might complicate his surgery. His blood pressure control should be optimized, aiming for targets that balance his cardiovascular and renal health.

From a cultural safety perspective, we must address his psychological readiness for surgery, particularly his anxiety related to his father's death. I would ensure an Aboriginal Liaison Officer is involved from the pre-operative period and arrange for traditional healing practices to be incorporated into his care plan if he desires.

Most urgently, given his rapid aneurysm growth, I would expedite these pre-operative assessments while simultaneously working with his family and cultural supports to ensure he feels safe proceeding with surgery. This balanced approach addresses both his medical risks and cultural needs to optimize his surgical outcomes.

#### Case 5 – Type 1 Diabetes mellitus and driving

##### Examiner 1

**Examiner:** Specifically, what would you do to improve his glycemic control?

**Doctor:** I would implement an intensive diabetes optimization plan focused on three key areas. First, I would adjust his insulin regimen, increasing his basal Glargine to 26 units at bedtime and implementing a more structured carbohydrate-counting approach for his Aspart dosing. This change acknowledges his farming schedule while providing better coverage for his needs.

Second, I would provide him with a continuous glucose monitoring system. This is essential given his hypoglycemic unawareness and irregular monitoring schedule. The constant availability of glucose data and predictive alerts would help him manage his blood sugar more effectively while working on the farm, and critically, would improve safety during driving and machinery operation. I would ensure he and his wife are thoroughly trained in using the system and responding to alerts.

Third, I would establish a simplified meal plan that accounts for his farming schedule. Rather than expecting perfect carbohydrate counting during busy periods, I would create a modified basal-bolus regime with pre-calculated insulin doses for typical meals he can manage while working. I would ensure he has easily accessible hypoglycemia treatment supplies in all vehicles and machinery, with clear instructions for his wife and neighboring farmers on emergency response protocols.

This plan balances the need for improved glycemic control with the practical constraints of his farming lifestyle. While it may not achieve perfect control, it prioritizes safety and sustainability, particularly regarding his crucial need to continue driving and operating machinery safely. I would schedule monthly reviews initially to fine-tune this approach based on his continuous glucose monitoring data and real-world experience.

**Examiner:** What barriers do you foresee in this patient being a candidate for kidney transplant?

**Doctor:** I would identify four significant barriers to kidney transplantation for Mr. Mitchell. The most pressing is his poorly controlled diabetes with an HbA1c above 8.5%. I would need to demonstrate sustained improvement in his glycemic control before transplant centers would likely consider him a candidate, as poor control significantly increases post-transplant complications risk.

His current alcohol consumption of 5-6 standard drinks daily presents another major barrier. Most transplant programs require either abstinence or significant reduction in alcohol intake. His current consumption could impact both his candidacy for transplantation and his ability to reliably comply with post-transplant care requirements.

His untreated mental health conditions, particularly his worsening depression and PTSD, constitute the third barrier. These conditions need to be stabilized and appropriately managed before transplantation could be considered. The lack of regular mental health support makes this particularly challenging given his rural location.

Finally, his limited social support network is concerning. While his brother has offered to be a donor, which is positive, Mr. Mitchell's rural location and primary caregiver responsibilities for his farm and family could complicate post-transplant care adherence. The distance to transplant centers and ongoing care facilities would make frequent post-transplant monitoring challenging.

I would address these barriers by first focusing on diabetes control and alcohol reduction, while simultaneously establishing regular mental health support through telehealth services. However, I would also begin early discussions with transplant services to understand their specific requirements, allowing us to work toward meeting these criteria while managing his advancing kidney disease.

**Examiner:** How would you go about supporting this patient to reduce his alcohol use?

**Doctor:** I would take a comprehensive but practical approach to addressing Mr. Mitchell's alcohol use, recognizing that his intake significantly impacts both his current health and future transplant candidacy. Initially, I would have a frank discussion about how his current consumption of 5-6 standard drinks daily affects his diabetes control, kidney disease progression, and medication effectiveness. I would specifically highlight how alcohol interferes with his ability to recognize and respond to hypoglycemic episodes, which is particularly dangerous given his work involving machinery.

Rather than immediately pushing for complete abstinence, I would collaborate with him to set achievable reduction goals. Given his rural location and family responsibilities, I would utilize telehealth services to connect him with an addiction specialist for regular support sessions that can fit around his farming schedule. I would also prescribe thiamine supplementation and monitor closely for withdrawal symptoms as he reduces his intake.

Critically, I would integrate alcohol reduction strategies with his other health priorities. For example, I would help him identify high-risk periods for drinking, such as after stressful farm work, and develop alternative coping strategies. I would involve his wife in these discussions with his permission, as her support will be crucial. Additionally, I would coordinate this alcohol reduction plan with his mental health treatment, as his increased drinking appears linked to his worsening depression.

The goal would be to reduce his alcohol intake to levels acceptable for transplant candidacy while providing him with sustainable alternative coping mechanisms that acknowledge his challenging circumstances.

**Examiner:** Do you need to stop this patient driving?

**Doctor:** Based on my assessment today, I would have to immediately restrict Mr. Mitchell from driving due to several critical safety concerns. The combination of his documented hypoglycemic unawareness, his recent episode of asymptomatic severe hypoglycemia (2 mmol/L), and his irregular glucose monitoring before driving creates an unacceptable risk to both himself and others on the road. Additionally, his progressive diabetic retinopathy requiring urgent ophthalmology review, combined with his reduced visual acuity, particularly in his right eye (6/12), further compounds these safety concerns.

However, I deeply understand the devastating impact this decision has on his livelihood and family responsibilities. Therefore, I would implement a clear pathway for him to return to driving. This would require him to demonstrate consistent blood glucose monitoring before driving, with documented readings above 5 mmol/L, use of continuous glucose monitoring with alarms, and completion of a hypoglycemia awareness retraining program. He would also need urgent ophthalmology review and treatment of his retinopathy.

In the interim, I would work with him to identify alternative transportation arrangements, possibly involving his wife or neighboring farmers, while expediting the necessary medical reviews and interventions. My goal would be to help him return to driving as quickly as possible, but only when it's safe for him to do so. This requires balancing our duty of care to public safety with recognition of the critical importance of driving to his work and family life.

**Examiner:** What are the consequences of this patient’s disease for his family, both now and potentially in the future?

**Doctor:** Mr. Mitchell's medical conditions are creating significant immediate and long-term consequences for his family. Currently, his wife and young child face increasing uncertainty about their financial stability as his medical conditions threaten his ability to work effectively on the farm. The immediate driving restriction I've had to implement today will create substantial practical challenges for managing both the farm operations and his medical care, likely requiring his wife to take on additional responsibilities for transportation.

Looking to the near future, his progressing kidney disease will require dialysis or transplantation, which will significantly impact family dynamics. The rural location means either lengthy travel times to dialysis centers, potentially three times weekly, or consideration of relocating closer to medical facilities - either option creates substantial disruption to their farming lifestyle and their child's stability. His wife may need to take on more farm management responsibilities or consider additional employment to maintain financial stability.

The longer-term outlook presents even more challenges. His diabetic complications and mental health issues could progress to a point where he becomes unable to work, potentially forcing difficult decisions about the farm's future - their primary asset and source of income. His young child may need to adapt to a significantly changed lifestyle and potentially take on caring responsibilities at a young age. Additionally, the financial strain of managing his complex medical conditions could impact their ability to save for their child's education or their own retirement.

These impacts underscore why maintaining his ability to work safely must be a central consideration in all our medical decisions, while we simultaneously plan for potential future scenarios that his family may face.

##### Examiner 2

**Examiner:** You mentioned your patient was diagnosed at age 12, with type 1 diabetes. What are the relevant serum antibodies that may be tested to help differentiation between type 1 and type 2 diabetes in patients presenting with a new likely diagnosis of diabetes mellitus?

**Doctor:** In differentiating between Type 1 and Type 2 diabetes, I typically look for four key autoantibodies that are characteristic of Type 1 diabetes. The most sensitive and specific is anti-glutamic acid decarboxylase (anti-GAD) antibody, which is present in about 80% of Type 1 diabetes cases at diagnosis. I also test for islet cell antibodies (ICA), which were historically the first identified and remain important diagnostically. Additionally, I check for insulin autoantibodies (IAA), though these must be tested before insulin therapy is initiated as they become less useful after insulin exposure. The fourth antibody I look for is anti-islet antigen 2 (IA-2A), which is particularly helpful in confirming Type 1 diabetes when positive alongside other antibodies.

I find testing for zinc transporter 8 autoantibodies (ZnT8A) particularly valuable in cases where other antibody results are equivocal, as it can help identify an additional 10-15% of Type 1 cases that might otherwise be missed. The presence of multiple positive antibodies strongly supports a Type 1 diagnosis, though I always interpret these results in conjunction with clinical presentation, age of onset, and family history.

In Mr. Mitchell's case, while antibody testing would have been valuable at his initial diagnosis, his clinical presentation at age 12 with classical symptoms and subsequent disease course has clearly confirmed his Type 1 diabetes status.

**Examiner:** Should your patients renal function continue to deteriorate, renal replacement therapy may be required though, as you mentioned, it is difficult in the setting of his remote residence. What are some possible alternative renal replacement therapy options to hospital-based or unit-based dialysis?

**Doctor:** For Mr. Mitchell, I would strongly advocate for home-based peritoneal dialysis as our primary renal replacement therapy option. Given his remote location and need to maintain his farming work, this modality would provide him the greatest flexibility while minimizing travel burden. Peritoneal dialysis can be performed overnight using an automated cycler, allowing him to maintain his daytime farming activities. This approach would also help preserve his independence and ability to continue working, which is crucial for his family's financial stability.

If peritoneal dialysis isn't suitable, I would then consider home hemodialysis as our second option. While this requires more extensive training and home modifications, it would still allow him to dialyze on his own schedule without regular travel to a distant center. The initial investment in training time – typically 6-8 weeks – would be offset by the long-term benefits of treatment autonomy and better clinical outcomes compared to in-center hemodialysis.

Both these home-based options would need to be supported by a robust remote monitoring system and clear protocols for emergency situations. I would ensure we have a solid training plan in place, involving his wife in the training process, and establish clear communication channels with our dialysis team. The ultimate goal would be to maintain his ability to work while ensuring adequate dialysis delivery, making home-based therapy the most practical solution for his situation.

**Examiner:** What are some of the nuances that must be considered when prescribing and managing a patient with poorly controlled diabetes mellitus undergoing peritoneal dialysis?

**Doctor:** Managing diabetes in the context of peritoneal dialysis requires careful attention to several key factors. First, I need to account for glucose absorption from the dialysate, which can contribute an additional 100-200 grams of glucose per day to Mr. Mitchell's system. This constant glucose exposure will likely increase his insulin requirements and could further complicate his already poor glycemic control.

I would need to adjust his insulin regimen to account for the timing of his dialysis exchanges. For overnight automated peritoneal dialysis, I would increase his basal insulin during the dialysis period to counteract the glucose load. His current insulin doses – 22 units of Glargine and 8-10 units of Aspart with meals – would likely need significant upward adjustment.

The presence of peripheral neuropathy and retinopathy makes accurate self-administration of both insulin and peritoneal dialysis more challenging. Given his hypoglycemic unawareness, I would set higher blood glucose targets (7-10 mmol/L) to ensure safety while performing farm work. I would also implement a strict monitoring protocol before and during dialysis exchanges.

Additionally, I need to consider the impact of uremia on insulin sensitivity and the altered pharmacokinetics of insulin in renal failure. The risk of peripheral edema from peritoneal dialysis could also affect insulin absorption. Therefore, I would start with more conservative insulin adjustments and titrate based on frequent blood glucose monitoring, aiming to balance diabetes control with his need to work safely on the farm.

**Examiner:** You mentioned that your patient does not attend psychotherapy given difficulty accessing this service in his rural location. Given he is unable to attend in-person psychotherapy, what are some alterative non-pharmacological management options that may benefit your patient's depression and PTSD?

**Doctor:** For Mr. Mitchell's mental health management, I would prioritize implementing telehealth-based cognitive behavioral therapy through our state's rural and remote mental health service. This would allow him to access structured psychological support from his farm, eliminating travel barriers while providing evidence-based treatment for both his depression and PTSD. I would aim to schedule these sessions around his farm work commitments to ensure consistent engagement.

I would also strongly recommend a digital mental health platform that combines mood tracking with guided self-help modules. These platforms offer validated therapeutic techniques that he can access at any time, particularly useful during periods of increased stress or when PTSD symptoms emerge during farm work. The digital format would allow him to progress at his own pace while providing us with objective data about his mental health status.

Given his wife's involvement in his care, I would incorporate couples-based therapeutic techniques they can practice together, particularly focusing on communication strategies and shared problem-solving. This approach acknowledges his family context while building resilience within his primary support relationship. Additionally, I would teach him specific grounding techniques he can use independently during farming activities, especially useful for managing PTSD symptoms that may arise during work.

These interventions, combined with regular monitoring through our telehealth check-ins, would provide a comprehensive non-pharmacological support framework that respects both his rural location and need to maintain his farming responsibilities.

**Examiner:** You suggested several options to optimise the management of your patient's diabetes, to allow the ability to continue to drive. What is the likelihood of significantly improving your patient's hypoglycaemic unawareness, and do you feel your patient should not be allowed to drive?

**Doctor:** Based on the current evidence, I believe Mr. Mitchell should not continue driving until we achieve significant improvements in his diabetes management. His combination of hypoglycemic unawareness, irregular glucose monitoring, and only checking levels "about half the time" before driving presents an unacceptable risk to both himself and others. Additionally, his progressive retinopathy and peripheral neuropathy further compound these safety concerns.

While hypoglycemic unawareness can potentially be improved through strict avoidance of hypoglycemic episodes for several weeks, this would require a complete overhaul of his current diabetes management. Given his irregular monitoring, busy farming schedule, and current high alcohol intake, I do not believe we can achieve this safely while he continues to drive. The likelihood of successfully reversing his hypoglycemic unawareness is low unless we first establish a period of intensive blood glucose monitoring and management under conditions where he is not operating vehicles or machinery.

I recognize this position will significantly impact his livelihood and family responsibilities. However, as his physician, I must prioritize safety. I would immediately begin working with him to develop alternative transportation arrangements and adjust his farming operations while we implement strict glucose management protocols. Once we achieve stable glucose control with documented awareness of hypoglycemia and consistent monitoring, we can reassess his fitness to drive. This may take several months, but it's the only responsible course of action given his current clinical status.

### Supplementary Information 6 – Scorer sheets

#### Case 1 – Cystic fibrosis and lung transplant

##### Examiner 1

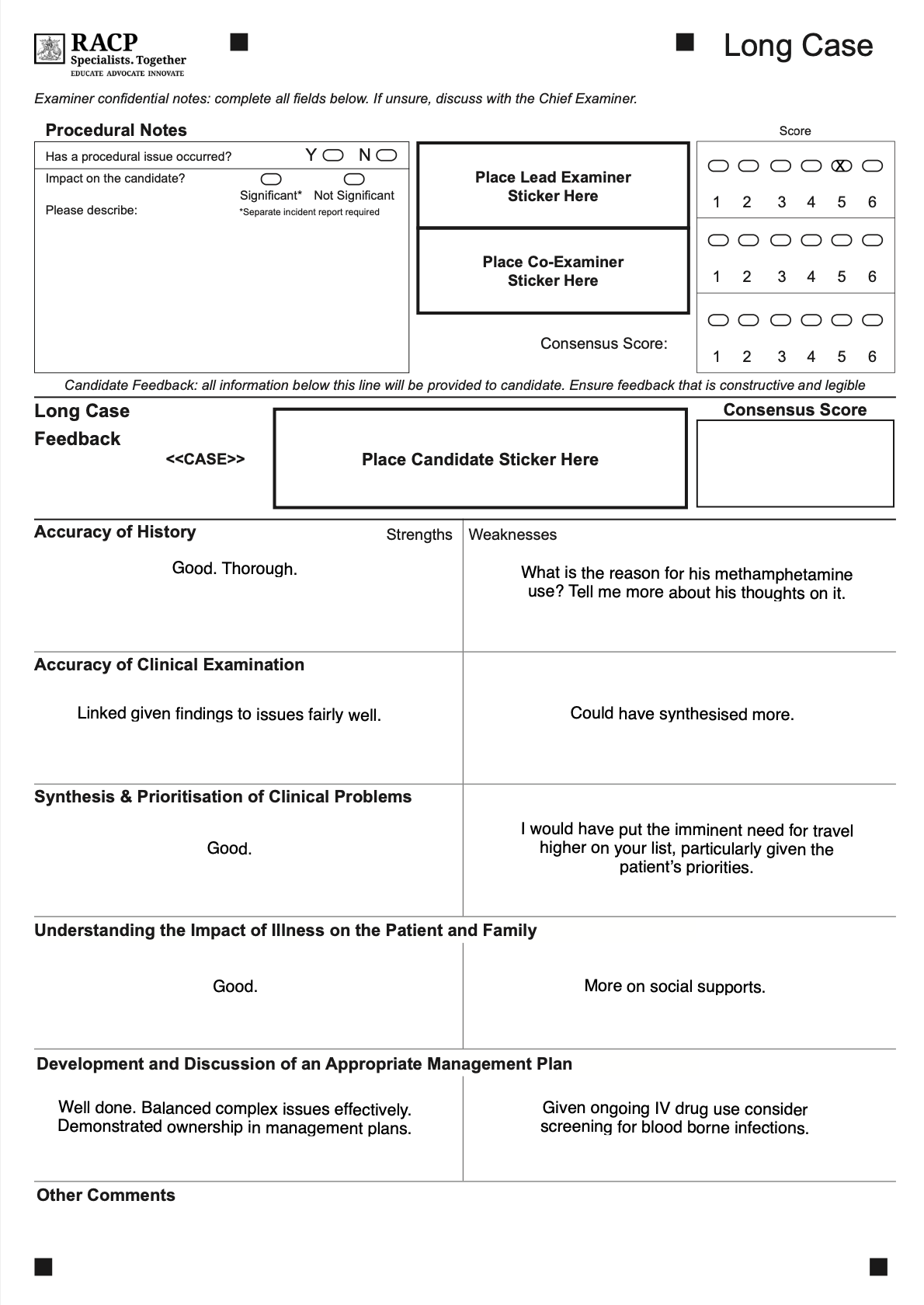

##### Examiner 2

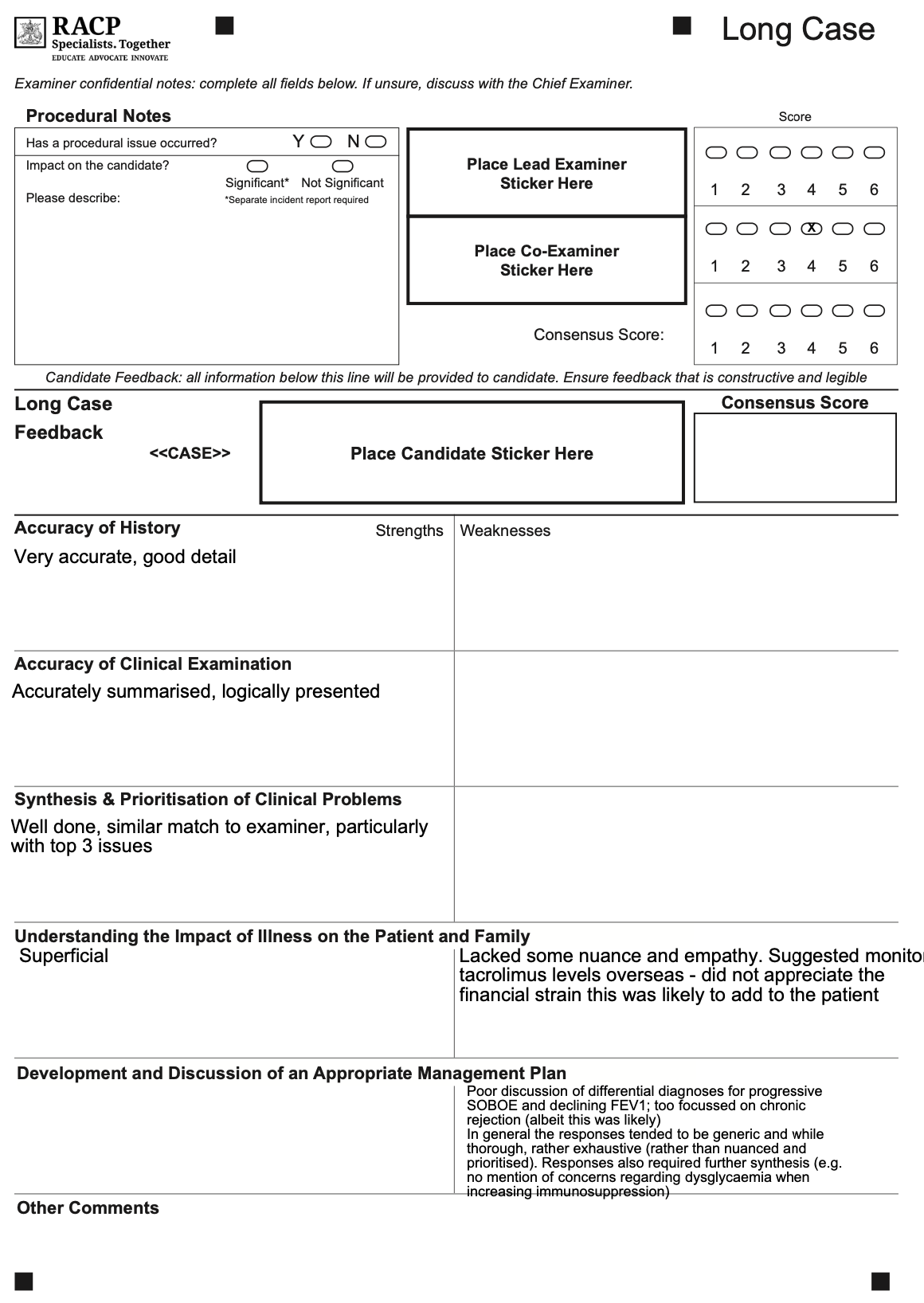

#### Case 2 – Heart Failure with reduced Ejection Fraction and falls

##### Examiner 1

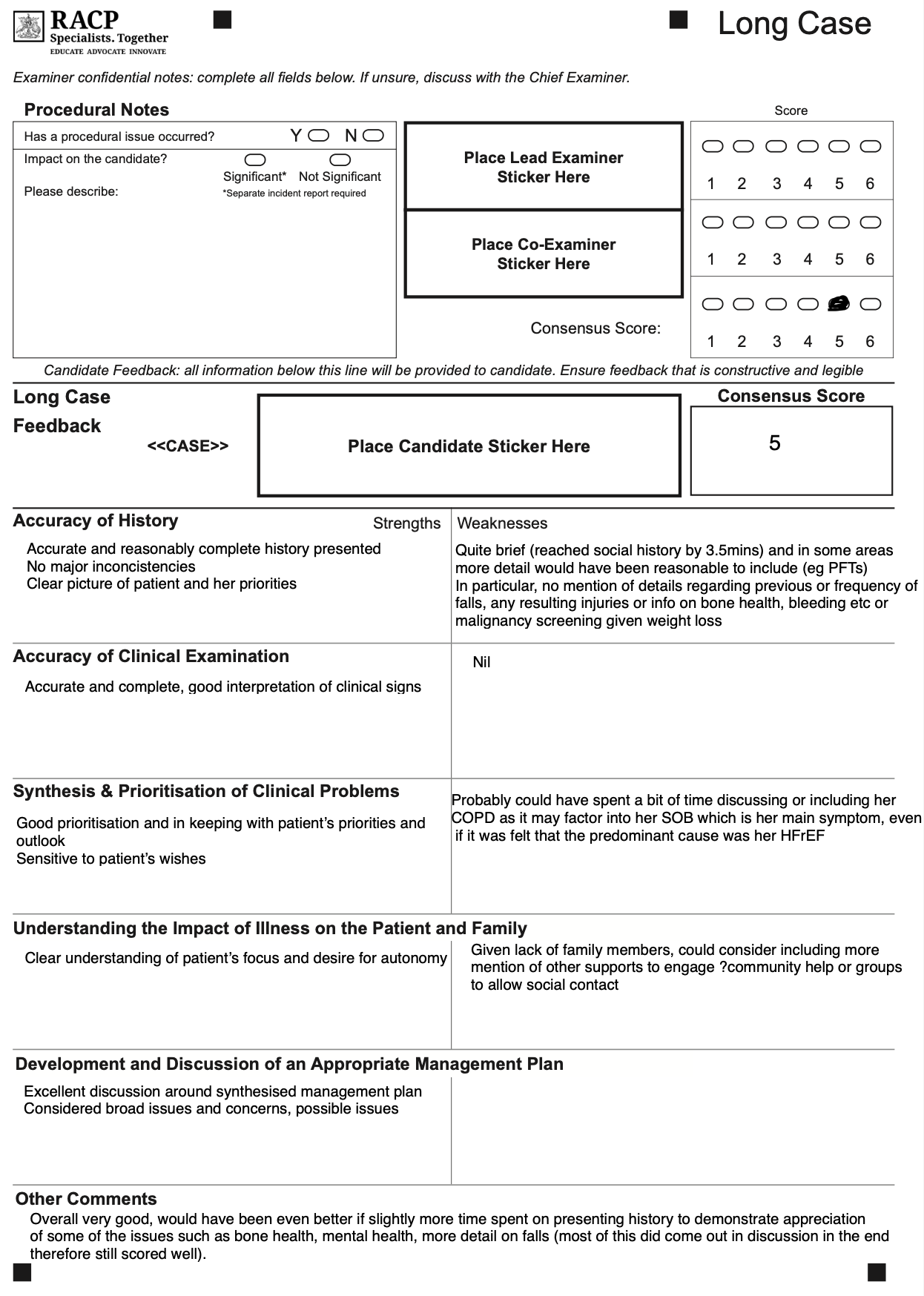

##### Examiner 2

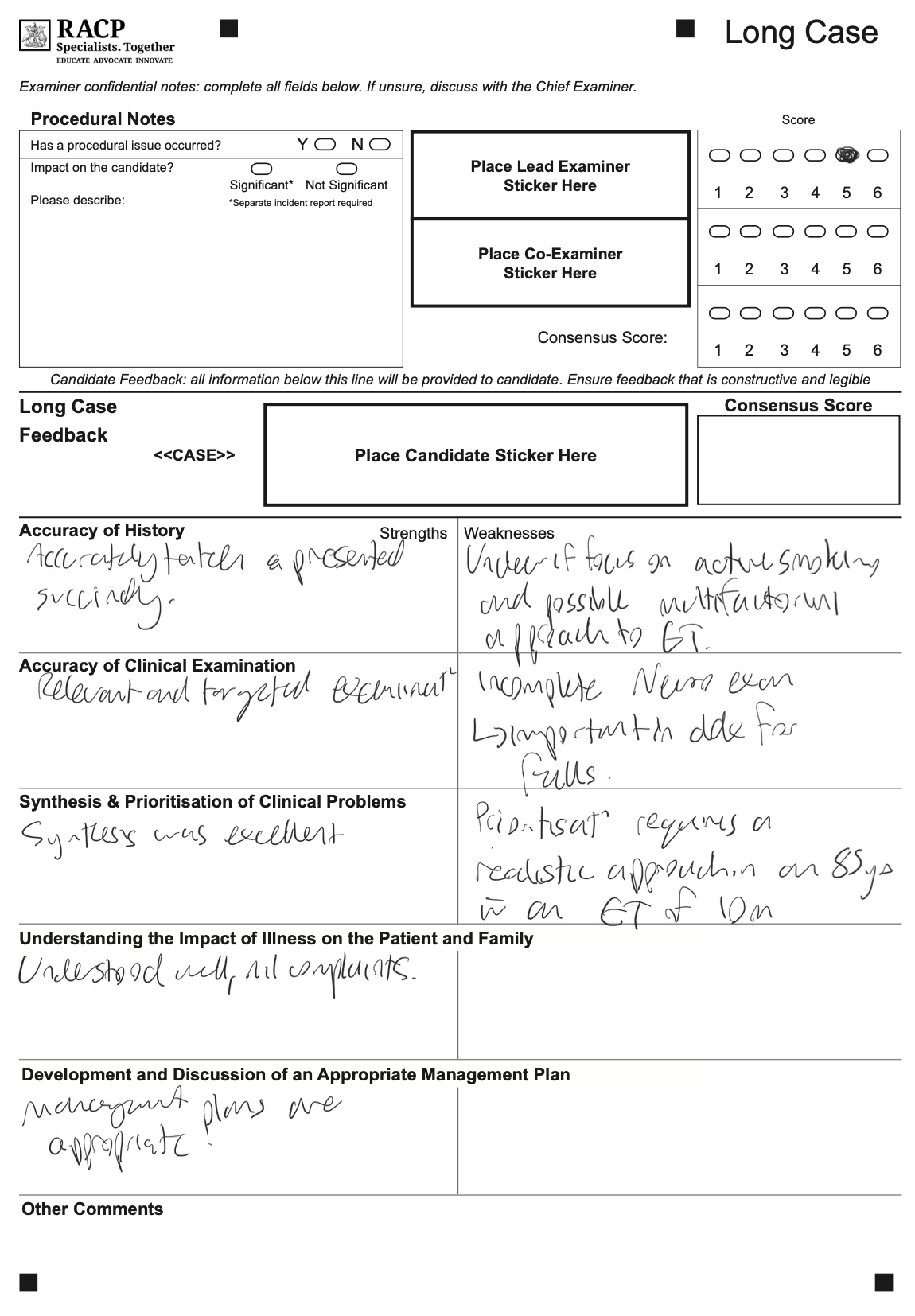

#### Case 3 – Myasthenia gravis and osteoporosis

##### Examiner 1

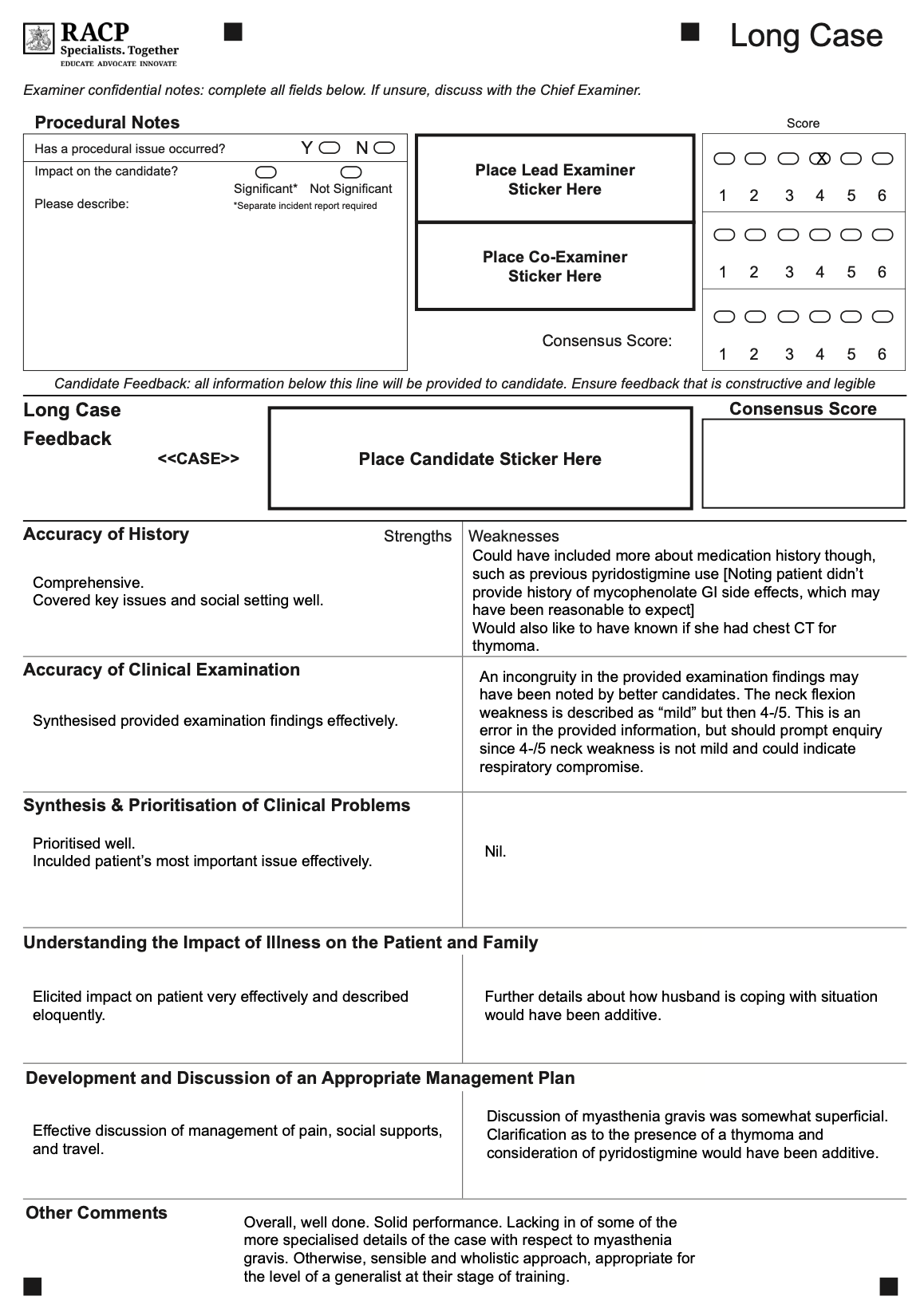

##### Examiner 2

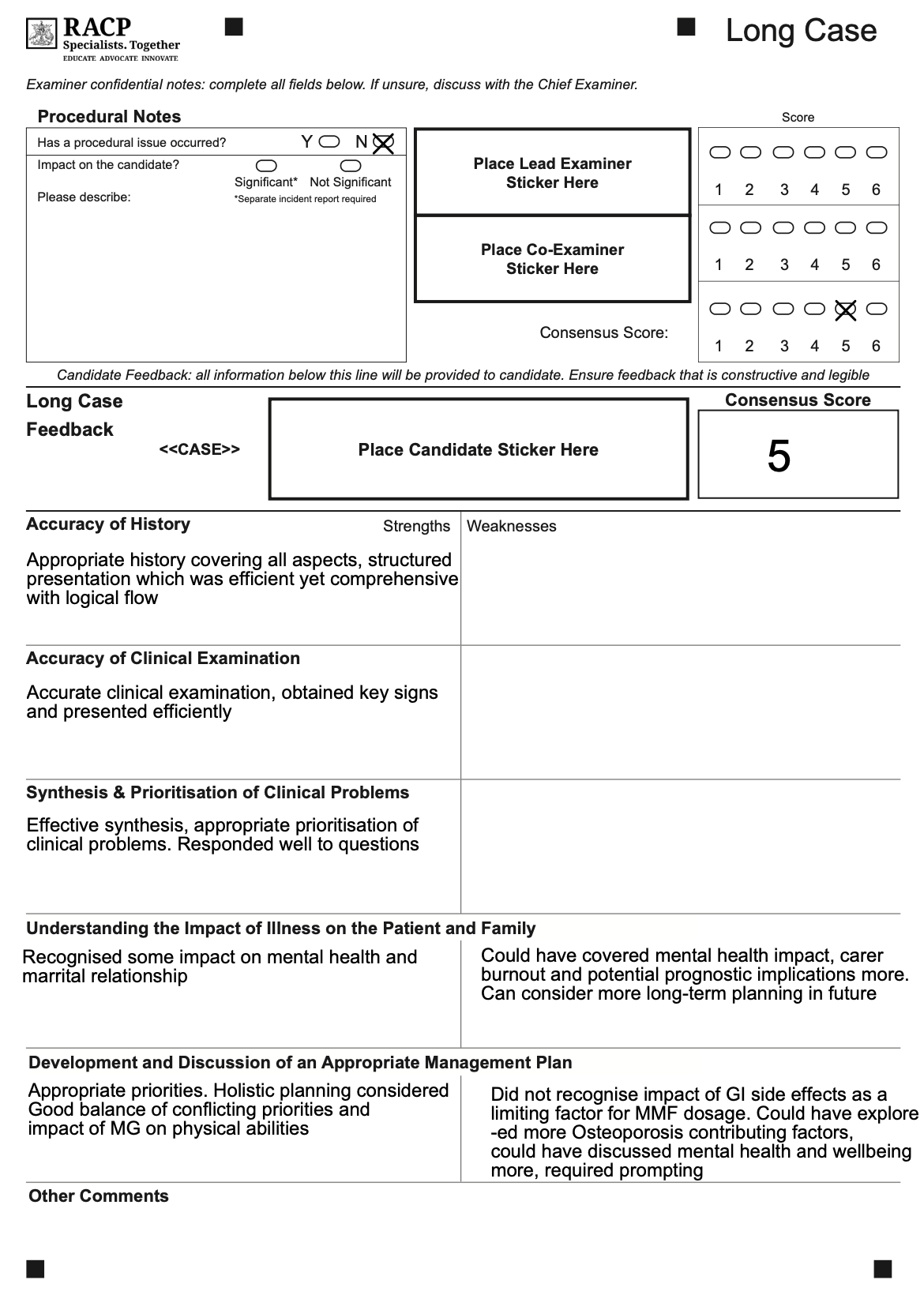

#### Case 4 – Perioperative and rural

##### Examiner 1

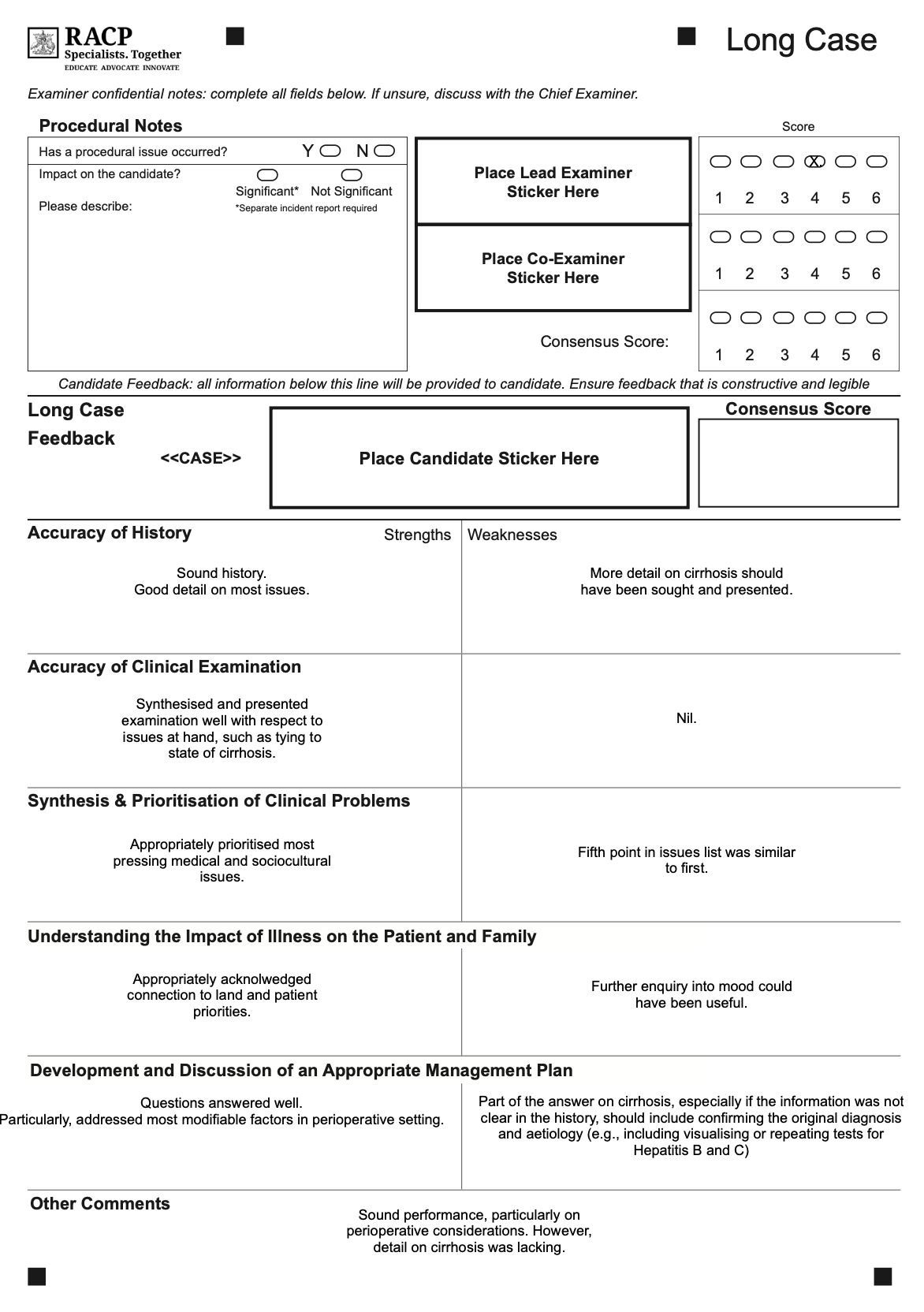

##### Examiner 2

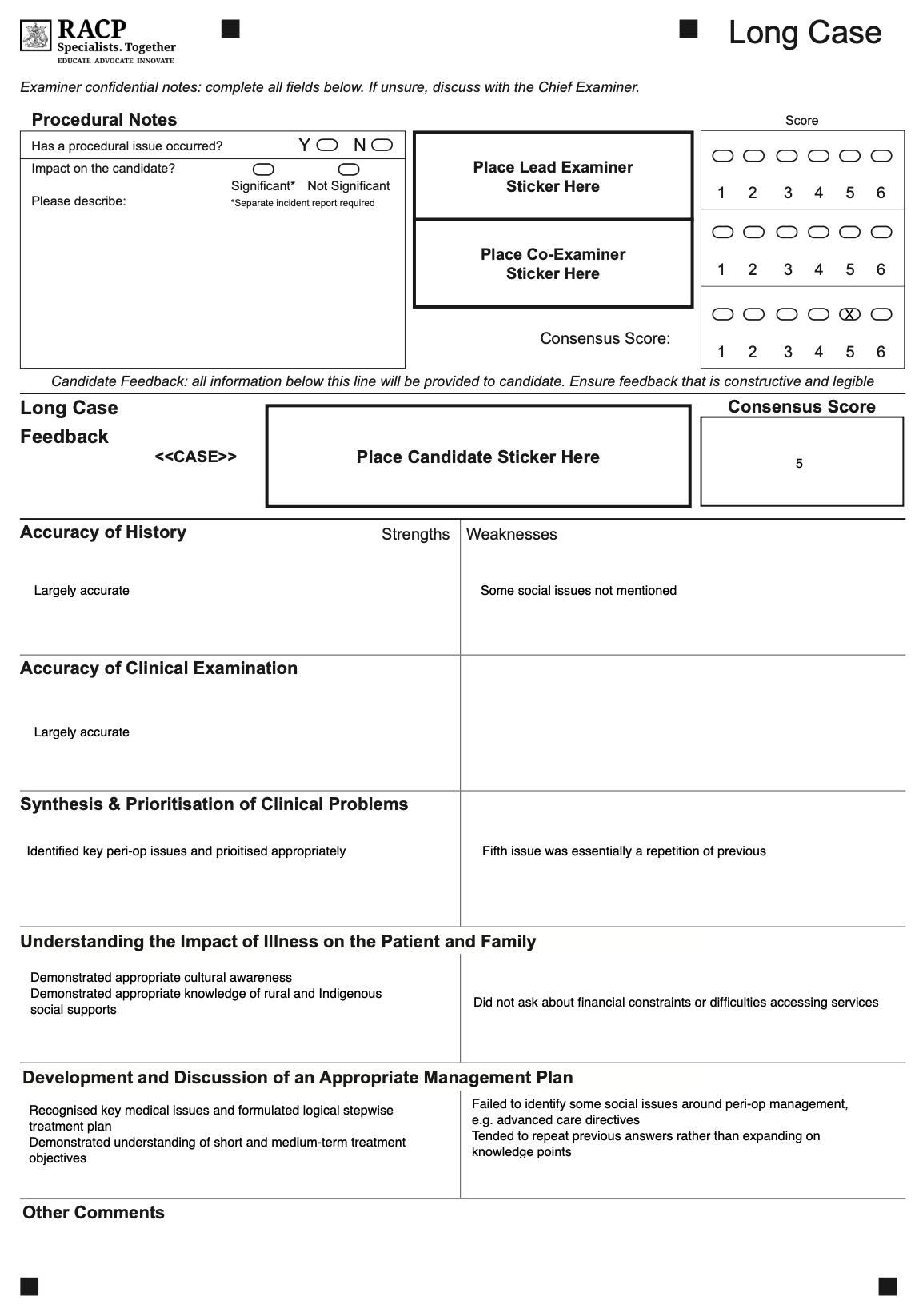

#### Case 5 – Type 1 Diabetes mellitus and driving

##### Examiner 1

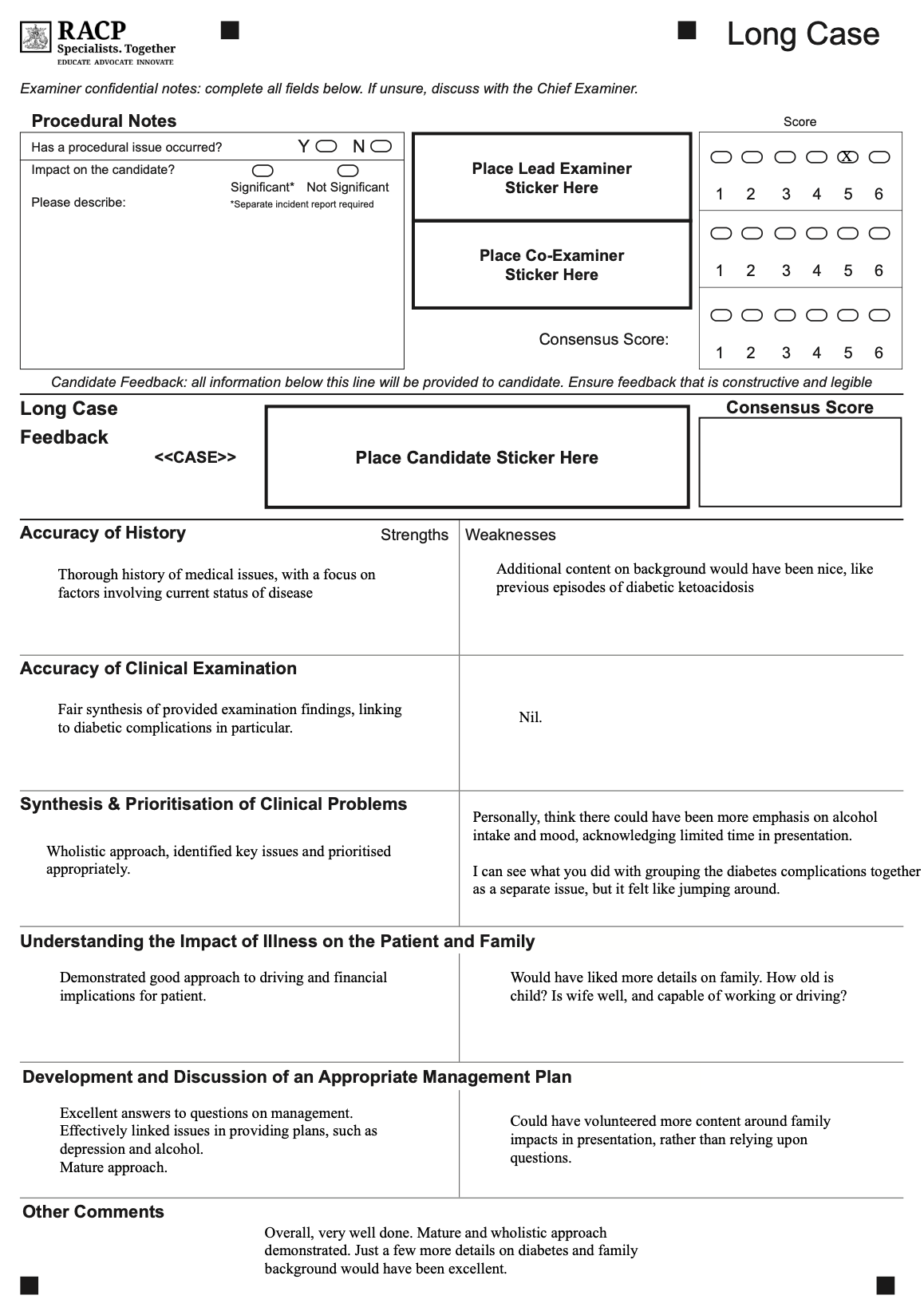

##### Examiner 2

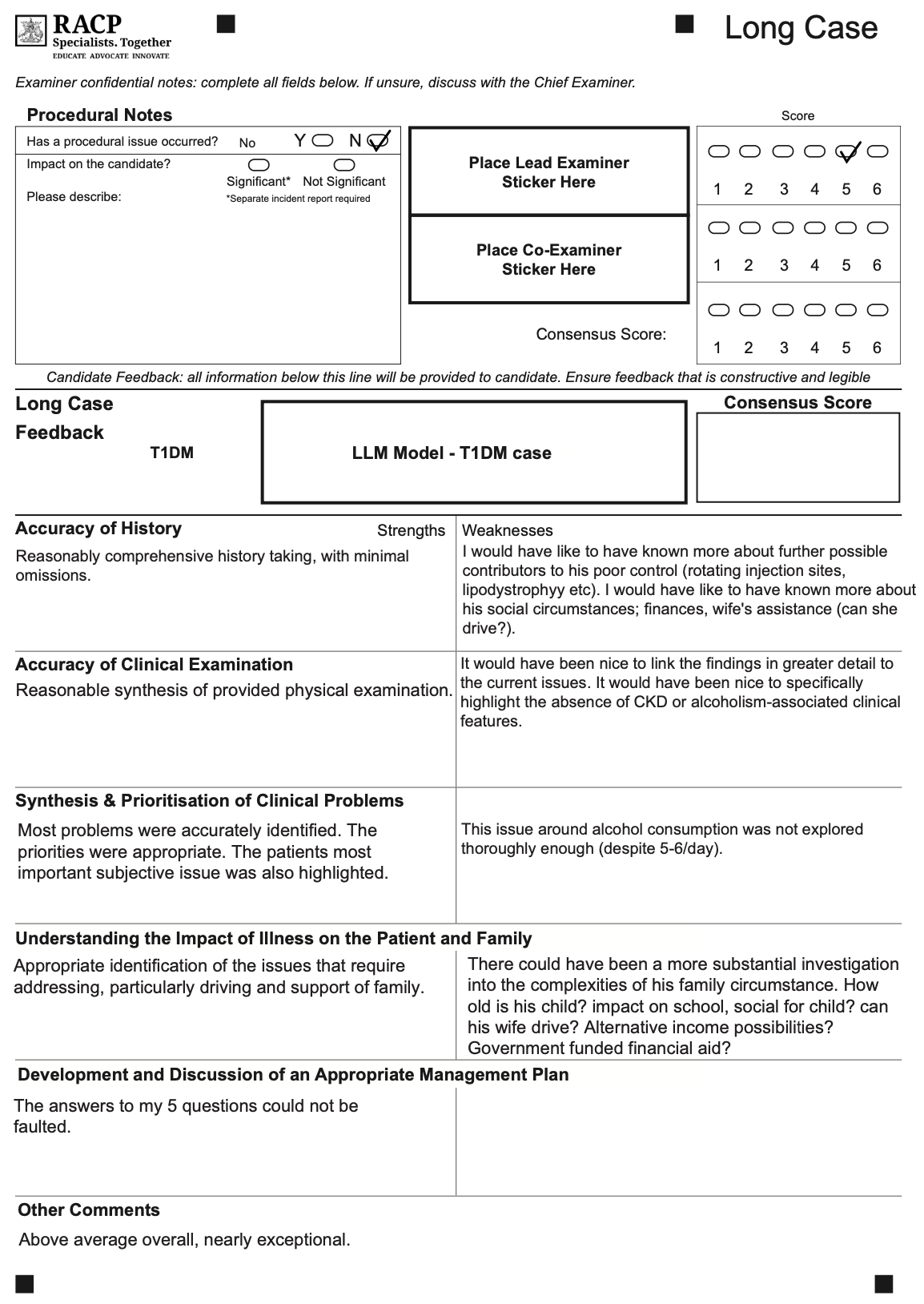
